# Metabolome-derived kidney function scores as prognostic readouts of cardiovascular-kidney-metabolic health

**DOI:** 10.64898/2026.11.09.26367636

**Authors:** Petros Andrikopoulos, Janis M. Nolde, Rima Chakaroun, Manyi Jia, Sofia K. Forslund-Startceva, Judith Aron-Wisnewsky, Trine Nielsen, Elena Butz, Peggy Sekula, Sara Vieira-Silva, Gwen Falony, Angelos Manolias, Antonis Myridakis, Julien Chilloux, Kanta Chechi, Elliot Murphy, Francesc Puig-Castellvi, Christine Rouault, Christine Poitou, Pierre Bel Lassen, Romina Pacheco-Tapia, Inés Castro-Dionicio, Eugeni Belda, Emmanuelle Le Chatelier, MetaCardis Consortium, GCKD Consortium, Matthias Blüher, Jens Nielsen, Jeroen Raes, Peer Bork, Lars Køber, Henrik Vestergaard, Torben Hansen, Philippe Froguel, Jean-Daniel Zucker, Mark Lathrop, Parminder Raina, Kai-Uwe Eckardt, Jean-Michel Oppert, S. Dusko Ehrlich, Fredrik Bäckhed, Anna Köttgen, Michael Stumvoll, Oluf Pedersen, Karine Clément, Marc-Emmanuel Dumas

**Affiliations:** Division of Systems Medicine, Department of Metabolism, Digestion and Reproduction, Imperial College London, London, W12 0NN, United Kingdom; Imperial – CNRS International Research Lab in Multiscale Metabolism “Antoine Lavoisier”, Imperial College London, Du Cane Road, London W12 0NN, UK; Department of Nephrology, Faculty of Medicine and Medical Centre - University of Freiburg, Freiburg, Germany; Institute of Epidemiology and Prevention, Department of Data Driven Medicine, Faculty of Medicine and Medical Center - University of Freiburg, Freiburg, Germany; Medical Department III –Endocrinology, Nephrology, Rheumatology, University of Leipzig Medical Center, Leipzig, Germany; The Wallenberg Laboratory, Department of Molecular and Clinical Medicine, Institute of Medicine, Sahlgrenska Academy, University of Gothenburg, Gothenburg, Sweden; LeiCeM - Leipzig Center of Metabolism, Leipzig University, Leipzig, Germany; Charité-Universitätsmedizin Berlin, Freie Universität Berlin and Humboldt-Universität zu Berlin, Berlin, Germany; Max Delbrück Center for Molecular Medicine in the Helmholtz Association (MDC), Berlin, Germany; Experimental and Clinical Research Center, Max Delbrück Center for Molecular Medicine and Charité-Universitätsmedizin Berlin, 26 Berlin, Germany; DZHK (German Centre for Cardiovascular Research), partner site Berlin, Berlin, Germany; Structural and Computational Biology Unit, EMBL, Heidelberg, Germany; Sorbonne Université, Inserm, Nutrition and Obesities: Systemic Approaches, NutriOmics, Research Unit, Paris, France; Assistance Publique - Hopitaux de Paris, Department of Nutrition, Pitié-Salpêtrière Hospital, Sorbonne University, Paris, France; Novo Nordisk Foundation Center for Basic Metabolic Research, Faculty of Health and Medical Sciences, University of Copenhagen, Copenhagen, Denmark; Steno Diabetes Center Copenhagen, Herlev, Denmark; Institute of Medical Microbiology and Hygiene and Research Centre for Immunotherapy (FZI), University Medical Centre of the Johannes Gutenberg-University Mainz, Germany; Institute of Molecular Biology (IMB), Mainz, Germany; Institute for Quantitative and Computational Biosciences (IQCB), Johannes Gutenberg University, Mainz, Germany; Translational Research of Gastrointestinal Disorders (TARGID), KU Leuven, Belgium; Host Microbe Interactomics Group, Wageningen University & Research, Wageningen, The Netherlands; University of Lille, Multi-omics and pathophysiology of metabolic disease unit, UMR8199 CNRS, U1283 INSERM, Institut Pasteur de Lille, Lille University Hospital, Lille 59045, France; Sorbonne Université, IRD, Unité de Modélisation Mathématique et Informatique des Systèmes Complexes, UMMISCO, Bondy, France; Metagenopolis, INRA, AgroParisTech, Université Paris-Saclay, Paris, France; Helmholtz Institute for Metabolic, Obesity and Vascular Research (HI-MAG) of the Helmholtz Zentrum München at the University of Leipzig and University Hospital Leipzig, Leipzig, Germany; Department of Life Science, Chalmers University of Technology, SE412 96 Gothenburg, Sweden; BioInnovation Institute, Ole Maaløes Vej 3, DK2200 Copenhagen N, Denmark; VIB Center for Microbiology, VIB, Leuven, Belgium; EMBL, Heidelberg, Germany; Department of Cardiology, Rigshospitalet, University of Copenhagen, Copenhagen, Denmark; The Victor Phillip Dahdaleh Institute of Genomic Medicine at McGill University, Montreal, Canada; Department of Health Research Methods, Evidence, and Impact, McMaster University, Hamilton, Ontario, Canada; McMaster Institute for Research on Aging, McMaster University, Hamilton, Ontario, Canada; Department of Nephrology and Medical Intensive Care, Charité-Universitätsmedizin Berlin, Berlin, Germany; Department of Nephrology and Hypertension, University Hospital Erlangen, Friedrich-Alexander-Universität Erlangen-Nürnberg, Erlangen, Germany; Department of Clinical and Movement Neurosciences, UCL Queen Square Institute of Neurology, London, UK; Region Västra Götaland, Sahlgrenska University Hospital, Department of Clinical Physiology, Gothenburg, Sweden; Novo Nordisk Foundation Microbiome Health Initiative and the National Food Institute, Technical University of Denmark, Kongens Lyngby, Denmark; Center for Clinical Metabolic Research, Herlev-Gentofte University Hospital, Denmark; Department of Clinical Medicine, Faculty of Health and Medical Sciences, University of Copenhagen, Copenhagen, Denmark

## Abstract

Kidney disease is a rapidly growing global health concern, tightly linked to cardiovascular and metabolic disease within the recently defined cardiovascular-kidney-metabolic (CKM) syndrome. Routine clinical assessment of kidney function primarily relies on creatinine-based estimated glomerular filtration rates (eGFRcr) which can be affected by non-renal factors and declines only after substantial kidney damage. Using data from the MetaCardis study (N=1,723), we derived metabolomic kidney function gaps (mKFGs)-the difference between metabolome-predicted eGFRcr and measured eGFRcr. A physiology-guided feature-selection strategy - anchored on three cardiometabolic markers (haemoglobin, albumin and the Neutrophil-to-Lymphocyte Ratio) that associate with mKFGs but not with eGFRcr - yielded compact sex-stratified metabolite panels (40-98 out of 1,514 metabolites) that tracked cardiovascular-kidney-metabolic risk. Machine-Learning models revealed distinct metabolic signatures, including biomarkers of meat consumption and androgens, highlighting potential sex-specific pathways linked to CKM health. External validation in the general-population Canadian Longitudinal Study on Aging (N=9,958) and German Chronic Kidney Disease Study (N=5,113) cohorts revealed that mKFGs added incremental predictive value for future CKM events and all-cause mortality beyond eGFRcr and established risk equations.

Our findings open new avenues for personalised risk stratification for CKM disease progression and targeted interventions based on metabolically defined kidney health.

## Introduction

Chronic kidney disease (CKD) is a growing health burden^1^, largely driven by rising rates of type-2 diabetes (T2D) and hypertension^1,2^. Even mild declines in kidney function significantly increase the risk of Cardiovascular Disease (CVD) and all-cause as well as cardiovascular mortality^3^. This is greatly amplified in persons with T2D, where CKD can lead to a tenfold rise in cardiovascular death^4^. Renal, cardiac and vascular pathology can each bidirectionally affect the others within a broader cardiovascular-kidney-metabolic (CKM) syndrome framework that also encompasses metabolic disorders^5^. Integrated readouts of CKM health, rather than any single-organ measure, may therefore enable timely and effective risk stratification and intervention.

Kidney function is commonly assessed as the estimated Glomerular Filtration Rate, derived from serum creatinine, age and sex using the CKD-EPI formula (eGFRcr, thereafter)^6^. While eGFRcr is inexpensive, routinely measured in clinical practice, sensitive to cardiometabolic risk factors such as insulin resistance^7^, and can reflect the risk of future CVD and CKD^8^, it has limitations: serum creatinine levels are influenced by muscle mass and diet^9^, eGFRcr underestimates kidney function in women^10^ and it is a lagging indicator of renal decline, becoming abnormal only after considerable kidney damage has occurred^9^. These limitations underscore the need for earlier, more sensitive markers that capture CKM risk beyond eGFRcr alone.

Recent studies coupling Machine Learning (ML) with multi-omics data (proteomics, metabolomics, metagenomics, organ imaging) have predicted biological organismal or organ-specific age^11–14^, cardiorespiratory fitness^15^, disease trajectories^16,17^ or BMI^18,19^. By integrating multiple signals related to health status not captured by their conventional equivalents, these omics-derived scores can complement existing biomarkers and provide deeper insights into metabolic health and disease trajectories, including response to intervention^18,20^.

The circulating metabolome can capture multi-organ human pathophysiology, with two-thirds of significant metabolite-disease associations spanning two or more noncommunicable diseases^21^.

Here, we hypothesise that a metabolome-derived score anchored on eGFRcr, by integrating multi-organ cues across cardiovascular-kidney-metabolic axes, could complement eGFRcr and established clinical risk scores^22–24^ in predicting future cardiovascular-kidney-metabolic events in both general and CKD populations and identify candidate metabolic pathways for follow-up mechanistic and clinical studies.

Using ML and data from 16,794 participants from three independent studies, *i.e.*, MetaCardis^25–28^, Canadian Longitudinal Study on Aging^29,30^ (CLSA), German Chronic Kidney Disease study^31^ (GCKD), we demonstrate that circulating metabolites track personalised risk of kidney function decline or CVD events beyond eGFRcr and established clinical risk scores (Fig.1A). We also uncover sex-specific metabolome signatures predictive of kidney function, offering insights into purported mechanistic differences in CKM physiology between sexes. To expedite translation, we use a physiology-guided feature selection strategy to devise generalisable and sex-specific metabolomic panels (40-98 serum metabolites) that outperform models using all available metabolites. Our work suggests application of ML algorithms and serum metabolites for monitoring risk of future cardiovascular-kidney-metabolic events and informs mechanistic and clinical studies to uncover the role of metabolic perturbations in CKM health.

**Figure 1.**
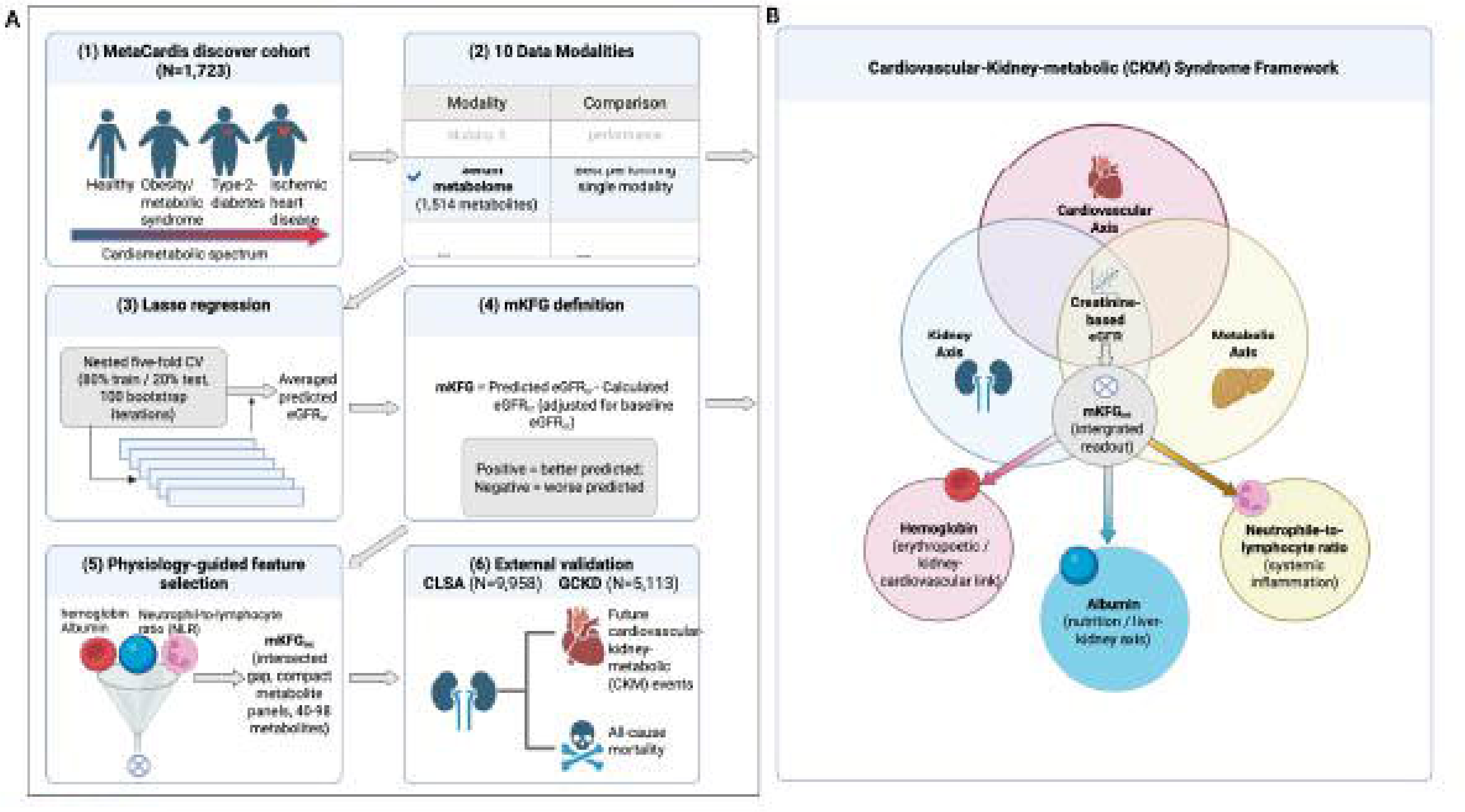
Study design and the cardiovascular-kidney-metabolic (CKM) framework for metabolome-based kidney function gaps (mKFG). (**A**) Analysis strategy. (1) The MetaCardis discovery cohort (n = 1,723) spans the cardiometabolic spectrum from healthy individuals to obesity/metabolic syndrome, type 2 diabetes and ischaemic heart disease. (2) Of ten data modalities compared, the serum metabolome (1,514 metabolites) was the best single predictor of creatinine-based eGFR (eGFRcr). (3) A nested 5-fold cross-validation (80% train / 20% test, 100 bootstrap iterations) was used to predict eGFRcr via Lasso, giving an averaged predicted eGFRcr per participant. (4) The metabolomic kidney function gap is defined as mKFG = predicted minus calculated eGFRcr, residualised on baseline eGFRcr (positive, better-than-expected; negative, worse-than-expected predicted kidney function). (5) Physiology-guided feature selection, anchored on haemoglobin, albumin and the neutrophil-to-lymphocyte ratio (NLR), yields the intersected gap (mKFGint) from compact metabolite panels (40 to 98 metabolites). (6) Associations with cardiometabolic markers were examined in MetaCardis, and model performance was validated in the Canadian Longitudinal Study on Aging (CLSA; N = 9,958) and the German chronic kidney disease (GCKD) study (N = 5,113) to predict myocardial infarction, stroke, all-cause mortality, cardiovascular mortality, kidney failure, acute kidney injury, and 3-/4-point major adverse cardiovascular events in GCKD, as well as kidney disease incidence, incident myocardial infarction, incident type 2 diabetes, cerebrovascular disease incidence, and unplanned hospitalization in CLSA. (**B**) Creatinine-based eGFR (eGFRcr) sits at the intersection of the cardiovascular, kidney and metabolic axes. Physiology-guided optimisation tunes mKFGint toward three cross-axis CKM markers, haemoglobin (erythropoietic and kidney-cardiovascular link), albumin (nutrition and liver-kidney axis) and the neutrophil-to-lymphocyte ratio (NLR; systemic inflammation), so that mKFGint serves as an integrated readout that is more sensitive than eGFRcr to CKM-syndrome biology and to downstream CKM events. Created with BioRender.

## Results

In the cross-sectional MetaCardis^25–28^ cohort (N=1,723; Suppl.Table.1), we used Lasso regression to predict eGFRcr from circulating metabolites and nine other data modalities (Fig.2A). We reasoned that models trained across the cardiometabolic disease spectrum would better capture cardiovascular-kidney-metabolic health in the presence of co-morbidities and generalise across the range of cardiometabolic and kidney disease burden.

**Figure 2.**
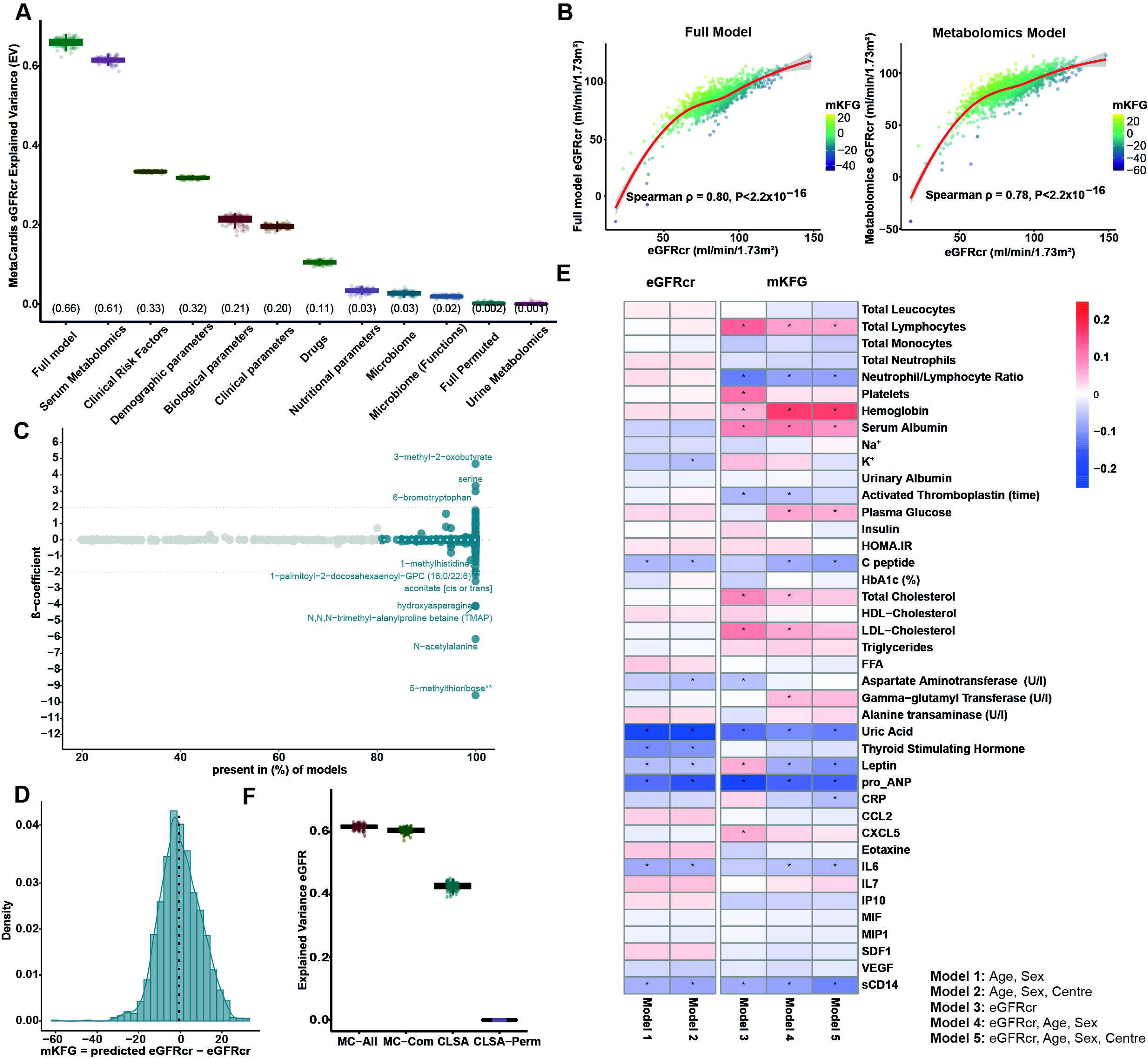
Serum metabolomics is the best eGFRcr predictor in the MetaCardis cohort. (**A**) Explained Variance (EV) of eGFR in MetaCardis (N=1,723) using lasso regression with input from the different data modalities shown on the x-axis. The EV for each category is shown above its label. (**B**) Scatterplots show the association between predicted and calculated eGFR for the full model (left) and the metabolomics-only model (right). Each participant is coloured by their mKFG (range shown). The red line indicates the Locally estimated scatterplot smoothing (LOESS) fit. (**C**) Averaged metabolite β-coefficients from 100 Lasso models trained with Metabolomics in the entire MetaCardis cohort. Metabolites present in more than 80% of models are coloured in blue. (**D**) Distribution of mKFGs in MetaCardis participants. (**E**) Partial Spearman correlations between baseline eGFR or mKFGs and bioclinical variables in MetaCardis. Correlations were adjusted for: Model 1: age and sex, Model 2: age, sex and country of recruitment, Model 3: baseline eGFR, Model 4: baseline eGFR, age and sex, Model 5: baseline eGFR, age, sex and country of recruitment. Positive correlations are shown in red, and negative correlations in Blue, * denotes significant correlations (FDR <0.1). (**F**) Validation in CLSA. Explained variance of eGFR (z-scored) in MetaCardis with all available Metabolites (1,514 metabolites, MC-all), explained variance from metabolites common in MetaCardis and CLSA (MC-Com), explained variance in CLSA with models trained in MetaCardis with common metabolites (CLSA) and in CLSA when the metabolite values for each CLSA participant were randomly permuted (CLSA-Perm). In B, E, F and H, the boxplots show median and interquartile ranges (IQR) with whiskers specifying ±1.5*IQR. Abbreviations: eGFR: estimated glomerular filtration rate; FFA: free fatty acids; BMIS: metabolic syndrome or risk factors; T2D: type 2 diabetes; IHD: ischaemic heart disease; CCL2: C-C motif chemokine ligand 2; CRP: C-reactive protein; CXCL5: C-X-C motif chemokine ligand 5; HDL: high-density lipoprotein; LDL: low-density lipoprotein; IL6: interleukin-6; IL7: interleukin-7; IP-10: interferon gamma-induced protein 10; MIF: macrophage migration inhibitory factor; MIP1: macrophage inflammatory protein 1; K⁺: potassium; Pro-ANP: pro-atrial natriuretic peptide; SDF1: stromal cell-derived factor 1; Na⁺: sodium; VEGF: vascular endothelial growth factor; sCD14: soluble cluster of differentiation 14.

### Serum metabolites outperform other variable groups in predicting eGFRcr in the European MetaCardis study

We took an agnostic approach to determine which of the ten data modalities (including clinical, lifestyle and dietary information, biochemical variables, serum and urine metabolomes and gut metagenomes) collected in MetaCardis participants (Suppl.Data.1) better predicts eGFRcr (Fig.2A).

The serum metabolome (1,514 metabolites, excluding creatinine) predicted eGFRcr similarly to the full model with averaged explained variance of 0.61 (Spearman ρ=0.78) *versus* 0.66 for the full model (Spearman ρ=0.80) and 0.33 for traditional clinical risk factors (smoking, age, sex, systolic blood pressure, glycated haemoglobin and serum lipoprotein levels)^27,28^ (Fig.2A&B). To confirm the appropriateness of Lasso to predict eGFRcr in our population we demonstrated significant linear associations between input metabolites and eGFR (Suppl.Fig.2A) consistent with Lasso assumptions^32^. Moreover, an alternative tree-based ML algorithm (boosted-trees^28,33^“*Methods*”) achieved modest improvements in eGFR explained variance with high overlap (12/20) in metabolites driving predictions with Lasso (Suppl.Fig.2B-D; Suppl.Table.2). Among the ten data modalities compared, the urine metabolome, profiled by ^1^H-NMR spectroscopy was the weakest predictor of eGFRcr (Fig.2A).

To pinpoint influential metabolites driving eGFR predictions, we averaged β-coefficients from Lasso models for each metabolite (Fig.2C). Higher levels of the reno-protective^34^ α-ketoacid 3-methyl-2-oxobutyrate, an intermediate of valine catabolism, the amino acid serine (possibly reflecting tubular re-absorption^35^) and 6-bromotryptophan, linked to diminished CKD progression^36^ drove eGFR predictions higher. Conversely, 5-methylthioribose an intermediary of the methionine salvage pathway^37,38^, the marker of N-terminal protein acetylation N-acetylalanine^39^ associated with incident CVD^40^, N,N,N-trimethyl-alanylproline betaine (TMAP) a metabolite of unknown function associated with lower eGFR^41^, and hydroxyasparagine a proposed marker of fibrosis^42^, were the top metabolites driving lower eGFR predictions.

Building on a conceptual framework developed for aging clocks^11–13^, we defined personalised metabolomic Kidney Function Gaps (mKFGs) by subtracting from predicted eGFRcr values the corresponding calculated eGFRcr. Positive mKFG values denote higher predicted, whilst negative values imply lower predicted eGFR compared to the calculated eGFR. mKFGs, adjusted for baseline eGFR, were distributed around a mean of -0.1 ml/min/1.73m^2^ (Fig.2D) indicating a broadly equal number of participants with predicted improved or diminished eGFR. mKFGs correlated inversely but much less strongly with age compared to eGFRcr (ρ=-0.12, P=6.8×10^-7^ *versus* ρ=-0.53, P<2.2×10^-16^, respectively; Suppl.Fig.1C-D) indicating that mKFGs reflect the aging process to a lesser degree.

Collectively, our analyses indicate that the circulating metabolome captures most phenotypic information related to eGFRcr.

### Sex-specific metabolic signatures of eGFRcr in men and women

As there is a higher CKD incidence in women^43^ but higher CVD mortality in men with CKD^43^, we next tested whether differences in CKM outcomes between sexes are reflected in the metabolome by training sex-stratified serum metabolomics models to predict eGFRcr. Models performed better in men (N=875) than those in women (N=848) in predicting eGFR (explained variance 0.63 *versus* 0.52, Spearman ρ=0.79 *versus* 0.73; Suppl.Figs.3A-C). Discrete metabolic signatures drove predictions between men and women (Suppl.Figs.3D-E). Of 55 metabolites common to both sexes (present in 80% of the models), 54 shared the direction of their β-coefficient, yet only 7 of the top 20 by absolute β-coefficient overlapped. Common metabolites included the microbial metabolite γ-butyrobetaine^44^ implicated in kidney fibrosis^28^, TMAP, 5-methylthioribose and 3-methyl-2-oxobutyrate, as in models trained with all participants. The thyroid hormone thyroxine associated with higher predicted eGFRcr only in women, suggesting possible divergent mechanisms underlying inter-organ crosstalk related to cardiovascular-kidney-metabolic health between sexes.

### Metabolomic kidney function gaps (mKFGs) reflect cardiovascular-kidney-metabolic health better than eGFRcr

To determine whether mKFGs inform on CKM health status, we tested their associations with biochemical variables and contrasted them to eGFRcr. We adjusted mKFGs for age, sex, country of recruitment and the corresponding eGFRcr to determine their prognostic value independently of underlying eGFRcr. This approach also accounts for the well-documented tendency of ML predictions to revert to the mean of the predicted variable^45,46^, as previously applied for eGFRcr^11^.

Like eGFRcr, mKFGs inversely associated with levels of pro-Atrial Natriuretic Peptide (proANP), indicative of cardiac stress^47^, C-peptide, a marker of endogenous insulin secretion reflecting compensatory hyperinsulinaemia in insulin-resistant individuals^48^, the pro-inflammatory cytokine IL6 and the marker of monocyte activation, sCD14^49^ (Fig.2E). Unlike eGFRcr, mKFGs correlated directly with haemoglobin, regulated by the kidney^50^ and anticorrelated with incident diabetic nephropathy and biopsy-proven renal fibrosis^51^ or higher Kidney Failure (KF) risk^52^. mKFGs also associated with higher levels of albumin, also linked to risk of incident CKD and eGFRcr decline^53^, potentially through an underlying liver-kidney axis^54^ and anticorrelated with the Neutrophil-to-Lymphocyte ratio (NLR), a marker of systemic inflammation and cardiovascular mortality^55^. Similar patterns of association were observed with mKFGs from models trained in men (Suppl.Fig.4A) or women (Suppl.Fig.4B).

To enable external validation, we trained models predicting eGFR in MetaCardis with the 1,276 serum metabolites with available measurements in CLSA (common-model), an independent cohort representative of the Canadian adult population^29^ (Suppl.Table.3) as input. We next used these models to predict eGFR in CLSA and observed a median eGFRcr explained variance of 0.43 for all participants (Fig.2F), 0.54 for men (Suppl.Fig.4C) and 0.36 in women (Suppl.Fig.4C). This may reflect the documented divergence between eGFRcr measurements in women and actual kidney health^43^.

Collectively, these analyses indicate that metabolome-derived kidney gaps are associated with additional aspects of CKM health not correlated with eGFRcr.

### A physiology-guided feature selection strategy to optimise mKFGs improves their association with cardiovascular-kidney-metabolic health readouts

To extend the utility of mKFGs in reflecting CKM health status not captured by eGFRcr, we adopted a permutation-based approach recently applied to proteomic age clocks^11^. In the absence of classic biochemical readouts of kidney function (urinary Albumin Creatinine Ratio (uACR), urea, urate, cystatin C, calcium phosphate) in the non-CKD MetaCardis and CLSA cohorts, we sought metabolites that strengthen significant correlations of mKFGs with all the biochemical measures of CKM health not associated with eGFRcr (Fig.2E; haemoglobin, albumin or NLR; “*Methods*”). To that end, we randomly permuted each metabolite present in at least 20% of our eGFR-predicting Lasso models (436/1,276 serum metabolites common between MetaCardis and CLSA to facilitate validation) in the held-out test group, predicted eGFRcr, computed new mKFGs and correlated them with haemoglobin, albumin or NLR (Fig.3A). Metabolites that after permutation decreased the absolute correlation between mKFGs and the target variable were considered to strengthen their association. We discovered 66 serum metabolites, excluding drugs, that strengthened absolute associations of mKFGs with all three target measures (Fig.3B&C). We hypothesised that these metabolites (Intersected (int) panel thereafter) could simultaneously capture aspects of CKM physiology reflected by these biochemical measures but not by eGFRcr alone and trained Lasso algorithms with the intersected panel to predict eGFRcr and compute gaps (mKFG_int_).

**Figure 3.**
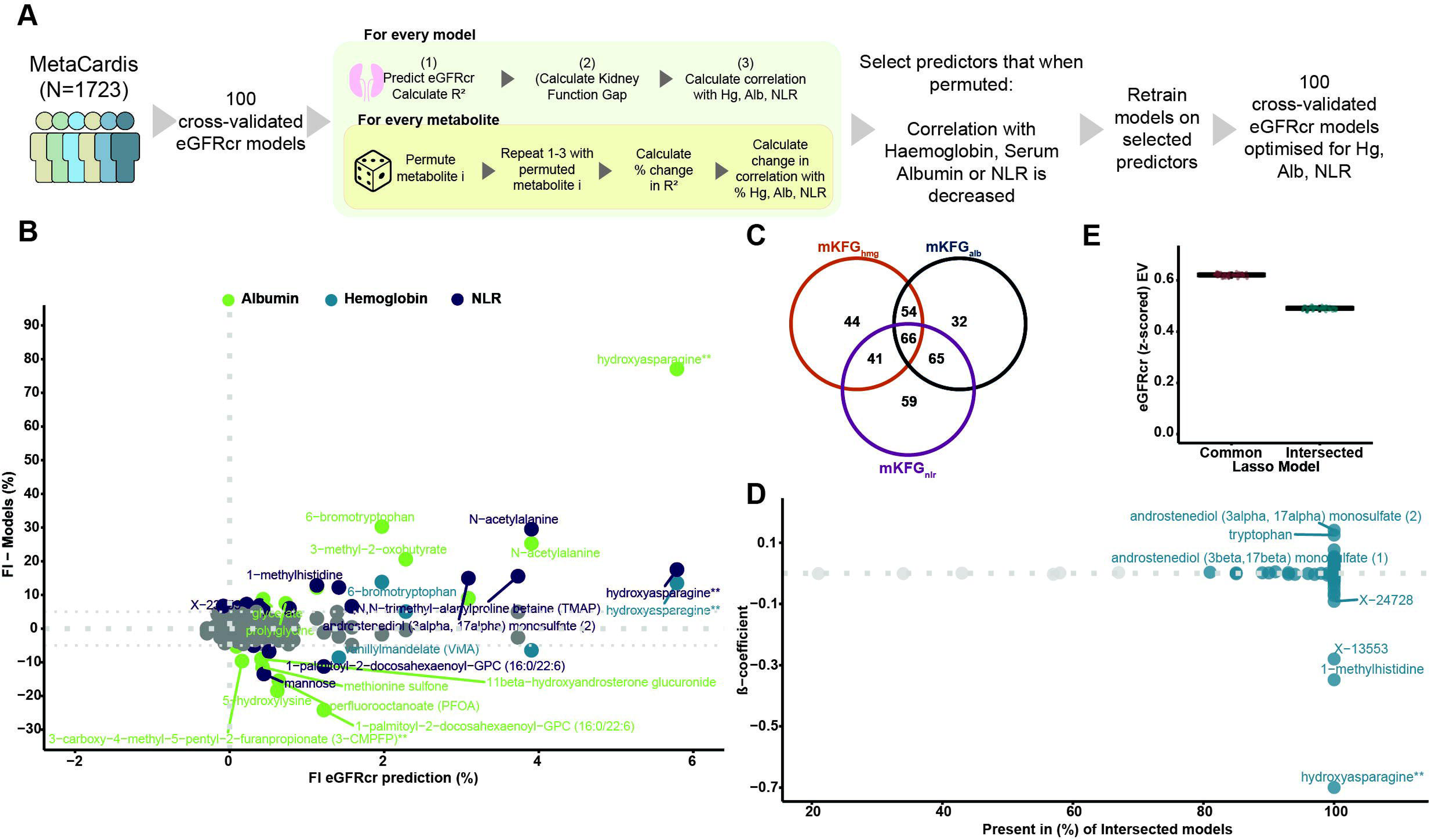
Physiology-guided optimisation of models predicting eGFRcr in the MetaCardis cohort. (**A**) Schematic of the optimisation flow: The metabolomic function kidney gap (mKFG) is calculated as described in Figure 1 and correlated with haemoglobin (Hb), albumin (Alb), and the Neutrophil-to-Lymphocyte Ratio (NLR). Each metabolite in the model is then permuted in the test set, and eGFR prediction is repeated to evaluate changes in explained variance (R^2^) and in the Spearman correlations with Hb, Alb, and NLR (adjusted for eGFRcr, age, sex and country of recruitment). Metabolites whose permutation reduces the correlation of gaps with these biochemical parameters are retained in separate panels for each variable (Hb, Alb, and NLR). (**B**) Most important metabolites (more than 7% difference in correlations with targeted variable after permutation) derived from the optimisation described in (A) for the targeted biochemical variables (serum albumin, Haemoglobin, NLR). X-axis: difference in eGFRcr R^2^after each given metabolite was randomly permuted. Y-axis: difference in correlation with each targeted biochemical variable following permutation in the test set. Positive values denote reduction in the strength of association after permutation in the test-set. FI: Feature Importance, each metabolite is coloured according to the targeted biochemical variable (**C**) Venn diagram of metabolites strengthening associations with each target bioclinical variable. 66 metabolites after drugs are excluded strengthen associations for all three targeted variables and are used to train the intersected model and derive mKFG_int_ (intersected mKFG). (**D**) Averaged metabolite β-coefficients from 100 Lasso models trained with the intersected metabolomics panel in all MetaCardis participants (n=1,723). Metabolites present in more than 80% of models are coloured in blue. (**E**) Explained Variance (EV) of eGFRcr in all MetaCardis participants with models trained with the 66 intersected metabolites. We also include predictions from the common metabolites (Com) model for comparison. Boxplots show median and interquartile ranges (IQR) with whiskers specifying ±1.5*IQR.

From the intersected panel, after extracting models β-coefficients, the proposed fibrosis biomarker hydroxyasparagine^42^ and 1-methylhistidine a metabolite reflecting dietary protein intake^56^ were the top metabolites driving lower eGFR predictions. Conversely the androgen androstenediol (3beta, 17beta) monosulfate together with the amino acid tryptophan were the top metabolites predicting higher eGFR (Fig.3D).

Intersected panel models, despite decreasing eGFRcr explained variance (Fig.3E) increased the associations of gaps with corresponding target biochemical variables (Fig.4A) compared to mKFG_all_, indicating that optimisation strengthened correlations with target biochemical measures. mKFG_int_ additionally associated with lower urine albumin, plasma Insulin and glycated haemoglobin (Fig.4B) and anti-correlated with standardised Odds Ratios (OR) of co-morbidities adversely affecting kidney health^2^ (i.e., hypertension, T2D, CAD) or atrial fibrillation (AF), more strongly that mKFG_all_ (Fig.4C) suggesting superior performance in reflecting CKM status than eGFRcr or mKFG_all_. Associations of mKFG_all_ and mKFG_int_ with comorbidities remained significant after inclusion of serum albumin, haemoglobin and NLR in the models (Fig.4D), demonstrating that intersected mKFGs capture information beyond their target biochemical variables.

**Figure 4.**
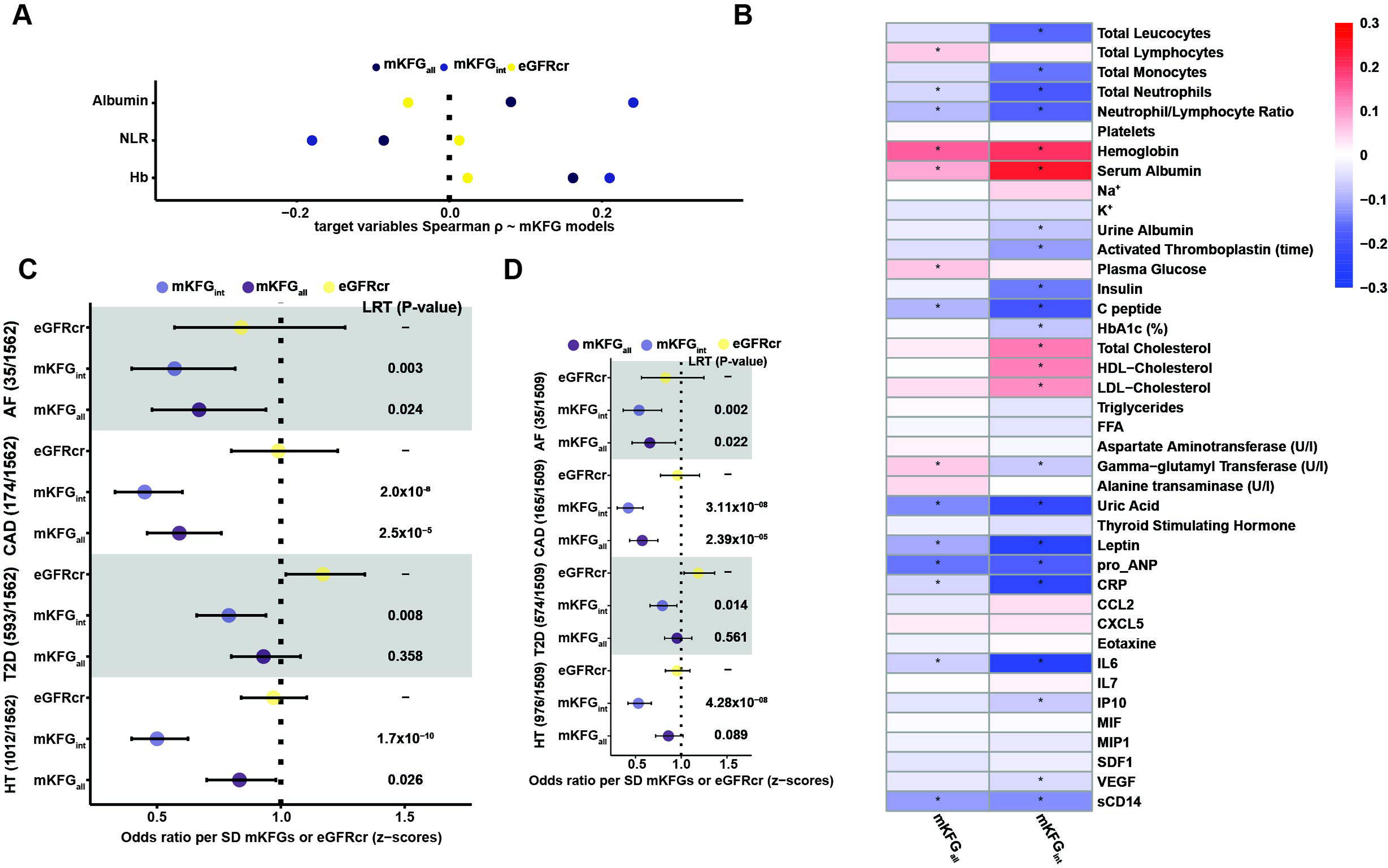
Metabolomic kidney function gaps reflect cardiovascular-kidney-metabolic status in MetaCardis. (**A**) Spearman correlations coefficients derived from partial correlations (adjusted for age, sex, country of recruitment and eGFR for mKFGs) are shown for eGFR, mKFG_int_ and kidney gaps generated from models trained with all common metabolites, in relation to the target biochemical variables (albumin, haemoglobin, and NLR). The colours indicate the corresponding mKFG or eGFR being correlated and x-axis the Spearman ρ. (**B**) Partial Spearman correlations between optimised mKFG_int_ (as defined in (F)) and gaps from models with all metabolites with biochemical variables in all MetaCardis participants. Correlations were adjusted for age, sex and country of recruitment. Positive correlations are shown in red, and negative correlations in Blue, * denotes significant correlations (FDR <0.1). (**C**) Forest plots illustrating standardised Odds ratio per SD of mKFGs or eGFR for hypertension (HT), T2D, coronary artery disease (CAD) and atrial fibrillation (AF) (sample size in parentheses) in all MetaCardis participants (N=1,723). P-values from the likelihood ratio test (LRT) comparing the baseline model (eGFR, age, sex, and country) with the same model plus mKFG are shown next to each bar. Bars indicate the 95% confidence interval. (**D**) Same analysis as in (C) with models also containing z-scores for serum albumin, serum haemoglobin and NLR. Abbreviations as in Figure 1.

Applying this strategy separately by sex identified 98 metabolites strengthening associations with all three biochemical measures in men (Suppl.Fig.5A-B) and 46 in women after excluding drugs (Suppl.Fig.6A-B). Although the intersected models reduced predicted eGFRcr explained variance (Suppl.Fig.5C, men; Suppl.Fig.6C, women) they associated more strongly with the target biochemical measures (Suppl.Fig.5D, men; Suppl.Fig.6D, women) and CKM readouts, including lower urine albumin in women, than mKFG_all_ or eGFRcr (Suppl.Fig.5F, men; Suppl.Fig.6F, women). mKFG_int_ anticorrelated with ORs of cardiometabolic disease prevalence in both sexes (Suppl.Fig.5G, men; Suppl.Fig.6G, women). Sex-specific drivers of eGFRcr predictions differed. In men, the inactive glucocorticoid cortisone^57^ and the anti-fibrotic pentose ribitol^58^ raised predicted eGFRcr; whilst TMAP^41^, hydroxyasparagine^42^ and propionylcarnitine, indicative of reduced lipid β-oxidation^59^ lowered it (Suppl.Fig.5E). In women the a-ketoacid 3-methyl-2-oxobutyrate^34^, serine and the androgen androstenediol (3alpha, 17alpha) monosulfate raised predicted eGFRcr, whereas the dietary protein intake biomarker^56^ 1-methylhistidine and gulonate (a kidney inositol catabolism intermediary^60^) lowered it (Suppl.Fig.6E). All MetaCardis association results are summarised in Suppl.Table.4.

Altogether, our analyses suggest that mKFGs, unlike eGFRcr, are sensitive to co-morbidities detrimental to kidney function and better associate with biochemical readouts of CKM health separately in men or women, and in all participants; with optimised panels of serum metabolites (46-98) improving on models containing all measured metabolites.

### mKFGs predict cardiovascular-kidney-metabolic status and disease trajectories in the general population-based Canadian CLSA cohort

To test whether mKFGs could assess CKM status and future disease trajectories in the CLSA cohort, we trained models in MetaCardis to predict eGFR in CLSA participants with available metabolomics (N=9,958).

Intersected models increased eGFRcr explained variance from 0.43 (for all metabolites; Spearman ρ=0.66) to 0.54 (Spearman ρ=0.74) (Fig.5A&B). As mKFGs in the MetaCardis cohort, mKFGs in CLSA, corrected for calculated eGFR distributed around 0 (Fig.5C). mKFG_int_ exhibited stronger absolute associations than mKFG_all_ or eGFR with albumin, haemoglobin and NLR (Fig.5D) demonstrating that physiology-guided optimisation in MetaCardis also strengthens associations in the CLSA cohort. mKFGs in CLSA, unlike eGFRcr, reflected cardiometabolic health correlating with lower systolic blood pressure, T2D duration, gynoid and android fat whilst associating with higher grip strength (a measure of reduced frailty^61^; Fig.5E) and lower ORs of co-morbidities (hypertension, T2D, MI) negatively impacting kidney function stronger than eGFRcr (Fig.5F). Interestingly, mKFGs correlated with increased serum lipid measures (total, high-density lipoprotein and low-density lipoprotein cholesterol) probably reflecting the medical management of lipids in individuals with prevalent cardiometabolic disease. Sex-stratified CLSA models (men N=4,882; women N=5,076) reproduced the generalisable model patterns for eGFRcr explained variance, associations with CKM readouts or cardiometabolic disease prevalence at recruitment (Suppl.Figs.7A-F, men; Suppl.Figs.8A-F, women). Moreover, sex-optimised intersected panels (98 metabolites in men, 46 metabolites in women) more strongly inversely associated with cardiometabolic disease prevalence than the generalised intersected panel-derived models (66 metabolites) (Suppl.Fig.7F, men; Suppl.Fig.8F, women), suggesting they better represent sex-specific CKM health.

**Figure 5.**
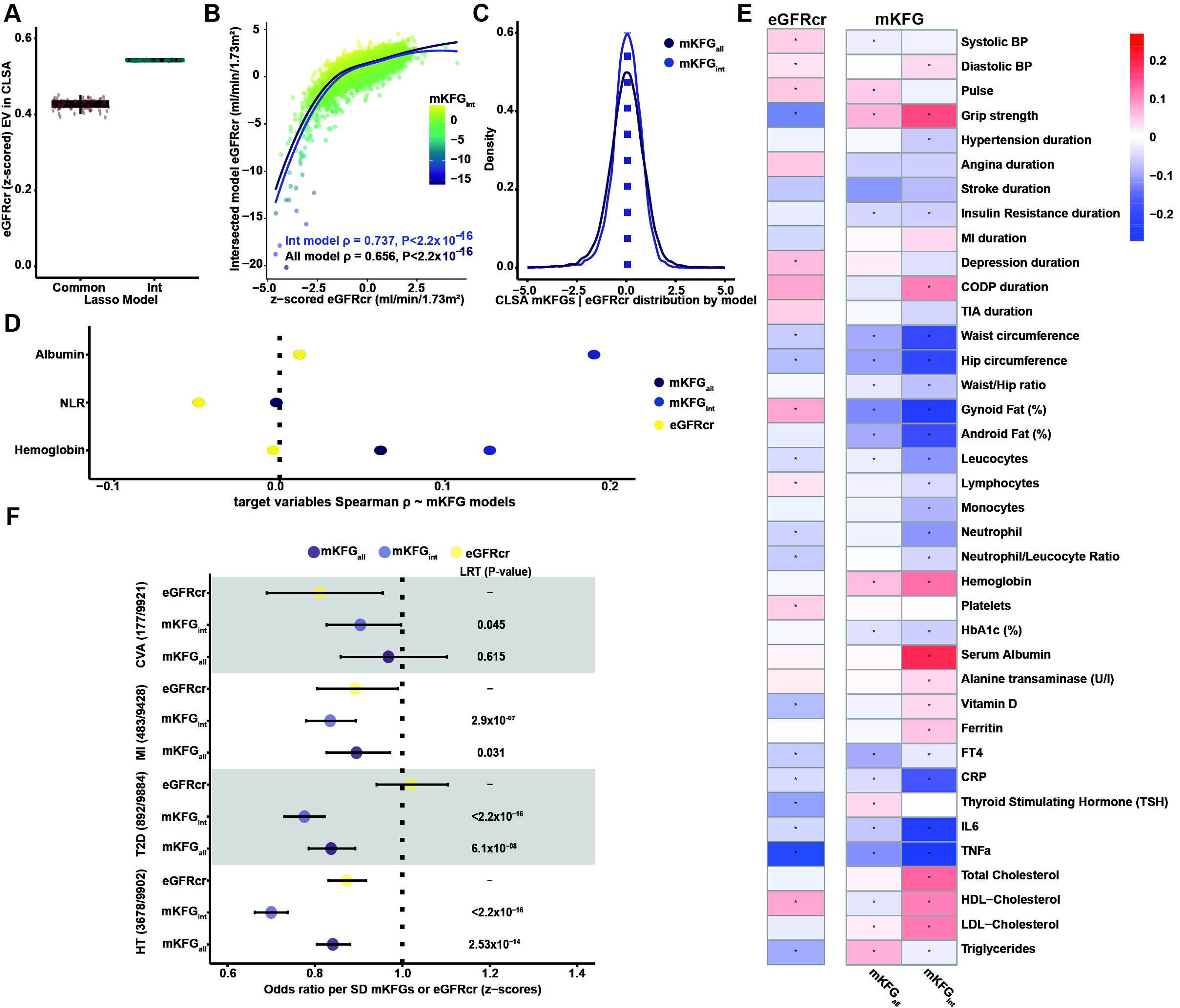
mKFGs reflect cardiovascular-kidney-metabolic status in the CLSA cohort. (**A**) Explained variance of eGFR in CLSA predicted with models trained in MetaCardis with the intersected panel of metabolites (66; Int) or all common metabolites (Common). The boxplots show median and interquartile ranges (IQR) with whiskers specifying ±1.5*IQR. (**B**) Scatterplot illustrating the association between predicted and calculated eGFR from intersected panel (mKFG_int_ in blue 66 metabolites) and the metabolomics model with all metabolites (in purple). The blue and purple lines denote the LOESS curves of corresponding model predictions. (**C**) Density plot showing the distribution of mKFG_all_ (purple) and mKFG_int_ (blue) in CLSA derived from eGFR predictions using models trained in MetaCardis. (**D**) Spearman correlations coefficients derived from partial correlations (adjusted for age, sex and also additionally eGFR for mKFGs) are shown for eGFR, and gaps derived from models trained with all intersecting metabolites between all variables (mKFG_int_) or all metabolites (mKFG_all_) with the targeted biochemical variables in CLSA (Albumin, NLR and Haemoglobin). The colours indicate the corresponding mKFG or eGFR being correlated. (**E**) Partial Spearman correlations between baseline eGFR (adjusted for age and sex) or mKFG_int_ or gaps from the intersected panel metabolites (adjusted for baseline eGFR, age and sex) with biochemical variables in all CLSA participants. Positive correlations are shown in red, and negative correlations in Blue, * denotes significant correlations (FDR <0.1). (**F**) Forest plots illustrating standardised Odds ratios for z-scored mKFGs or eGFR with prevalence at recruitment of hypertension (HT), T2D, Myocardial infarction (MI) or cerebrovascular disease (CVA) in CLSA. P-values from the likelihood ratio test (LRT) comparing the baseline model (eGFR, age, and sex) with the same model plus mKFG are shown next to each bar. LRTs were conducted in all CLSA participants with available metabolomics. Bars indicate the 95% confidence interval. Prevalence for each studied pathology in parentheses.

In those under the age of 74 (so FRS^24^ could be computed; N=6,638) with baseline and 1^st^ follow-up (3 years) and eGFRcr available, mKFGs inversely associated with eGFRcr rate of decline whilst eGFR exhibited positive and FRS no significant associations (Fig.6A) with similar trends observed for sex-stratified analyses (Suppl.Fig.9A, men; Suppl.Fig.9B, women). We also tested associations with rapid kidney function decline^62^, which also independently associated with all-cause mortality in a non-CKD population^63^. mKFGs significantly anticorrelated with ORs of rapid eGFRcr decline, which survived FRS inclusion, and improved performance when included in models, whilst eGFRcr associated with increased ORs of rapid kidney decline (Fig.6B). mKFGs additionally associated with lower ORs of incident CKD (Fig.6C) with once more FRS inclusion to models not improving performance. We additionally investigated whether mKFGs can predict incident T2D, future CV events (MI, CVA) or unplanned hospitalization (predictive of frailty and mortality^64^) in the CLSA cohort at the 3-year follow-up. mKFG, unlike eGFRcr, associated with lower ORs of unplanned hospitalizations and of incident MI, improving on baseline models whilst exhibiting a trend towards lower incident T2D (Fig.6D). Altogether, our work suggests that mKFGs associate with aspects of CKM health independently of conventional cardiovascular risk factors.

**Figure 6.**
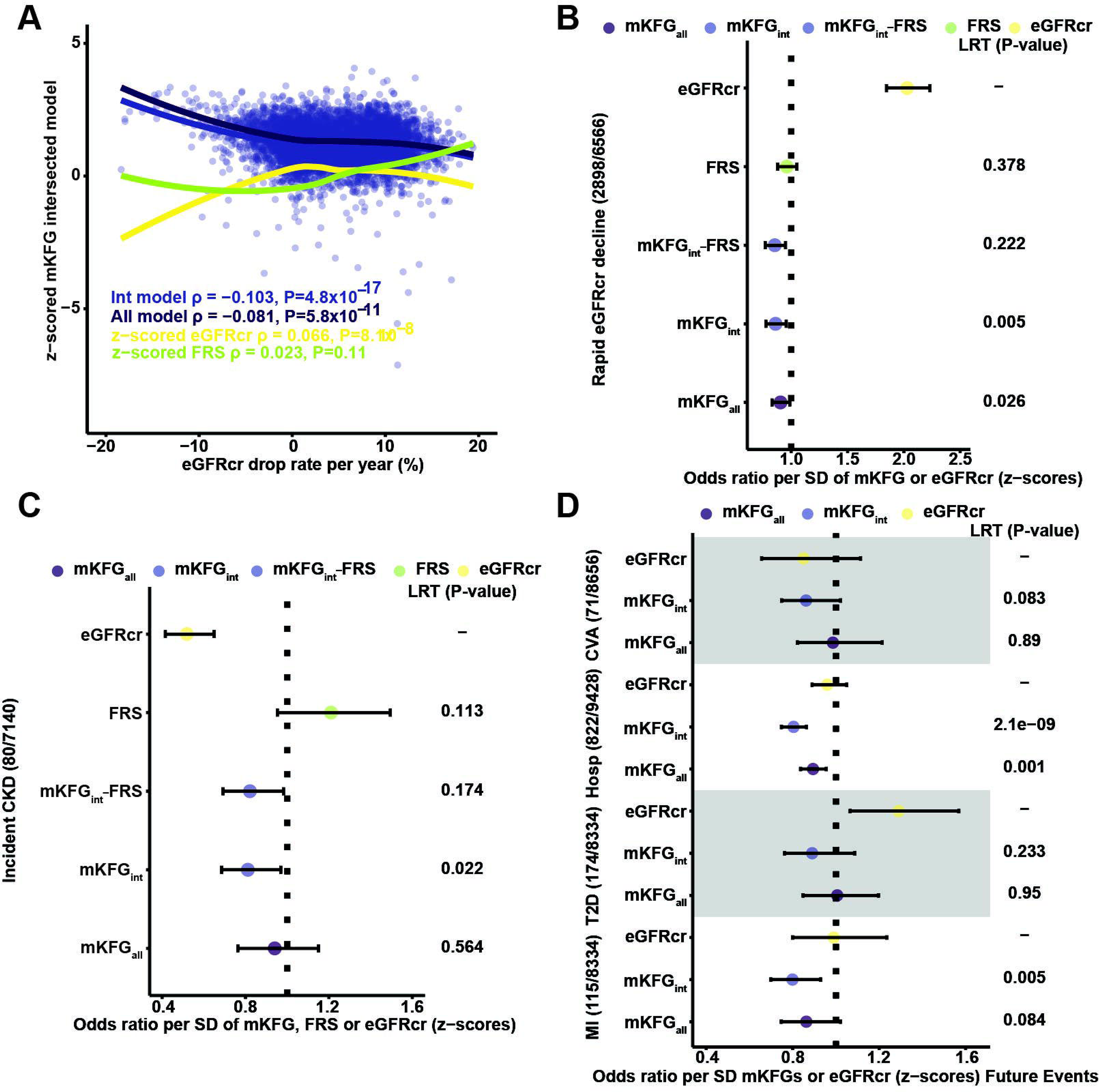
Metabolomic kidney gaps predict future cardiovascular-kidney-metabolic events in the CLSA cohort. (**A**) Scatterplot of eGFR drop rate (%) in CLSA participants with baseline and 1^st^ follow-up (3 years) calculated eGFR measurements *versus* z-scored kidney gap computed at baseline with the intersected model. Lines denote LOESS curves with mKFG calculated on all metabolites (All model in purple), intersecting metabolites (Int in blue), Framingham score (FRS in green) and eGFR (in yellow). (**B**) Forest plots of standardised odds ratios (ORs) per standard deviation for the metabolomic function kidney gaps (mKFGs), the Framingham Risk Score (FRS), or eGFR in predicting rapid kidney decline (≥5 mL/min/1.73m^2^ drop in eGFR per year, or ≥15 over three years). Baseline models included eGFR, age, and sex; p-values from likelihood ratio tests (LRT) comparing these baseline models to models additionally including each mKFG are shown beside the bars. Including FRS in the intersected model (mKFG_int__FRS) did not improve prediction (LRT = 0.25). The prevalence of rapid kidney decline in the CLSA is shown in parentheses, and bars/whiskers represent 95% confidence intervals. (**C**) Forest plot of standardised ORs per standard deviation for mKFG, FRS or eGFR in predicting incident CKD after 3-years. Baseline models included eGFR, age and sex; LRT p-values comparing baseline models to those additionally including each mKFG in all CLSA participants are reported. Adding FRS to the model with the mKFG calculated using intersecting metabolites (mKFG_int__FRS) did not significantly improve prediction (LRT = 0.18). The prevalence of incident CKD is shown in parentheses, and bars/whiskers denote 95% confidence intervals. (**D**) forest plot of standardised Odds ratio for z-scored mKFGs or eGFR with incident Myocardial infarction (MI), T2D, cerebrovascular disease (CVA) in the 3-year follow-up or unplanned hospitalization in the last year in CLSA participants. LRT were performed between baseline models including eGFR, age and sex or models including indicated mKFGs, baseline eGFR age and sex in all CLSA participants with p-values denoted next to the bars. Event rate is shown in parentheses, and bars/whiskers indicate 95% confidence intervals.

We next tested the relationship between mKFGs and all-cause mortality, the only outcome with time-to-event data in the CLSA cohort (373 deaths/9,205 participants at 6.5-year follow-up). CLSA participants in the high, medium and low tertiles of mKFG_all_ or mKFG_int_ after baseline calculated eGFRcr regression, exhibited divergent all-cause mortality risk after 6.5 years of follow-up (Fig.7A&B). The highest cumulative incidence rate was observed in the “Low” mKFG_all_ and mKFG_int_ tertiles, suggestive of worse predicted eGFRcr (5.7% and 6.6% respectively) whilst those at the “Highest” mKFG_all_ and mKFG_int_tertiles (indicative of better predicted eGFRcr) had 2-to 3-fold lower cumulative incidence rates (2.9% and 1.8% respectively). In Cox models also including eGFRcr, sex and age (Model 2; Fig.7C) each SD of mKFG_all_ or mKFG_int_ conferred a near 20% lower Hazard Ratio (HR) of all-cause mortality and improved discrimination (C-statistic; ΔC = +0.004 and +0.016, respectively), model fit (Likelihood Ratio Test; LRTs, P<0.05), “goodness-of-fit” of the data (informativeness^65^; by 121.2% and 154.4%, respectively), and patient reclassification (NRI: Net Reclassification Index^15^; by 8.4% and 15.6%, respectively). The mKFG association with lower all-cause mortality HR held true after adding FRS to covariates improving NRI by 27.2% (Model 3). This more stringent model (“*Methods*”) consistently improved classification of individuals at higher risk of death for a wide range of False Positive Rates (Fig.7D). Models including mKFG_int_ were well-calibrated, with bootstrap-corrected calibration slopes comparable to the baseline models, indicating that the gains in discrimination and reclassification reflect added prognostic information rather than differential calibration (Suppl.Fig.10). As a sensitivity test, applying the identical optimisation pipeline to 100 random triplets drawn from a pool of cardiovascular- and metabolic-disease-relevant biochemistry uncorrelated with both eGFRcr and mKFG_all_ (“*Methods”*) did not recapitulate mKFG_int_’s broader CKM performance in CLSA (Suppl.Fig.11A-B). Moreover, mKFG_int_’s associations with CLSA cardiorenal outcomes broadly survived direct adjustment for Hb, albumin and NLR (Suppl.Fig.12A-F) demonstrating that mKFG_int_ is not merely a proxy for these variables. Additionally, to confirm that mKFG_int_ is not simply a metabolome-based filtration estimate, we adjusted the CLSA models for a metabolome-derived eGFR computed from the validated metabolite GFR-estimating equation of Coresh *et al*^66^. mKFG remained an independent predictor of future CKM events after this adjustment (Suppl.Table.5). As a further sensitivity test we confirmed that mKFG_int_ retained positive incremental discrimination over the eGFRcr, age and sex baseline within strata of age, sex, diabetes status and baseline kidney function, with its prognostic effect strongest where creatinine-based eGFR is least informative (HR 0.59 per SD for eGFR ≥60 versus 0.82 for eGFR <60; mKFG_int_ × eGFR interaction P=1.6×10⁻⁵; Suppl.Table.6). Finally, to confirm the reproducibility of the top metabolite selections, we trained Lasso models to predict eGFRcr *de novo*in CLSA using the common (1,276) metabolites. 35 of the 40 leading MetaCardis predictors (ranked by absolute β-coefficient) were also robustly selected in CLSA (≥20% of models; Suppl. Fig. 13). All main CLSA association and mortality results are summarised in Suppl.Table.7.

**Figure 7.**
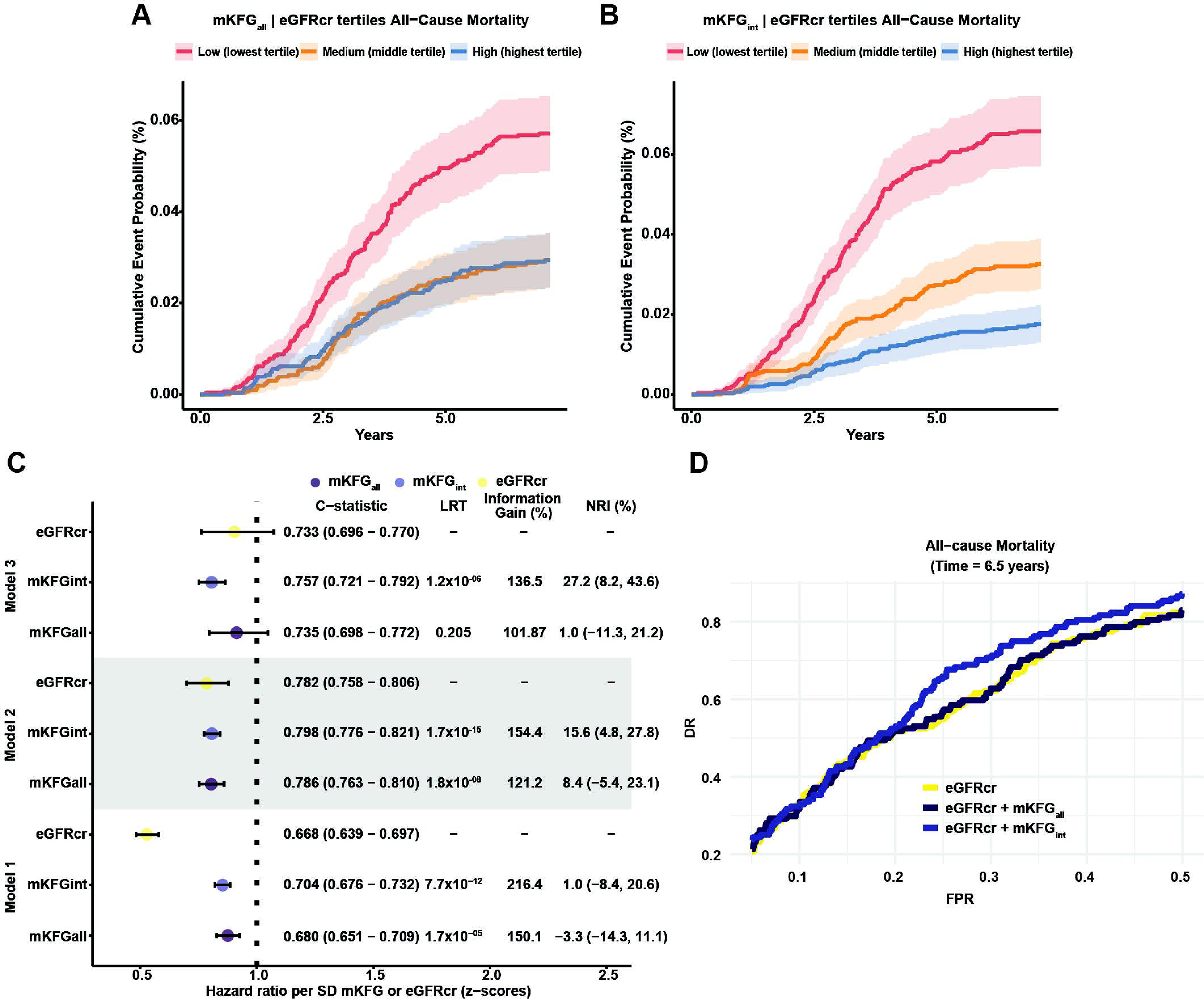
mKFGs improve all-cause mortality prediction in the CLSA cohort. **(A, B)** Cumulative all-cause mortality over 6.5 years among CLSA participants, grouped by mKFG tertiles using either (E) all common metabolites or (F) the intersecting 66-metabolite panel from MetaCardis (both adjusted for baseline eGFR). The “Low” tertile represents the worst predicted kidney function (lowest mKFG), and the “High” tertile represents the best predicted kidney function (highest mKFG). (**C**) Forest plot illustrating hazard ratios per SD of eGFR and mKFG computed from models using all common metabolites or the intersecting 66-metabolite panel for predicting all-cause mortality in CLSA over 6.5 years. Cox regression was applied with three baseline models: (1) baseline eGFR, (2) eGFR + age + sex, and (3) eGFR + age + sex + Framingham Risk Score (FRS). The mKFG (metabolomic function kidney gap) was added to each baseline model as indicated. The c-statistic (95% CI), likelihood ratio test (LRT) results, information gain (model fit improvement), and net reclassification index (NRI) are provided relative to the corresponding baseline model. (**D**) Detection Rates (DRs) across different False Positive Rate Thresholds (FPR) for All-cause mortality in CLSA derived from Cox regression models in (G) with the most stringent model (Model 3) containing kidney gaps from all or the intersected panel metabolites.

Collectively, we validated the utility of mKFGs derived from models trained in the MetaCardis cohort to detect future CKM outcomes in a geographically unrelated general population cohort. Crucially, mKFGs from models trained with the intersected panel of metabolites (mKFG_int_) consistently performed better than models trained with all metabolites, eGFRcr, or FRS, indicating their potential clinical utility.

### mKFGs are predictive of future cardiovascular-kidney-metabolic events in participants of the German Chronic Kidney Disease cohort with mild-to-moderate CKD

We next queried whether mKFGs can discriminate CKM disease trajectories in patients with predominately mild-to-moderate CKD of diverse aetiologies, participating in the GCKD study (N=5,113)^31^ (Suppl.Table.8). To this end, we trained models in the MetaCardis cohort using all serum metabolites (1,152) or 60 metabolites (out of 66) of the intersected panel common between the three cohorts to predict eGFRcr and computed mKFGs in the GCKD study. Models trained with all metabolites explained 62.1% of eGFR variance in GCKD (Spearman ρ=0.79) as opposed to 51.4% (Spearman ρ=0.72) for intersected panel models (Fig.8A-B). mKFGs inversely associated with proteinuria at recruitment more strongly than eGFRcr (Fig.8C&Suppl.Fig.14A).

**Figure 8.**
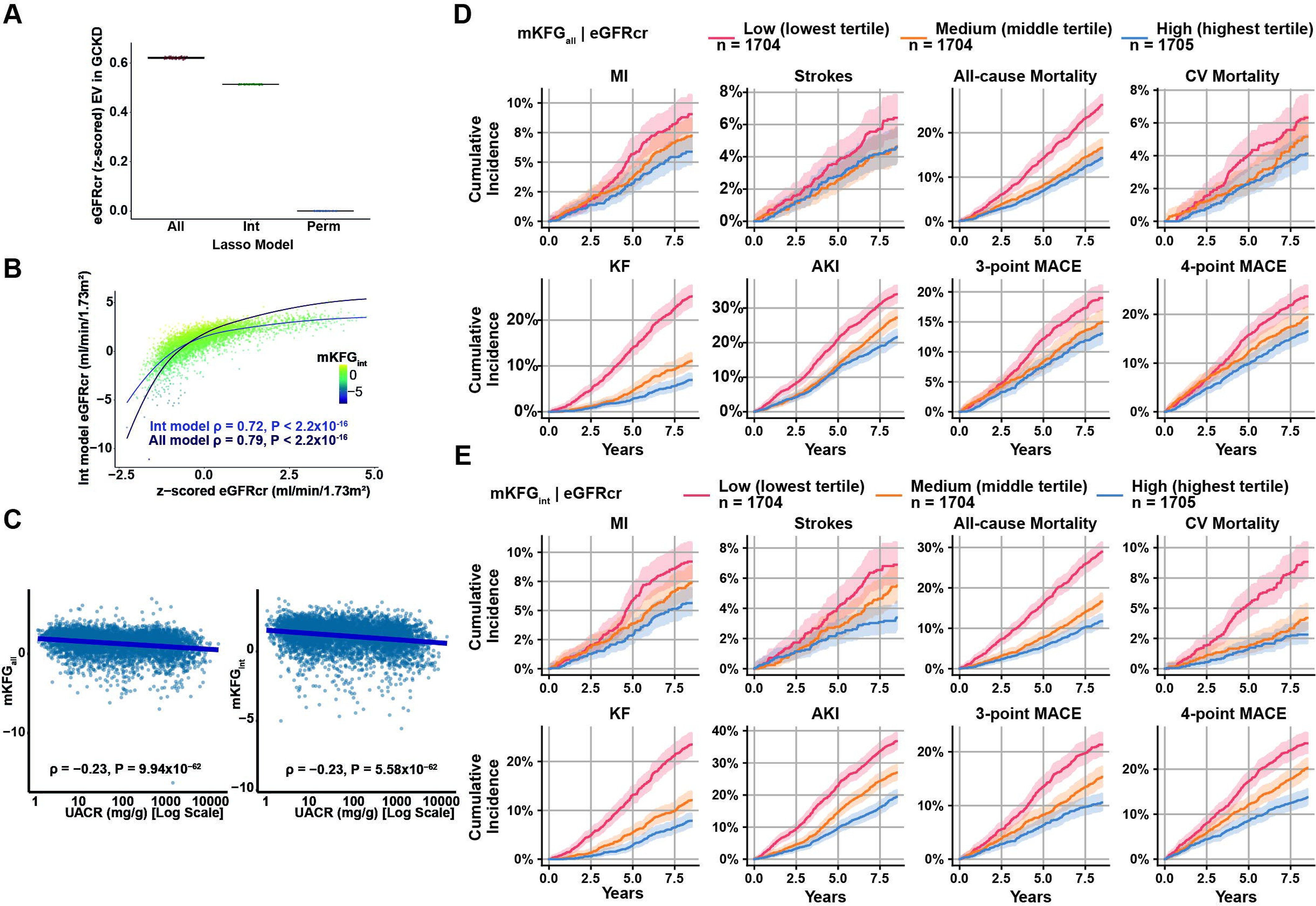
Metabolomic Kidney Function gaps reveal divergent cardiovascular-kidney-metabolic disease trajectories in the GCKD cohort. (**A**) Explained variance (EV) of eGFR in GCKD predicted with models trained in MetaCardis with the intersected metabolite panel (60), all common metabolites (1,152) available in all studied cohorts, or all common metabolites when eGFR values were randomly permuted (Perm). (**B**) Scatterplot illustrating the association between predicted and calculated eGFR from intersected model and the model containing all common metabolites. LOESS curves and Spearman correlations between predicted and calculated eGFR are provided for both models. Each participant is coloured according to the mKFG derived from the intersecting-metabolite model (range indicated). (**C**) Association between kidney gaps derived from eGFR predicted from all-common metabolites (left panel) or intersecting metabolites (right panel) with urinary Albumin-Creatinine ratio (uACR) in all GCKD participants. Spearman ρ from partial correlations corrected for eGFR, age and sex between mKFGs and uACR are shown in the insert. (**D**) Cumulative event probabilities for Myocardial infarction (MI), Stroke, All-cause Mortality, Cardiovascular (CV) Mortality, Kidney Failure (KF), Acute Kidney Injury (AKI), 3-and 4-point Major Adverse Cardiovascular Events (MACE) in GCKD participants over 8.5 years. The cohort was divided into tertiles based on residuals from a linear model regressing mKFGall against eGFR. The “low” tertile corresponds to the most negative residuals (i.e., worse predicted kidney function relative to calculated eGFR), whereas the “high” tertile corresponds to the most positive residuals (i.e., better predicted kidney function). (**E**) As in (D) for mKFG derived with the intersected metabolite panel (mKFG_int_, 60 metabolites).

We next leveraged the extensive longitudinal follow-up (up-to 8.5 years) of GCKD participants to query whether mKFGs computed on baseline metabolomics predict future CKM event incidence. We observed divergent cumulative incidence rate trajectories for all eight CKM events included in our analysis (3-point Major-Adverse Cardiovascular Events (MACE), 4-point MACE, acute kidney injury (AKI), all-cause mortality, CV mortality, KF, MI or stroke) in the “High” (higher predicted kidney function), median and “Low” (lower predicted kidney function) tertiles of mKFG_all_ and mKFG_int_ after regressing mKFGs on baseline calculated eGFR and using the residual. Cumulative incidence rates between the ’Low’ and ’High’ tertiles differed by 1.4-fold (stroke) to 3.6-fold (KF) for mKFG_all_, and were consistently better separated by the intersected panel, ranging from 1.6-fold (MI) to over 3-fold (CV mortality) for mKFG_int_ (Fig.8D-E).

To demonstrate that mKFGs do not merely represent improved GFR estimation, we subtracted from the model predictions eGFR calculated from serum cystatin C (eGFR_cys_), which better estimates true (measured) GFR and future CV risk^3^ but is used predominately for confirmatory clinical tests^62^ and plotted cumulative incidence rates as above. We still broadly observed considerable separation in cumulative incidence rates particularly for mKFG_int_ tertiles after eGFR_cys_ regression (Suppl.Fig.14B&C), consistent with our hypothesis that metabolome-derived mKFGs integrate signals broader than glomerular filtration relevant to kidney health.

Using Cox regression, we observed approximately 15-20% lower HR per SD of mKFG_all_ for all monitored outcomes except for stroke and CV mortality in the most stringent model (Model 4). These models included age, sex, uACR, and FRS, except the AKI model that included both FRS and the Kidney Failure Risk Equation (KFRE) score^22,23^ as a surrogate of baseline renal risk and the KF model substituted KFRE for FRS. Maximum improvement in model informativeness (“goodness-of-fit”) was observed for AKI (128%; Fig.9A-D, Suppl.Fig.14D-G, left panels and Suppl.Table.12) with an NRI of 20.6%. Models including mKFG_int_ performed significantly better than baseline models or models containing mKFG_all_, associating with reduced HRs by 15-33% for all outcomes in Model 4 exhibiting maximum improvement in informativeness by 167% for AKI with an 30.6% NRI (Fig.9A-D, left panels, Suppl.Fig.14D-G). When eGFRcr was replaced by combined creatinine-cystatin C eGFR (eGFRcr-cys^67^, a more accurate estimate of glomerular filtration, in the fully-adjusted GCKD models, mKFG_int_ retained significant incremental prognostic value for the broader CKM outcomes (Suppl.Table.9), indicating that it carries information beyond filtration. Using Model 4 to predict risk of all eight outcomes in GCKD participants for the 8.5-year follow-up period (“*Methods*”), we consistently obtained improved classification of individuals at high risk of event incidence for a wide range of False Positive Rates (10-40%) with models including mKFG_int_ outperforming those with all metabolites (Fig.9A-D and Suppl.Fig.14D-G, right panels). Across all eight outcomes, models with and without mKFG_int_ were comparably well-calibrated (bootstrap-corrected calibration slopes 0.96 to 0.99; all eight outcomes in corresponding Source data; all-cause mortality in Suppl.Fig.15). Sensitivity analyses confirmed that mKFG_int_ provided significant incremental discrimination over the fully-adjusted Model 4 baseline at administratively right-censored shorter follow-up horizons of 3 and 5 years for all eight outcomes (8.5 years being the primary horizon itself; Suppl.Table.10), and across prespecified subgroups defined by age, T2D status, KDIGO categories^68^, and CKD aetiology for all eight outcomes, with no systematic decay in effect size across horizons or strata (Suppl.Table.11). Sex-stratified analyses (sex-specific intersected panels: 91/98 metabolites in men, N=3,071; and 40/46 in women, N=2,042) reproduced these findings (Suppl.Fig.16-17), with the largest discrimination gains for AKI in men (ΔC +0.027, NRI 32%) and MI in women (ΔC +0.042, NRI 54%). All GCKD outcome results, for all participants and by sex, are summarised in Suppl.Table.12.

**Figure 9.**
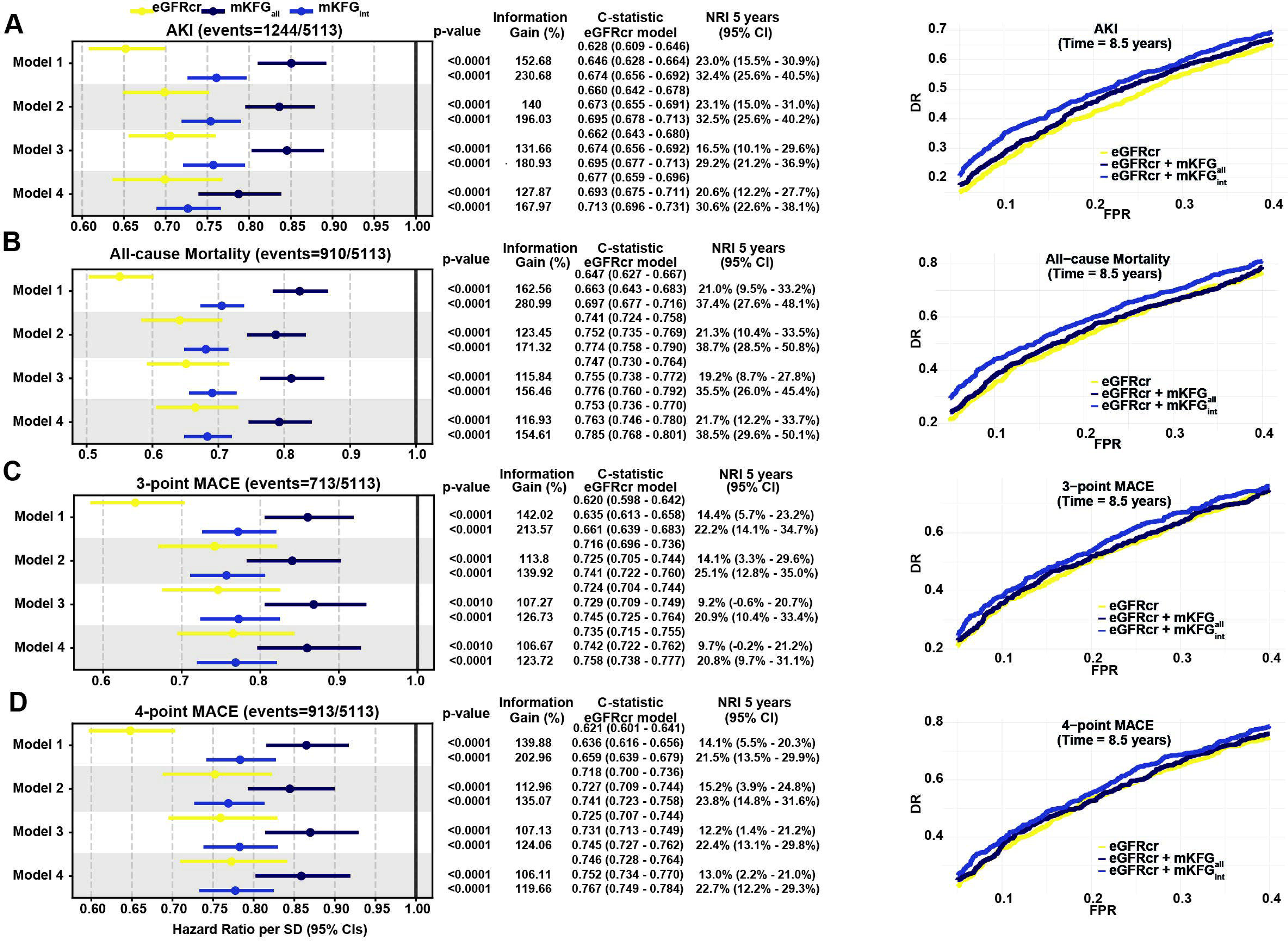
mKFGs improve prediction of cardiovascular-kidney-metabolic outcomes in the GCKD cohort. (**A-D**) (**left panels**) Forest plots of hazard ratios per SD of eGFR or mKFGs computed with models with all or the intersected panel metabolites for acute kidney injury (AKI), All-cause mortality, 3-point MACE and 4-point MACE in GCKD for the 8.5-years follow-up. Cox regression was applied with four baseline models: Model 1: baseline eGFR; Model 2: baseline eGFRcr, age and sex; Model 3: baseline eGFRcr, age, sex, urinary albumin creatinine ratio (uACR); Model 4: as model 3 with the Framingham risk score (FRS) for CV events (All-cause mortality, 3- and 4-point MACE) or with FRS and the 5-year Kidney Failure Risk Equation (KFRE) for AKI. Nested models additionally included each mKFG. The C-statistic (with 95% confidence intervals, in parentheses) and the Likelihood Ratio Test (LRT) P-value, the improvement in the Informativeness (“goodness-of-fit”) and Net Reclassification Index (NRI) are reported for each comparison between the baseline models and those containing mKFGs. (**Right panels**) Detection Rates (DRs) across different False Positive Rate Thresholds (FPR) for events in (A-D) derived from the corresponding Cox regression models with the most stringent model (Model 4) containing mKFG_all_ and mKFG_int_. In all shown forest plots the bar represents the 95% confidence interval.

We determined that, in addition to a cohort representative of the general population, mKFGs could detect future CKM outcomes in a mild-to-moderately severe CKD population with intersected metabolite panels consistently outperforming mKFGs from all metabolites and significantly improving performance of models containing eGFRcr, uACR or established scores of CKM risk in both sexes.

## Discussion

By harnessing the ability of ML algorithms to detect patterns in high dimensional data, we derived personalised metabolomic kidney function gaps (mKFGs) from ML-based eGFR predictions using circulating metabolites as input. mKFGs reliably distinguished baseline kidney and associated cardiometabolic health status and predicted future CKM disease trajectories across three independent cohorts of varying study designs and kidney disease burden totaling 16,794 participants. These cohorts were chosen to span the cardiovascular-kidney-metabolic disease spectrum, from a general population without overt CKD (CLSA) to an established CKD population at high cardiovascular risk (GCKD), allowing us to test whether a single baseline score predicts future CKM events across this range. Using a physiology-guided feature selection strategy, we optimised serum metabolite panels for men, women and all participants and captured CKM health status and risk more effectively than using all metabolites, creatinine-based eGFR and established risk scores only.

Despite extensive adjustment for multiple kidney health-related covariates (age, sex, eGFR, uACR), clinical risk scores (FRS, KFRE) and use of more precise method to estimate GFR (eGFR_cys_), models including mKFGs improved classification of individuals across CKM trajectories. Anchoring our metabolomic prediction on eGFRcr leverages an already CKM-relevant starting point rather than refining pure glomerular filtration. The physiology-guided optimisation toward haemoglobin, albumin and NLR - CKM-axis variables uncorrelated with eGFRcr in MetaCardis - extends this CKM-sensitivity beyond what eGFRcr alone captures. These findings imply that mKFGs are not merely refined estimates of GFR - that is, a risk-enriched metabolomic deviation from creatinine-based kidney filtration - but rather represent, by integrating metabolic signals, a multifaceted score of kidney and associated cardiometabolic health independently of existing markers. Specifically, mKFG_int_ inclusion, improved patient discrimination (from 0.012 (stroke) to 0.032 (all-cause mortality)) and reclassification (up to 38.5% for all-cause mortality) for cardiovascular outcomes comparable to those previously reported with proteome-based models^69,70^ underscoring the interplay between kidney and cardiovascular health. We additionally considerably improved patient discrimination for kidney-related outcomes (up to 0.036 C-statistic increase for all participants for AKI) leading to as much as a ∼31% improvement in patient classification. Further highlighting their utility, mKFGs derived from women-focused panels demonstrated effect sizes often exceeding those from models including all participants or men only, underscoring their potential value in predicting CKM outcomes among women, where there is a clinical need to more accurately assess kidney status^10^.

Our metabolite panels yielded candidate metabolites for future investigation and pointed to potential lifestyle or clinical interventions to mitigate CKM risks in a sex-specific manner. We identified in men, women and all participants the biomarker of meat consumption 1-methylhistidine^56^ as one of the top serum metabolites of reduced predicted kidney function, suggesting that moderating meat intake may confer kidney health benefits, in line with a recent study^71^. In women, androstenediols were predictive of better kidney function, aligning with previous research on the relevance of sex hormone status on female cardiovascular health^72^, warranting further dedicated kidney-centred research. Additionally, the α-keto acid 3-methyl-2-oxobutyrate associated with improved kidney function, suggesting that enhanced branched-chain amino acid catabolism could be renoprotective paralleling evidence from cardiac health^73^.

Despite omics-based risk scores often improving patient classification, their adoption in clinical practice is often hindered by their complexity and costs. The optimised metabolomic panels we derived, by reducing the metabolomic space required to monitor CKM health by up to 40-fold combined with improvements in automated sample preparation and metabolite annotation could pave the way to overcome some of these limitations. A comparable path, from broad discovery to a compact clinically deployed test, has recently been realised in liver disease, where a serum panel of 12 lipids combined with routine clinical measures can prioritise patients at risk of steatohepatitis^74,75^. Similarly, our work could serve as a paradigm for further development of selective metabolome-informed scores for clinical assessment of CKM health allowing for personalised risk stratification and early intervention. Early detection of individuals at higher CKM risk is particularly important, since new-generation anti-diabetic drugs (Glucagon-like peptide-1 receptor analogs and Sodium-glucose Transport Protein 2 inhibitors) which have documented renoprotective properties^76–78^ may help preserve kidney function if initiated early. Our findings could facilitate early detection of individuals at high risk of AKI, the most prevalent renal exacerbation event in patients with CKD, commonly leading to severe complications with high mortality^79^, allowing for targeted clinical vigilance and medication management.

Our study has some limitations. Our cohorts predominantly consisted of white participants albeit from varying geographical and environmental backgrounds (Europe and North America), questioning the generalisability of our findings across other ancestries. Additionally, we primarily relied on linear modelling to derive kidney function gaps and whilst a non-linear ML method in our training cohort did not confer appreciable benefits, non-linear ML could potentially further improve our approach particularly as computational power increases in the future. Untargeted metabolomics typically provide relative metabolite quantifications rather than absolute concentrations, preventing us from establishing universal reference concentrations. Devising targeted quantitative assays for our optimised metabolite panels, along with prospective validation, is an essential next step towards clinical application. Importantly, the relationships we report between mKFGs and cardiovascular-kidney-metabolic health are observational associations and do not, in themselves, establish causal or physiological mechanisms. The biological interpretations we offer for individual metabolites and pathways are therefore hypothesis-generating and warrant dedicated experimental validation. The lack of serial metabolomic measurements in our cohorts precluded direct assessment of whether mKFGs change over time in response to disease progression, lifestyle modification, or therapeutic intervention; this remains to be evaluated in cohorts or trials with serial metabolomic profiling. Additionally, urine metabolome data were not available across all three cohorts, so we could not test whether urinary metabolomics might outperform or complement the serum metabolome as a readout of CKM health. This remains a promising avenue for future work. Finally, like other omics-derived scores, our current panels rely on high-throughput metabolomics platforms that are not yet widely accessible and currently limit deployment in low-resource settings. Further reduction to compact, targeted assays, along with validation in ancestrally diverse cohorts will be needed to broaden accessibility.

To conclude, we demonstrate that leveraging ML with circulating metabolite profiles can refine estimates of CKM health and improve risk prediction of future CKM diseases beyond widely adopted clinical markers. Our work sets the stage for developing noninvasive tests to monitor CKM disease risk and informs future mechanistic and clinical studies aiming to preserve kidney function in both the general and the CKD population.

## Data availability statement

Raw shotgun sequencing data supporting the findings of this study are deposited in the European Nucleotide Archive under accession codes “<u>PRJEB37249</u>”, “<u>PRJEB38742</u>”, “<u>PRJEB41311</u>”, and “<u>PRJEB46098</u>”. In adherence to EU and national privacy laws, unrestricted access to patient phenotypic data cannot be provided for MetaCardis. Researchers, wishing to access individual phenotypic data would need to submit argued applications to the relevant National Data Protection Agencies. These are the Danish Data Protection Agency (https://www.datatilsynet.dk/english) for phenotypic data from study participants recruited in Denmark, the Federal Commissioner for Data Protection (https://www.bfdi.bund.de/EN/Home/home_node.html) for phenotypic data from study participants recruited in Germany and the Commission Nationale Informatique & Libertés (https://www.cnil.fr/en/home) for phenotypic data of study participants recruited in France. Application procedures are given on the outlined websites. If such permission is granted, phenotypic data will be then made available by the corresponding authors within 5 weeks. For CLSA, de-identified data are available from the Canadian Longitudinal Study on Aging (www.clsa-elcv.ca) for researchers who meet the criteria for access. For GCKD, public posting of individual-level participant data is not included in the informed patient consent form. To allow researchers to incorporate metabolomic kidney gaps in their analyses, we provide β-coefficients for all generalisable, sex-stratified and optimised metabolite panels so they can be computed in any cohort with metabolomics analyses done on the Metabolon platforms. Source Data are provided with this paper.

## Code availability statement

No new software or software library was developed in this study. Scripts implementing the key analyses will be deposited on Zenodo (DOI to be provided upon publication).

## Methods

### Cohorts Description

#### MetaCardis description, study design and bio-sample collection

MetaCardis (ClinicalTrials.gov Identifier: NCT02059538) is a cross-sectional study comprising individuals ranging from metabolically healthy to metabolic syndrome and/or obesity through T2D and IHD, aged 18–75-years old recruited in Denmark, France and Germany between 2013 and 2015. Patients under care of the participating hospitals meeting the inclusion criteria of the study were invited to enroll. Ethical approvals were obtained from the Ethics Committee CPP Ile-de France, the Ethical Committees of the Capital Region of Denmark (H-3-2013-145), and Ethics Committee at the Medical Faculty at the University of Leipzig (047-13-28012013). Study participants provided written informed consent, and the study was undertaken according to the Helsinki Declaration-II guidelines. Study design, recruitment and exclusion criteria have been extensively described^25–27,80^. Participants with severe kidney failure and/or on dialysis therapy, or with eGFR < 50 mL/min/1.73 m² (body surface area), were excluded, as specified in the registered MetaCardis protocol (ClinicalTrials.gov NCT02059538). Estimated GFR in the present analyses was calculated from serum creatinine using the CKD-EPI equation (eGFRcr). In the present study all participants with available eGFR measurements, serum metabolomics and microbiome composition were included in the analysis (N=1,723). In MetaCardis the following definitions for disease classification were used: hypertension, as defined by the American Heart Association^81^; obesity, as defined by the World Health Organization^82^, metabolic syndrome, as defined by the International Diabetes Federation^83^, T2D as defined by the American Diabetes Association^84^.

Within the MetaCardis ischaemic heart disease spectrum, cardiovascular phenotypes were defined as mutually exclusive categories per the registered protocol (ClinicalTrials.gov NCT02059538): acute coronary syndrome (ACS; STEMI or NSTE-ACS), chronic ischaemic heart disease (CIHD; stable coronary artery disease with LVEF ≥45% and no heart failure) and heart failure (HF; LVEF <45% or NYHA class II–IV, of ischaemic or non-ischaemic aetiology). CIHD and HF are thus distinguished by left-ventricular function and were not combined, as described^27^. Here, cardiometabolic disease is used as an umbrella term for all the above conditions. Biomaterial collection has been extensively described^26–28,80^. Blood samples were collected in the morning after overnight fasting, and fecal samples were collected at home by participants, frozen immediately and transferred to study centres on dry ice within 48eh. All samples were stored at −80°C until use. Clinical history, medication and phenotypic information were acquired as described^26,27^ with standardised procedures across centres. Food intake was reported through a customized Food Frequency Questionnaire (FFQ), validated according to established practices by three web-based dietary recalls^85^ for a subset of study participants (Ne=e324). Biochemical variables were measured in a single centre according to standard procedures^27^. Estimated GFR was calculated with the CKD-EPI formula without ethnicity adjustment^86^.

#### Canadian Longitudinal Study on Aging (CLSA) cohort description study design and bio-sample collection

CLSA is a Canadian cohort that follows 51,338 women and men recruited between the ages of 45–85eyears recruited from 2010-2015 and followed-up longitudinally every 3 years^87^. All participants provided written informed consent. Additional information for available bio-clinical data and collection, recruitment and exclusion criteria, ethics governance and oversight and data availability can be found here www.clsa-elcv.ca. In this study we used data up to Follow-up 1 (FU1; approximately 3 years since recruitment), except for all-cause mortality where data were available also for Follow-up 2 (FU2; approximately 6 years since recruitment), which was also the only outcome with time-to-event data in CLSA. Incident disease was defined as disease reported in FU1 but not at baseline. Rapid kidney function decline was defined as an annual eGFR decline >5 ml/min/1.73 m². (here >15 ml/min/1.73 m² over the 3-year FU1 interval). The use of CLSA data for the purpose of this study was further approved by the Imperial Research Ethics Committee under application number 21IC7388. Here we used CLSA participants, regardless of ethnicity with available eGFR measurement (with the CKD-EPI formula without ethnicity adjustment^86^) and serum metabolomics at recruitment (N=9,958).

#### German Chronic Kidney Disease (GCKD) cohort description study design and bio-sample collection

The German chronic kidney disease (GCKD) study is an ongoing prospective observational cohort that enrolled 5,217 patients with mainly CKD stage 3 between 2010 and 2012. Details of the study design have been published previously^31,88^. Participants were recruited under nephrological care based on an eGFR of 30–60 mL/min/1.73 m² or an eGFR >60 mL/min/1.73 m² with overt proteinuria, defined as a urinary albumin-to-creatinine ratio >300 mg/g or a urinary protein-to-creatinine ratio >500 mg/g. The study included adults aged 18–74 years. Detailed sociodemographic and clinical data were collected through standardised questionnaires and quality-controlled procedures. Biosamples collected at baseline were shipped frozen to a central biobank for storage at −80°C and were analyzed for laboratory parameters^89^. During follow-up of participants, incident events are adjudicated^89^. There are eight outcomes of interest herein: all-cause mortality, CV mortality, MI, stroke, 3-point Major Adverse Cardiovascular Events (MACE; defined as composite endpoint combining MI, stroke, CV death), 4-point MACE (defined as composite endpoint combining peripheral artery occlusive disease and related deaths in addition to 3-point MACE components), KF (defined as start of dialysis, kidney transplantation or death due to untreated KF) and AKI (based on Acute Kidney Injury Network criteria stage 3 per 2012 KDIGO guideline or temporary dialysis^90^.

In this project, data of 5,113 individuals with complete information, plasma metabolites and follow-up (median: 8.5 years) were utilised. The study was registered in the national registry for clinical studies (DRKS 00003971), approved by local ethics committees of participating institutions, and all participants provided written informed consent.

### Metabolomics

For all 3 studies circulating metabolites were profiled by Metabolon Inc. (Durham, NC) using Ultra-Performance Liquid Chromatography-Tandem Mass Spectrometry (UPLC-MS/MS). Analytical platforms, sample preparation, processing and metabolite coverage across the three cohorts are summarised in Supplementary Table 13. Urine ^1^H-Nuclear Magnetic Resonance (NMR) data were available only for MetaCardis. Specifically:

#### MetaCardis Serum Metabolomics

Serum samples, randomized across the study, were extracted and profiled by Metabolon Inc. using a UPLC–MS-based HD4 platform^27^. Metabolites and unknown features (denoted X-00000) were identified by automated matching of sample features with ion features, in a reference database of pure chemical standards and previously detected unknowns^91^, followed by detailed visual inspection and quality control by Metabolon, including repeatability, missing value imputation and batch stitching, as reported^92^. Missing values were imputed using the Half Minimum (HM) method^93^.

#### CLSA Plasma Metabolomics

Untargeted metabolomics were performed with UPLC-MS/MS methodology by Metabolon Inc. in CLSA plasma samples as previously described^94^, using the Metabolon Inc. HD4 platform. Quality control, batch correction and metabolite assignment were performed by Metabolon Inc. as described for MetaCardis above. We used the batch-normalised metabolite table, as provided by Metabolon Inc., with missing values imputed with the HM method, as for MetaCardis.

#### GCKD Plasma Metabolomics

Untargeted metabolomics were performed using stored plasma obtained on baseline as previously described^95,96^, similarly to MetaCardis and CLSA by Metabolon Inc. In this study, the batch-corrected imputed metabolite relative quantifications in GCKD, as performed by Metabolon Inc. were used. Missing values were imputed with the HM method as for MetaCardis and CLSA.

#### Urine H-NMR Metabolomics in MetaCardis

Spectra were acquired using an Avance spectrometer (Bruker) at 600eMHz; and chemical structures were assigned as extensively described previously^27,28^. In the present study we used absolute quantifications derived using the Brucker “In Vitro Diagnostics for research” (IVDr) quantification BI-Quant-UR algorithm (https://www.bruker.com/en/products-and-solutions/mr/nmr-clinical-research-solutions/b-i-quant-ur.html, v1.1).

### Shotgun metagenomics in MetaCardis

Gut metagenomic data were only available in MetaCardis and were processed as below:

#### MetaCardis Phylogenetic microbiota profiles

We built microbiota taxonomic profiles, after correction for bacterial load, as we have previously extensively described^25–27,80^ using a modified established protocol^97^. The 9.9 million gene integrated reference catalog of the human microbiome was used to perform gene abundance profiling as described^25–27,80^. Metagenomic data were processed using the following libraries: METEOR (v.3.2; https://forgemia.inra.fr/metagenopolis/meteor), Alientrimmer v0.4.0, Bowtie2 v2.3.4, MetaOMineR (momr, v1.31), Omixer-RPM (v1.0).

#### Gut microbial module analysis (GMM)

Microbial enzymatic module sets focusing on anaerobic bacterial and archaeal fermentation reactions were manually curated and assembled as previously described^25,27,28^.

### Statistical analyses

Statistical analysis was undertaken using R (v4.1.3; R Core Team (2022). R: A language and environment for statistical computing. R Foundation for Statistical Computing, Vienna, Austria; https://www.R-project.org/), unless otherwise indicated. For comparison of groups, we used non-parametric Mann–Whitney U (two groups) and Kruskal–Wallis (>2 groups) tests. Unadjusted Spearman correlation coefficients (ρ), we adjusted Spearman correlation coefficients with the R-package ppcor (v1.1). P values were corrected for multiple testing using the Benjamini–Hochberg (BH) method, as indicated.

### Machine Learning (ML) analysis

#### Data modalities

MetaCardis phenotypic variables were classed into ten groups (Supplementary Data 1), as previously described^28^. Briefly, biological parameters included biochemical and clinical serum laboratory tests, measured using standard enzymatic methods^27^. Serum creatinine and the derived CKD-EPI eGFRcr were excluded when bioclinical variables were used as predictors in Lasso models. Clinical parameters consisted of clinical history, clinical variables (BMI, systolic and diastolic blood pressure), anthropometric variables, and stool frequency and type. Demographic information included age, physical activity, educational and financial attainment, smoking, ethnicity and centre of recruitment. Drug variables reported intake of common medication, antibiotic courses in the last 5 years and total drugs per treatment category (antihypertensive, anti-diabetic, lipid-lowering) as described^26^. Nutritional parameters comprised habitual consumption of 37 food items, daily nutrients intake calculated and diet quality scores (alternative Healthy Eating (aHEI), Dietary Approaches to Stop Hypertension (DASH), Dietary Diversity (DDS) described in^85^. Clinical risk factors variables included age, sex, systolic and diastolic blood pressure, glycated haemoglobin levels, fasting cholesterol levels, smoking status and waist circumference as described^27,28^. Serum metabolomics comprised the relative levels of 1,514 circulating metabolites, excluding creatinine, determined by UPLC–MS/MS, as described above. Urine metabolomics included absolute quantification of 47 urine metabolites derived from ^1^H-NMR spectra, as described above. Microbiota variables included the abundance of 1,590 bacterial species present in at least 5% of MetaCardis participants after correction for microbial load and the first 10 principal components from a PCA of relative microbial gene abundances^28,98^. Microbiome (Modules) group included abundance of 116 manually curated bacterial modules, as described above. The full model comprised all variables across the 10 data modalities (3,600 variables in total; Supplemental Data 1). Categorical variables were converted into dummy variables using the caret library (v6.0.86).

#### Aggregated Least absolute shrinkage and selection operator (Lasso) eGFRcr models and external validation

To estimate eGFRcr in MetaCardis participants (N=1,723) with input each of the 10 data modalities defined above, we built Lasso regression-based eGFR models using the R library glmnet (v4.1.6). In MetaCardis, the prediction target, eGFRcr, was approximately normally distributed (Suppl.Fig.1A-B) and was therefore modelled untransformed. Overt CKD was an exclusion criterion in MetaCardis and we therefore postulated that training eGFR-predicting models in MetaCardis would capture kidney functional heterogeneity in the presence of common co-morbidities and would better generalise in independent cohorts. We employed bootstrap aggregation for model training and nested fivefold cross-validation to optimise the L1 regularization parameter, λ. Initially, we split our MetaCardis cohort into 5 equal parts with 4/5 as the training set and 1/5 as the test (left-out) set and mean-centred and variance-adjusted (z-scored) the input variables. We next used fivefold cross-validation in the training set (80% of participants) to determine the optimal regularization parameter lambda (from 200 values ranging from 10^−2^ to 10^1^) for predicting eGFR. We used the optimal λ to train a Lasso regression model in the training set and predicted eGFR in the left-out group (20% of the cohort). This process was repeated for each of the other four equal parts of the cohort, resulting in a predicted eGFR value for each participant which was then used to calculate the coefficient of determination using the rsq function from yardstick (v0.0.7) and an averaged β-coefficient from five models in each training split for each input variable. To account for cohort heterogeneity the process was repeated 100 times and the mean predicted eGFR and β-coefficients from all ensemble models were used in all subsequent analyses. In separate analyses, to exclude chance-biased predictions, eGFR was randomly permuted in the left-out group and predicted with metabolomics as input in all three studied cohorts, as indicated.

For metabolomics, which was the data modality best explaining eGFRcr variance, we also trained models in MetaCardis with 1,276 metabolites common between MetaCardis and CLSA; and with 1,152 metabolites common between MetaCardis, CLSA and GCKD, to predict z-scored eGFR (to account for different eGFR distributions between cohorts). We then used these models to predict eGFR in CLSA (N = 9,958) or GCKD (N=5,113) participants with available baseline metabolomics, as applicable. Metabolite intensities were z-scored within each cohort separately prior to modelling; the MetaCardis-trained models were applied to identically z-scored CLSA and GCKD data, without further cross-platform re-scaling and without information leakage between cohorts. To assess the robustness and reproducibility of the top metabolite selections, the eGFRcr-prediction Lasso ensemble (100 iterations, nested 5-fold cross-validation) was retrained *de novo* in CLSA on the 1,276 metabolites common to MetaCardis and CLSA, and per-metabolite selection frequencies were compared between the two cohort-specific ensembles. To uncover sex-specific metabolite panels relevant to kidney health we also performed model training as above exclusively in men or women participating in MetaCardis participants and predicted z-scored eGFR in males or females in CLSA or GCKD.

#### Metabolomics Kidney Function Gap (mKFG) calculation

To calculate personalized metabolomic kidney function gaps (mKFGs) for each participant of the three studies, conventional creatinine-based eGFRcr was subtracted from eGFR predictions of models trained in MetaCardis on serum metabolomics, as applicable. This is conceptually similar to proteomic ageing gaps^11^ and multi-omic BMI signatures^18,19^, defined as the numerical difference between a clinical reference variable and an omics-derived prediction of that variable. For association studies between mKFGs and biochemical variables or health outcomes gaps were adjusted with the corresponding conventional creatinine-based eGFRcr obtained at recruitment, in line with similar research with age gaps, to account for the well-documented tendency of ML algorithm predictions to revert to the mean of the predicted variable^45,46^, including eGFR^11^. This was achieved either by including conventional eGFR as covariate in corresponding models, as appropriate, or by regressing eGFR from mKFGs by extracting residuals from a linear model with kidney gaps as the dependent and conventional creatinine-based eGFR as the independent variable. To allow for direct comparisons between models, kidney function gaps from each model were z-scored.

#### Physiology-guided feature selection strategy

To discover metabolites strengthening associations with cardiovascular-kidney-metabolic phenotypes we adapted the permutation feature importance (PFI) method^99^, as further refined by Oh and colleagues for health-relevant outcomes^11^. We did not seek to identify metabolites that after permutation decrease the accuracy of predicted eGFRcr but instead focused on permuted metabolites that modify association between kidney gaps (predicted minus eGFRcr) with biochemical variables indicative of CKM health. In the absence of a universally accepted score of kidney health and biochemical metrics of kidney function (uACR, urea, urate, cystatin C, calcium phosphate) in our non-CKD cohorts (MetaCardis and CLSA), we optimised our models in MetaCardis with serum haemoglobin, serum albumin or the Neutrophil-to-Lymphocyte ratio as the target variables. These variables were selected in a data-driven way due to their association with mKFGall but not eGFRcr (Fig.2E). We reasoned that optimising our models towards these variables could integrate and reflect interlinked multi-organ information relevant to CKM health independently to that already contained in eGFRcr. A metabolite that after its permutation in the test set, kidney gaps using the resulting eGFR prediction after correcting for age, sex, creatinine-based eGFR and centre of recruitment, exhibited diminished Spearman ρ with the target variable than gaps without permutation, was deemed relevant in capturing associations between gaps and target variable. The feature importance (FI) score for each metabolite was calculated as the % difference between partial Spearman ρ between the target variable and the kidney gaps with or without permutation and was used to rank metabolites.

We applied this strategy to 436 metabolites (out of 1,276 in total) that were included with a non-zero value in a minimum of 20% of our eGFR-predicting Lasso models in MetaCardis, using the same cut-off as Oh and colleagues^11^ and defined metabolite panels for haemoglobin, serum albumin and NLR. We also defined an intersected metabolomics panel with metabolites that strengthen mKFG associations with all three target variables, removing drugs. The intersected metabolite panels were then used to train Lasso models in MetaCardis as described above.

To test whether optimising mKFGs towards any cardiometabolic-health-relevant variable serendipitously improves associations of mKFGs with CKM outcomes, we applied the same physiology-guided optimisation pipeline (as described above) to 100 random three-anchor triplets sampled from a 17-variable pool (HOMA.IR, FFA, CCL2, Eotaxine, HDL, TG, HbA1c, IL7, Insulin, IP10, ALT, MIF, MIP1, SDF1, sodium, VEGF, Monocytes) of cardiovascular- and metabolic-disease-relevant clinical biochemistry not significantly associated (FDR > 0.1) with either eGFRcr or mKFG_all_ in MetaCardis. Microalbumin and total neutrophils/lymphocytes were excluded as direct kidney-damage and NLR-overlapping markers, respectively. Lasso models trained in MetaCardis were applied to CLSA to predict eGFR and generate negative-control mKFGs, with downstream analyses restricted to the 36 triplets whose eGFRcr predictions in CLSA achieved R^2^ comperable to that achieved by models using all common metabolites between CLSA and MetaCardis (R^2^ > 0.4). These negative-control mKFGs were tested against four future cardiorenal endpoints in CLSA (eGFRcr decline rate, incident CKD, incident MI, all-cause mortality), using the same model specifications as for mKFG_int_ (described above).

#### XGBoost and SHapley additive exPlanations (SHAP) analysis

We also predicted eGFR levels in all MetaCardis participants (N=1,723) with serum metabolomics as input using gradient-boosting decision trees based on the xgboost algorithm (v1.3.2.1)^100^, as we previously described^28^, with modifications. We used two nested fivefold cross-validations to optimise the xgboost hyperparameters (eta, max_depth, min_child weight, gamma, lambda, colsample_bytree; for additional information on model parameters please see the XGBoost documentation (https://xgboost.readthedocs.io/en/latest/index.html) in the training cohort independently of the test cohort, as described for Lasso above. Briefly, we used a hyperparameter grid search of 1350 parameter combinations with early stopping set to 10 to identify the optimal parameters in each training set, evaluating model outcomes using the root-mean-square error (RMSE) metric. After parameter optimisation, eGFR was predicted in the left-out group (20% of cohort) with a model trained with the optimal parameters in the training group (80% of cohort) and this was applied to the other four training-test set pairs. Like Lasso and to account for cohort heterogeneity we repeated the process for 20 times and averaged eGFR predictions.

To assign relative importance to metabolites driving XGBoost eGFR predictions we used SHAP values, as implemented in tree-based models^101^ and recently reported by us^28^ and others^98^. Metabolite SHAP values for each individual in the five left-out groups were extracted from corresponding XGBoost models trained on the remaining 80% participants, as described above, using SHAPforxgboost (0.1.0) and averaged over 20 iterations. For each metabolite relative importance was assigned by calculating the mean absolute SHAP value for each MetaCardis participant, as described^28^. For swarm plots depicting each individual participants SHAP values, metabolite relative levels were inverse-normalised using the RNOmni (v1.0.0) package.

#### Logistic regression

For logistic regressions for prevalent disease at recruitment or incident events without time-to-event data in MetaCardis or CLSA, we used base the R generalised linear model function (glm) with family set to binomial with mKFGs, eGFR or other input variables z-scored to allow for direct comparison between models. Age, sex and in the case of MetaCardis country of recruitment were also included as covariates in the models. In certain models we also included the Framingham Risk score, serum albumin, haemoglobin or NLR (all z-scored), as indicated in the text. Likelihood Ratio Tests (LRTs) between nested models with or without kidney gaps, were performed using the base R anova function using the Likelihood Ratio (“LR”) test option throughout.

#### Cox proportional hazard models

To assess associations between mKFGs and all-cause mortality in CLSA, or 8 CKM events in GCKD with time-to-event data we used Cox proportional hazard models, as implemented in the survival library (v3.7.0) in R. For GCKD, competing events (i.e., death for other reasons than included in considered outcomes) were available and treated as censored observations (i.e., cause-specific analysis was conducted). In GCKD, study entry was defined as the time origin and the time scale was time since study entry; follow-up was restricted to the first 8.5 years after baseline. Time-to-event was defined as the interval from study entry to the event of interest. Participants who did not experience the event of interest were censored at the earliest of: date last known alive, date of death from causes unrelated to the event of interest, or 8.5 years after baseline. The censoring of deaths unrelated to the event of interest (competing events) in the analysis corresponds to a cause-specific analysis. To determine how covariates influence associations between mKFGs and CKM events we sequentially added covariates to the models that had this general structure: event riske≈emKFGe+eeGFR + covariates and compared them with nested models where only eGFR and the same covariates were included. To allow for direct model Hazard Ratio (HR) comparisons kidney gaps or eGFRcr were z-scored. In CLSA models included: Model 1: eGFRcr only; Model 2: eGFRcr, age and sex; Model 3: eGFRcr, age, sex and the Framingham 10-year Risk score^24^, as an indicator of baseline cardiovascular risk. In GCKD, Models 1 and 2 were identical with Model 3 including eGFRcr, age, sex and uACR whilst Model 4 included in addition FRS and in the case of kidney outcomes the 5-year Kidney Failure Risk (KFRE) equation^22,23^. C-statistics for each model along with 95% Confidence Intervals (CIs) were extracted from the models. Nested models with or without mKFGs, were compared using the base R anova function using the Likelihood Ratio (“LR”) test option. Informativeness, as a measure of improvement in the “goodness-of-fit” of the data with models including kidney gaps in comparison to baseline models, was calculated as the ratio of the Wald test χ2 statistic of the two models as described in^65^. Model calibration was assessed using bootstrap-corrected calibration slopes (500 resamples) computed with the validate and calibrate functions of the rms package (v6.8-1) in R, where a slope of 1 indicates perfect agreement between predicted and observed risk. As sensitivity analyses in GCKD, we repeated the fully-adjusted Cox models replacing eGFRcr with combined creatinine-cystatin C eGFR (eGFRcys_cr; CKD-EPI 2021^67^), and assessed the incremental performance of mKFGs within KDIGO eGFR and albuminuria risk categories^62^. Detection Rate curves across different False Positive Range ranges for tested outcomes in CLSA and GCKD were computed from the most stringent Cox regression models (Model 3 in CLSA and Model 4 in GCKD) with the riskRegression (v2023.3.22) library in R (v4.4.1).

Cumulative incidence curves for CLSA or GCKD. Participants were assigned into tertiles (low, medium, high) of mKFG residuals computed as follows. mKFGs were regressed on baseline eGFRcr using a linear model as described above. Residuals were then z-scored using the R scale() function so that per-SD effect sizes are comparable across cohorts. Participants were then assigned to tertiles using dplyr::ntile(residuals, 3), which produces approximately equal-sized groups corresponding to the 33rd and 67th percentile boundaries of the residual rank distribution. Cumulative incidence curves were then derived with the survival package and visualized with the ggsurvfit (v.1.1.0) package in CLSA and for GCKD with the Python (v.3.12.0) lifelines (v.0.27.8) package.

#### Framingham risk score calculation

The Framingham 10-year Cardiovascular Risk score^24^ (FRS) was computed for participants below 74years of age of our three cohorts using the CVrisk (v1.1.1) R library with age, sex, total cholesterol, HDL cholesterol, Systolic Blood Pressure, hypertensive medication intake, T2D incidence and smoking status.

#### 5-year Kidney Failure Risk Equation

Five-year risk of progression of CKD to Kidney Failure (KF) within 5 years was estimated using Tangri’s Kidney Failure Risk Equation (KFRE) implemented in the KFRE: Kidney Failure Risk Estimator package for Python (https://github.com/lshpaner/kfre?tab=readme-ov-file#references). This implementation applies a 4-variable version of the Kidney Failure Risk Estimator^22,22^ (KFRE), using the patient’s age, biological sex, eGFR, and urinary albumin creatinine ratio. This risk estimate was subsequently used to adjust our models and is referred to as KFRE, as described above.

#### Benchmarking against a metabolome-derived GFR

To test whether mKFGint reflects more than improved filtration estimation, we derived a metabolome-based GFR estimate (eGFR_metF) in CLSA using the creatinine-and demographics-free metabolite equation of Coresh et al. (2019)^66^: eGFR_metF = exp(3.04584 − 0.450817·ln[N-acetylthreonine] − 0.214876·ln[pseudouridine] − 0.253004·ln[myo-inositol] + 0.2265693·ln[tryptophan]), applied to the batch-normalised relative metabolite abundances. Because we applied this equation - fitted on absolute targeted concentrations - to relative metabolite abundances, eGFR_metF is an approximation of the estimate in Coresh et al.^66^ rather than an exact value. Metabolon relative abundances are normalised across the whole spectrum, so an individual metabolite’s relative value is not strictly proportional to its absolute concentration. We therefore used eGFR_metF only as a z-scored model covariate, a metabolome-derived filtration estimate against which to benchmark mKFG_int_, not a calibrated metabolic GFR.

## Supplemental Figure Legends

**SP.Fig.1. eGFRcr in MetaCardis is normally distributed. (A)** Density plot of eGFRcr in men (purple) or women (green) MetaCardis participants. (**B**) Quantile-quantile (Q-Q) plots describing the relationship between observed and expected for normal distribution statistics under the null for eGFRcr in MetaCardis. Scatter plots of eGFRcr (**C**) or metabolomic kidney function gaps (mKFGs) (**D**) versus age. The mKFG are defined as the predicted eGFRcr from Lasso regression (using metabolomics as input) minus the conventional eGFRcr. The line depicts the linear regression line and the 95% confidence interval. In the inset, Spearman ρ and P-values for the associations between age and either eGFR or mKFGs (corrected for baseline eGFR) are depicted.

**SP.Fig.2. Lasso predicts eGFRcr similarly to XGBoost in MetaCardis.** (**A**) Volcano plot of β-coefficients (x-axis) from linear models of individual (z-scored) metabolites used in Lasso-based eGFRcr prediction in MetaCardis. The y-axis shows the -log_10_ of Benjamini-Hochberg (BH) false discovery rate (FDR)–adjusted p-values. Red dots signify significant (FDR<0.1) positive associations and blue dots negative. (**B**) Boxplots of Explained variance in eGFRcr in MetaCardis with all available Metabolites (1,514 metabolites excluding creatinine) from Lasso or XGBoost algorithms (“*Methods*”) as indicated. (**C**) Scatterplot illustrating the association between predicted and calculated eGFRcr from XGBoost algorithms trained with metabolites in MetaCardis. Each participant was coloured by the difference between the predicted value minus the calculated value (metabolomics Kidney Function Gap (mKFG)) according to the shown scale. The red line depicts the locally weighted scatterplot smoothing (LOESS) with 95% CI shaded in gray. (D) Swarm plots illustrating the impact on eGFRcr predictions (SHAP values) from the XGBoost model for the top 20 circulating metabolites (based on highest absolute SHAP value) in MetaCardis participants (Ne=e1,723). Metabolites are ordered in descending mean absolute SHAP values across all participants. Each dot represents a single participant, coloured by the inverse-normalised value for the corresponding metabolite.

**SP.Fig.3. Sex-stratified models to predict eGFRcr**. (**A**) Explained variance of z-scored eGFRcr from models trained in MetaCardis with all MetaCardis participants (All), men only (Men) or women only (Women) with serum metabolomics as input. Scatterplots showing associations between predicted and conventional eGFRcr from Lasso algorithms trained with metabolomics exclusively with men (**B**) or women (**C**) MetaCardis participants. Each participant was coloured by the mKFG according to the shown scale and the Spearman ρ and P-value between the calculated and predicted eGFRcr is given. Averaged metabolite β-coefficients from 100 Lasso models trained with Metabolomics in men (**D**) or women (**E**) MetaCardis participants. Metabolites present in more than 80% of models are coloured blue. (**F**) Distribution of metabolomics kidney function gaps (mKFGs) after baseline eGFRcr was regressed in men or (**G**) women MetaCardis participants.

**SP.Fig.4. Association of sex-stratified kidney gaps (mKFG) with cardiometabolic parameters in MetaCardis and external validation in CLSA.** Partial Spearman correlations between baseline eGFRcr or sex-stratified mKFGs and biochemical variables in men (**A**) or women (**B**) MetaCardis participants. Correlations were adjusted for: Model 1: age, Model 2: age and country of recruitment (Centre), Model 3: baseline eGFRcr, Model 4: baseline eGFRcr and age, Model 5: baseline eGFRcr, age and country of recruitment (Centre) and corrected for multiple testing with the Benjamini-Hochberg method; *FDR<0.1. Colour and intensities depict the direction and strength of the association as shown in the corresponding scale. (**C**) Validation in CLSA. Explained variance of z-scored eGFRcr (CLSA) from models trained in MetaCardis on all shared metabolites (1,276) across the cohorts. Training was performed using (i) all MetaCardis participants (All), (ii) men only (Men), and (iii) women only (Women).

**SP.Fig.5. Physiology-driven optimisation of models predicting eGFR in men participating in MetaCardis**. (**A**) Most influential metabolites derived from feature selection for the targeted biochemical variables (serum albumin, serum haemoglobin, Neutrophil-to-Lymphocyte Ratio (NLR)) in men participating in MetaCardis. X-axis depicts the difference in eGFRcr variance explained (R^2^) after each given metabolite was randomly permuted in the test set. Y-axis depicts difference in correlation (adjusted for age, country of recruitment and eGFRcr) with each targeted biochemical variable, when each metabolite was permuted in the test set. Positive values denote percentage reduction in the strength of association after permutation in the test-set. Metabolites that after permutation change the absolute adjusted Spearman ρ by more than 7% are named and coloured by the corresponding target variable. (**B**) Venn diagram of metabolites strengthening associations with each target biochemical variable. 98 metabolites strengthen associations for all three targeted variables and are used to train the intersected model. (**C**) Explained Variance of eGFRcr in men participating in MetaCardis with models trained with the intersected metabolite panel. We also include predictions from the common (Common; 1,276) metabolites model for comparison. (**D)** Spearman ρ from partial correlations (corrected for age, country of recruitment and calculated eGFR for mKFGs) between eGFRcr, the intersected metabolite panel (mKFG_int_) and gaps derived from models trained with all common metabolites (1,276) with the targeted biochemical variables. (**E**) Averaged metabolite β-coefficients from 100 Lasso models trained with the intersected metabolomics panel in men participating in MetaCardis. Metabolites present in more than 80% of models are coloured blue. (**F**) Partial Spearman correlations between intersected mKFG and those from models with all metabolites with biochemical variables in men participating in MetaCardis. Correlations were adjusted for age and country of recruitment. Colour (hue) indicates the direction (positive/negative) of the correlation, and colour intensity reflects its strength, following the scale provided. (**G**) Forest plot illustrating standardised odds ratios per SD of mKFGs or eGFRcr for prevalent hypertension (HT), T2D, coronary artery disease (CAD) and atrial fibrillation (AF) (number of cases in parentheses) in men participating in MetaCardis. LRT: Likelihood Ratio Test between the eGFR model and the model including mKFG for each disease. All logistic regressions included age, country of recruitment and for mKFGs baseline eGFRcr as covariates.

**SP.Fig.6. Physiology-driven optimisation of models predicting eGFR in women participating in MetaCardis**. (**A**) Most important metabolites derived from physiology-driven optimisation for the targeted biochemical variables (serum albumin, serum haemoglobin, Neutrophil-to-Lymphocyte Ratio (NLR)) in women MetaCardis participants. X-axis depicts the percentage difference (FI: Feature Importance) in eGFRcr variance explained (R^2^) after each given metabolite was randomly permuted in the test set. Y-axis depicts percentage difference (FI: Feature Importance) in correlation with each targeted biochemical variable, when each metabolite was permuted in the test set. Positive values denote reduction in the strength of association after permutation in the test-set. Metabolites that after permutation change the absolute adjusted Spearman ρ by more than 7% are named and coloured by the relevant target variable. (**B**) Venn diagram of metabolites strengthening associations with each target bioclinical variable in women. 46 metabolites strengthen associations for all three targeted variables and are used to train the intersected model. (**C**) Explained Variance of eGFRcr in women MetaCardis participants with models trained with the 46 intersected metabolites. We also include predictions from the common metabolites model (1,276) for comparison. (**D)** Spearman ρ from partial correlations (corrected for age, country of recruitment and eGFRcr for mKFGs) between eGFRcr, the intersected metabolite panel (mKFG_int_) and gaps derived from models trained with all common metabolites with the targeted biochemical variables. (**E**) Averaged metabolite β-coefficients from 100 Lasso models trained with the combined metabolomics panel in female MetaCardis participants. Metabolites present in more than 80% of models are coloured blue. (**F**) Partial Spearman correlations between mKFG_int_ and mKFG_all_ (as defined in (D)) with biochemical variables in women MetaCardis participants. Correlations were adjusted for age and country of recruitment. Colour (hue) indicates the direction (positive/negative) of the correlation, and colour intensity reflects its strength, following the scale provided. (**G**) Forest plot illustrating standardised odds ratios per SD of mKFG or eGFRcr for prevalent hypertension (HT), T2D, coronary artery disease (CAD) and atrial fibrillation (AF) (number of cases in parentheses) in women MetaCardis participants. LRT: Likelihood Ratio Test between the eGFRcr model and the model including KFG for each disease. All logistic regressions included age, country of recruitment and for mKFGs calculated eGFRcr as covariates.

**SP.Fig.7. Validation of models trained with men in MetaCardis in CLSA men participants**. (**A**) Explained variance (EV) of eGFRcr in CLSA men participants predicted with models trained in men participants of MetaCardis with the intersected metabolites (98 metabolites; Int) or all common metabolites (1,276 metabolites; Common). (**B**) scatterplot showing the association between predicted and conventional creatinine-based eGFRcr from the model with intersecting metabolites and the model with all common metabolites. LOESS curves are also shown for each model (Int: intersected panel (blue), All: all common metabolites (purple)). Individuals are coloured by mKFG_int_ according to the depicted scale. (**C**) Density plots showing the distribution of mKFG_all_ (All; purple) and mKFG_int_ (blue) after adjustment for baseline conventional eGFRcr in men participants of CLSA derived from models trained in MetaCardis men participants and shown in B. (**D**) Spearman ρ from partial correlations (corrected for age or age and eGFRcr for mKFGs) between eGFRcr, mKFG derived from models trained with intersecting metabolites or all metabolites with targeted biochemical variables in men participants of CLSA. (**E**) Partial Spearman correlations between baseline eGFRcr (corrected for age) or mKFGs (corrected for age and creatinine-based eGFRcr) derived from models trained with intersecting metabolites or all metabolites and biochemical variables in men CLSA participants. Colour (hue) indicates the direction (positive/negative) of the correlation, and colour intensity reflects its strength, following the scale provided. (**F**) forest plot of standardised odds ratios for z-scored kidney mKFG or eGFRcr in relation to the baseline prevalence of hypertension (HT), T2D, Myocardial infarction (MI) or cerebrovascular disease (CVA) in CLSA. Likelihood Ratio Tests (LRT) were performed to compare baseline models including conventional eGFRcr and age against models additionally including indicated mKFG in men CLSA participants with available metabolomics. Prevalence for each studied pathology in parentheses. For comparison, mKFG were computed using models trained in MetaCardis with either all participants and all common metabolites (mKFG_all__all) or the universal intersecting 66-metabolite panel (mKFG_int__all). LRT deviance between the gap derived from men-trained models (mKFG_all_, mKFG_int_) and gaps in men derived from predictions with universal models (mKFG_all__all, mKFG_int__all) for each pathology are also given.

**SP.Fig.8. Validation of models trained with females in MetaCardis in CLSA women participants**. Explained variance (EV) of eGFRcr in CLSA women participants predicted with models trained in women participants of MetaCardis with the intersected metabolites (46 metabolites; Int) or all common metabolites (1,276 metabolites; Common). (**B**) scatterplot showing the association between predicted and calculated eGFRcr from the model with intersecting metabolites and the model with all common metabolites. LOESS curves are shown for the intersected model (blue) and the model with all common metabolites (purple). Individuals are coloured by mKFG_int_ according to the depicted scale. (**C**) Density plots showing the distribution of mKFG_all_ (purple) and mKFG_int_ (blue) after adjustment for baseline eGFRcr in women participants of CLSA derived from models trained in MetaCardis women participants and shown in B. (**D**) Spearman ρ from partial correlations (corrected for age or age and creatinine-based eGFRcr for mKFGs) between eGFRcr, mKFG derived from models trained with intersecting metabolites or all common metabolites and targeted biochemical variables in women CLSA participants. (**E**) Partial Spearman correlations between baseline eGFRcr (corrected for age) or mKFGs (corrected for age and conventional eGFRcr) derived from models trained with intersecting metabolites or all metabolites and biochemical variables in women CLSA participants. Colour (hue) indicates the direction (positive/negative) of the correlation, and colour intensity reflects its strength, following the scale provided. (**F**) forest plot of standardised odds ratios for z-scored kidney mKFG or eGFRcr in relation to the baseline prevalence of hypertension (HT), T2D, Myocardial infarction (MI) or cerebrovascular disease (CVA) in CLSA. Likelihood Ratio Tests (LRT) were performed to compare baseline models including eGFRcr and age against models additionally including indicated mKFG in women CLSA participants with available metabolomics. Prevalence for each studied pathology in parentheses. For comparison, mKFG were also computed using models trained in MetaCardis with either all participants and all common metabolites (mKFG_all__all) or the universal intersecting 66-metabolite panel (mKFG_int__all). LRT deviance between the gap derived from women-trained models (mKFG_all_, mKFG_int_) and gaps in women derived from predictions with universal models (mKFG_all__all, mKFG_int__all) for each pathology are also given.

**SP.Fig.9. eGFRcr rate of decline in men and women CLSA participants**. Scatterplot illustrating the association between eGFRcr drop rate (%) in men (**A**) or women (**B**) CLSA participants from baseline to the first follow-up (3 years) versus z-scored mKFG computed at baseline derived with models using intersecting metabolites (dark blue), all metabolites (purple), yellow for eGFR and light green for Framingham risk score (FRS). LOESS with all mKFG_all_, mKFG_int_, FRS and eGFRcr. Inset display partial Spearman correlation coefficients (ρ) for the rate of eGFRcr decline corrected for age and in the case of mKFGs also for calculated eGFRcr at baseline.

**SP.Fig.10. Calibration of all-cause mortality prediction in CLSA.** Bootstrap-corrected calibration of Cox models for all-cause mortality in Framingham-eligible CLSA participants (age <75; N = 7,556; 175 deaths), evaluated at t = 6.85 years (2,500 days). **(A)** Baseline model: eGFRcr + age + sex + Framingham Risk Score (FRS). **(B)** Baseline + mKFG_all_. **(C)** Baseline + mKFG_int_. In each panel the x-axis is model-predicted and the y-axis observed mortality; the dashed grey line denotes perfect calibration (observed = predicted), the black line the apparent and the blue line the 500-iteration bootstrap-corrected calibration (“Methods”). Insets report the bootstrap-corrected calibration slope and Harrell’s C-index. All three models are well calibrated (slope 0.96–0.98).

**SP.Fig.11. Negative control of physiology-guided optimisation pipeline in CLSA.** The physiology-guided optimisation pipeline used to derive mKFG_int_ was applied to 100 random triplets of cardiovascular- and metabolic-disease-relevant biochemical variables uncorrelated with eGFRcr and mKFG_all_ in MetaCardis. Triplets whose optimised gap tracked eGFRcr with cross-validated explained variance R² > 0.4 (36/100) were retained and compared with mKFGint across four cardiovascular-kidney-metabolic outcomes in CLSA: rate of eGFRcr decline (partial Spearman ρ, adjusted for age and baseline eGFRcr); incident chronic kidney disease (CKD) and incident myocardial infarction (MI) (odds ratios, adjusted for eGFRcr, age and sex); and all-cause mortality (hazard ratio, adjusted for eGFRcr, age, sex and the Framingham Risk Score, FRS). A triplet "beats" mKFG_int_ for an outcome when its effect estimate is more extreme (stronger association) than that of mKFG_int_. (A) Number of the 36 retained triplets beating mKFG_int_ on at least N of the four outcomes (cumulative counts). (B) Per-outcome dominance; each row is one triplet labelled by its three anchor variables with the number of outcomes beaten in parentheses (n/4). Blue indicates the triplet beats mKFG_int_ for that outcome and grey that it does not.

**SP.Fig.12. mKFGs and CLSA cardiorenal outcomes before and after direct adjustment for haemoglobin, albumin and the Neutrophil-to-Lymphocyte Ratio.** In every panel, estimates are shown for a core model (filled circles) and for the same model additionally adjusted for haemoglobin (Hb), serum albumin and the Neutrophil-to-Lymphocyte ratio (NLR) (+target; hollow circles). mKFG_all_ and mKFG_int_: metabolomic Kidney Function Gaps derived from all common metabolites or from the intersected panel; eGFRcr, creatinine-based eGFR; FRS, Framingham Risk Score. **(A)** Scatterplot of z-scored mKFGs, eGFRcr and FRS against the annual rate of eGFRcr decline (%) over the first 3-year follow-up, with LOESS curves; insets give partial Spearman correlation coefficients (ρ) for the rate of decline (corrected for age, sex and for mKFGs additionally for baseline eGFRcr) plus the target variables. **(B–D)** Forest plots of odds ratios per SD of each predictor (mKFG, eGFRcr or FRS) for **(B)** rapid eGFR decline, **(C)** incident chronic kidney disease (CKD) and **(D)** incident myocardial infarction (MI), hospitalisation, incident type 2 diabetes (T2D) and incident stroke; core logistic models are adjusted for eGFRcr, age and sex. **(E)** Forest plot of hazard ratios per SD of mKFG or eGFRcr for all-cause mortality in Framingham-eligible participants; the core model is eGFRcr + age + sex + FRS (Model 4) and +target additionally includes Hb, albumin and NLR (Model 5). The C-statistic (95% CI), likelihood-ratio test (LRT) P value, improvement in informativeness ("goodness-of-fit") and Net Reclassification Index (NRI) **versus** the corresponding baseline model are shown. **(F)** Detection rate (DR) versus false-positive rate (FPR) for all-cause mortality at t = 6.5 years, from Cox models in (E). All analyses are in CLSA participants with available metabolomics.

**Suppl.Fig.13. Cross-cohort reproducibility of the eGFRcr-prediction metabolite panel**. Per-metabolite selection frequency (% of 100 bootstrap-aggregated Lasso iterations) in MetaCardis (training cohort) versus a Lasso retrained de novo in CLSA on the 1,276 metabolites common to both cohorts, for the 40 metabolites with the highest absolute β-coefficient in MetaCardis. Dashed line, the 20% selection threshold; 35 of these 40 leading MetaCardis predictors are also selected in ≥20% of CLSA models.

**SP.Fig.14. mKFGs with baseline renal readouts and with eGFRcys in GCKD**. (**A**) Scatterplot showing associations between urinary Albumin Creatinine Ratio (uACR) with eGFRcr. Spearman partial correlation coefficients (ρ), adjusted for age and sex, are provided for the relationships between eGFRcr and uACR. (**B**) Cumulative event probabilities for Myocardial infarction (MI), Stroke, all-cause Mortality, Cardiovascular (CV) Mortality, Kidney Failure (KF), Acute Kidney Injury (AKI), 3-and 4-point Major Adverse Cardiovascular Events (MACE) in GCKD participants over an 8.5-year follow-up. Kidney gaps (mKFG_all_) were computed from eGFRcr predictions using models trained in MetaCardis with all common metabolites. To account for baseline eGFRcys, mKFGall was regressed on eGFRcys, and the residuals were used for stratification. Participants were divided into tertiles based on these residuals, with the low tertile representing those with the worst (most negative) residuals and the high tertile representing those with the best (most positive) residuals. (**C**) As in (B) for mKFG_int_ derived from eGFR predictions from models trained in MetaCardis with the intersected metabolite panel (60 metabolites). (**D-G**) (**left panels**) forest plots of hazard ratios per SD of eGFR and mKFGs computed with models with all or the intersected panel metabolites for Kidney Failure (KF), stroke, Myocardial Infarction (MI) and Cardiovascular (CV) in GCKD over 8.5 years. Cox regression was performed using the following models: Model 1: baseline eGFRcr also included in the mKFG model; Model 2: baseline eGFRcr, age and sex; Model 3: baseline eGFRcr, age, sex, urinary albumin creatinine ratio (uACR); Model 4: as model 3 with the Framingham score (FRS) or the 5-year Kidney Failure Risk Equation (KFRE) for KF, for AKI both FRS and KFRE were included in model 4. The C-statistic with 95% confidence intervals (in parentheses) for each model are displayed together with the Likelihood Ratio Test (LRT) P value, the improvement in the Informativeness (“goodness-of-fit”) of models and Net Reclassification Index (NRI) when compared to the corresponding baseline model. (**Right panels**) Detection Rates (DRs) across different False Positive Rate Thresholds (FPR) for events in (D-G) derived from the corresponding Cox regression models with the most stringent model (Model 4) containing kidney gaps from all or the intersected panel metabolites.

**SP.Fig.15. Calibration of all-cause mortality prediction in GCKD**. Bootstrap-corrected calibration of Cox models for all-cause mortality in GCKD participants (N = 4,776; 847 deaths), evaluated at t = 8.5 years (3,105 days). (A) Baseline model: eGFRcr + age + sex + Framingham Risk Score (FRS) + urinary albumin-to-creatinine ratio (uACR). (B) Baseline + mKFG_all_. (C) Baseline + mKFG_int_. In each panel the x-axis is model-predicted and the y-axis observed mortality; the dashed grey line denotes perfect calibration (observed = predicted), the black line the apparent and the blue line the 500-iteration bootstrap-corrected calibration ("**Methods**"). Insets report the bootstrap-corrected calibration slope and Harrell’s C-index.

**SP.Fig.16. mKFGs reveal divergent cardiovascular-kidney-metabolic disease trajectories in men participants of GCKD.** (**A**) explained variance (EV) of eGFRcr in men GCKD participants (N=3,071) predicted with models trained in men participants of MetaCardis with the men-specific intersected metabolites panel (91) or all common metabolites (1,152) between all studied cohorts. (**B**) Association between predicted and conventional eGFRcr from the model trained on intersecting metabolites and the model containing all common metabolites in men GCKD participants. LOESS curves are also shown for both models along with unadjusted Spearman ρ. (**C**) Cumulative event probabilities for Myocardial infarction (MI), Stroke, All-cause Mortality, Cardiovascular (CV) Mortality, Kidney Failure (KF), Acute Kidney Injury (AKI), 3-and 4-point Major Adverse Cardiovascular Events (MACE) in men participants of GCKD over 8.5-year follow-up. Kidney gaps (mKFG_all_) were computed from eGFRcr predictions using models trained in MetaCardis with all common metabolites. To account for baseline eGFRcr, mKFG_all_ was regressed on eGFRcr, and the residuals were used for stratification. Participants were divided into tertiles based on these residuals, with the low tertile representing those with the worst (most negative) residuals and the high tertile representing those with the best (most positive) residuals (**D**) As in (C) for kidney gaps derived from models trained in MetaCardis with the men-specific intersected metabolite panel (91 metabolites). (**E-L**) Forest plots of hazard ratios per SD of eGFRcr and mKFGs for 3-point MACE, 4-point MACE, MI, stroke, KF, AKI, all-cause mortality and CV mortality computed with models with all or the intersected panel metabolites in men participants of GCKD over 8.5 years. Cox regression was performed using the following models: Model 1: baseline conventional eGFRcr also included in the mKFG model; Model 2: baseline eGFRcr and age; Model 3: baseline eGFRcr, age, urinary albumin creatinine ratio (uACR); Model 4: as model 3 with the Framingham score (FRS) or the 5-year Kidney Failure Risk Equation (KFRE) for KF, for AKI both FRS and KFRE were included in model 4. The C-statistic with 95% confidence intervals (in parentheses) for each model are displayed together with the Likelihood Ratio Test (LRT) P value, the improvement in the Informativeness (“goodness-of-fit”) of models and Net Reclassification Index (NRI) when compared to the corresponding baseline models not containing mKFGs.

**SP.Fig.17. mKFGs reveal divergent cardiovascular-kidney-metabolic disease trajectories in women participants of GCKD.** (**A**) explained variance (EV) of eGFRcr in women GCKD participants (N=2,042) predicted with models trained in women participants of MetaCardis with the women-specific intersected metabolites panel (40) or all common metabolites (1,152) between the three studied cohorts. (**B**) Association between predicted and calculated eGFRcr from the model trained on intersecting metabolites and the model containing all common metabolites in women GCKD participants. LOESS curves are also shown for both models along with unadjusted Spearman ρ. (**C**) Cumulative event probabilities for Myocardial infarction (MI), Stroke, All-cause Mortality, Cardiovascular (CV) Mortality, Kidney Failure (KF), Acute Kidney Injury (AKI), 3-and 4-point Major Adverse Cardiovascular Events (MACE) in women participants of GCKD over 8.5-year follow-up. Kidney gaps (mKFG_all_) were computed from eGFRcr predictions using models trained in MetaCardis with all common metabolites. To account for baseline eGFRcr, mKFG_all_ was regressed on eGFRcr, and the residuals were used for stratification. Participants were divided into tertiles based on these residuals, with the low tertile representing those with the worst (most negative) residuals and the high tertile representing those with the best (most positive) residuals (**D**) As in (C) for kidney gaps derived from models trained in MetaCardis with the female-specific intersected metabolite panel (40 metabolites). (**E-L**) Forest plots of hazard ratios per SD of eGFR and mKFGs for 3-point MACE, 4-point MACE, MI, stroke, KF, AKI, all-cause mortality and CV mortality computed with models with all or the intersected panel metabolites in women participants of GCKD over 8.5 years. Cox regression was performed using the following models: Model 1: baseline conventional eGFRcr also included in the mKFG model; Model 2: baseline eGFRcr and age; Model 3: baseline eGFRcr, age, urinary albumin creatinine ratio (uACR); Model 4: as model 3 with the Framingham score (FRS) or the 5-year Kidney Failure Risk Equation (KFRE) for KF, for AKI both FRS and KFRE were included in model 4. The C-statistic with 95% confidence intervals (in parentheses) for each model are displayed together with the Likelihood Ratio Test (LRT) P value and the improvement in the Informativeness (“goodness-of-fit”) of models and Net Reclassification when compared to the corresponding baseline models not containing mKFGs.

## Supplemental Tables

**Suppl.Table.1**: MetaCardis Cohort characteristics for all, males and females

**Suppl.Table.2.** SHAP values from XGBOOST models

**Suppl.Table.3.** CLSA cohort overview.

**Suppl.Table.4**. MetaCardis results Summary

**Suppl.Table.5**. CLSA CKM outcome correction with a metabolome-derived GFR

**Suppl.Table.6**. CLSA all-cause mortality sensitivity analyses

**Suppl.Table.7**. CLSA results Summary

**Suppl.Table.8.** GCKD cohort characteristics

**Suppl.Table.9.** GCKD sensitivity analyses with eGFRcr-cys

**Suppl.Table.10.** GCKD event time censoring

**Suppl.Table.11.** GCKD subgroup sensitivity analyses

**Suppl.Table.12.** GCKD results summary

**Suppl.Table.13.** Summary of metabolomics methods across cohorts.

## Disclaimer

The opinions expressed in this manuscript are the author’s own and do not reflect the views of the Canadian Longitudinal Study on Aging.

## Acknowledgements

The authors would like to thank the MetaCardis, CLSA and GCKD study participants as well as clinical and non-clinical staff at all recruitment centres across the three studies. MetaCardis was funded by European Union’s Seventh Framework Program for research (FP7/2007-2013), technological development, and demonstration under grant agreement HEALTH-F4-2012-305312 (METACARDIS), a Transatlantic Networks of Excellence Award from the Leducq Foundation (17CVD01) and by the NIHR Imperial Biomedical Research Centre. Assistance Publique-Hôpitaux de Paris (AP-HP) is the promoter of clinical investigation (MetaCardis). The GCKD study was supported by the BMBF (FKZ 01ER 0804, 01ER 0818, 01ER 0819, 01ER 0820 and 01ER 0821) and the KfH Foundation for Preventive Medicine. Plasma metabolomics were partly funded as part of the Innovative Medicines Initiative 2 Joint Undertaking (JU) under grant agreement 115974. The JU had received support from the European Union’s Horizon 2020 research and innovation program and the EFPIA and the JDRF. Unregistered grants to support the GCKD study were provided by corporate sponsors (listed at https://gckd.org). We are grateful for the willingness of the patients to participate in the GCKD study. The enormous effort of the study personnel of the various regional centers is highly appreciated. We thank the large number of nephrologists who provide routine care for the patients and collaborate with the GCKD study. The GCKD investigators are listed in the Supplementary Note. The authors also acknowledge that this research was made possible using the data/biospecimens collected by the Canadian Longitudinal Study on Aging (CLSA). Funding for the Canadian Longitudinal Study on Aging (CLSA) is provided by the Government of Canada through the Canadian Institutes of Health Research (CIHR) under grant reference: LSA 94473 and the Canada Foundation for Innovation, as well as the following provinces, Newfoundland, Nova Scotia, Quebec, Ontario, Manitoba, Alberta, and British Columbia. This research has been conducted using the CLSA metabolomics data version 2, CLSA Comprehensive baseline dataset (v7), Comprehensive follow-up 1 dataset (v5), CLSA participant status data under Application Number 2104039. The CLSA is led by Drs. Parminder Raina, Christina Wolfson and Susan Kirkland. The authors gratefully acknowledge the time and commitment of the CLSA participants, without whom this research would not be possible. This research was also conducted as part of the CNRS–Imperial-ULille International Research Project in Integrative Metabolism “METABO-LIC”. This research was also partially funded by a Career Development Award from the Medical Research Council (Grant No. MR/Y010051/1) to P.A. M.E-D. acknowledges funding support from the EU IMMEDIATE consortium and UKRI Innovate UK under contract number 101095556 and by the National Institute for Health Research (NIHR) Imperial Biomedical Research Centre, grants from Guts UK (DG201808) and Diabetes UK (19/0006059), and a Medical Research Council grant to M.E-D. and P.F. (MR/X010155/1). The work of A.K. and P.S. was supported by the DFG Project-ID 431984000 - SFB 1453. The work of E.B. and P.S. was supported by the German Research Foundation (DFG, SE 2407/3-1). K.Cle. also thank the French National Agency of Research ANR (deepIntegromics project ANR-21-CE45-0030-02 and Nutrim-Check project ANR-23-CE17-0020) and the Fondation de France and Fondation Larue (MetaCardis evolution), as well as the European Innovation Council (EIC) Pathfinder Challenge project NUTRIMMUNE (Grant Agreement No. 101162457) and the Food Systems, Microbiomes and Health Priority Research Programme and Equipment (PEPR SAMS) funded by the French Government through France 2030, managed by the Agence Nationale de la Recherche (ANR) (ANR-24-PESA-0010). J.M.N. is supported by the Berta-Ottenstein-Programme for Clinician Scientists, Faculty of Medicine, University of Freiburg and the Deutsche Forschungsgemeinschaft (DFG, German Research Foundation) Project-ID 499552394 – SFB 1597. R.C. is the recipient of the Walter-Benjamin-Fellowship from the German Research Association (DFG), project number: 462524713 and also funded by the DFG under Germanýs Excellence Strategy – EXC-3105/1 – 533765739. The Novo Nordisk Foundation Center for Basic Metabolic Research is an independent research institution at the University of Copenhagen, partially funded by an unrestricted donation from the Novo Nordisk Foundation (NNF18CC0034900).

## Author Contributions Statement

P.A. developed the present study concept and protocol with input from M.-E.D., K.Cle., O.P., M.S., A.K., J.M.N. and R.C. K.Cle. (Coordinator of the MetaCardis EU consortium and principal clinical investigator), M.-ED., S.D.E, O.P., P.B., M.S., J.R., J.N., J.B.N. and F.B. conceived the overall objectives and study design of the MetaCardis consortium. MetaCardis cohort recruitment, phenotyping and lifestyle: J.A.-W., R.C., T.N., L.K., H.V., T.H., J.-M.O., and M.B. and supervised by K.Cle., M.S., and O.P. Data curation: R.C., S.K.F., J.A.-W., and T.N. Microbiota profiling: S.K.F.-S., S.V.S., G.F., J.R., P.B., S.D.E. Serum and urine metabolome profiling: A.M., J.C., E.M., F.P.-C, R.P.-T., I.C.-D., P.F. Bioinformatics and statistical analyses (MetaCardis): P.A., R.C., S.K.F., G.F, S.V.S., A.M., K.Ch., C.R., C.P., P.B.L., E.B., E.L.C. and J.-D.Z. (CLSA): M.J., K.Ch., M.L. and P.R. (GCKD): J.M.N., E.B., P.S., K.-U.E. and A.K. Modeling of phenotypic data: P.A. The manuscript was drafted by P.A. with input from J.M.N., R.C., A.K., K-U.E., F.B., S.D.E., M.S., O.P., K.Cle. and M.-E.D. All authors participated in project development, discussion of results and revision of the article and approved the final version for submission.

## MetaCardis Consortium

The MetaCardis consortium †: Rohia Alili, Renato Alves, Ehm Astrid Andersson Galijatovic, Olivier Barthelemy, Jean-Philippe Bastard, Jean-Paul Batisse, Randa Bittar, Matthias Blüher, Frederic Bosquet, Olivier Bourron, Mickael Camus, Cecile Ciangura, Jean-Philippe Collet, Arne Dietrich, Morad Djebbar, Angélique Doré, Line Engelbrechtsen, Leopold Fezeu, Sebastien Fromentin, Nathalie Galleron, Marianne Graine, Caroline Grünemann, Agnes Hartemann, Bolette Hartmann, Gerard Helft, Malene Hornbak, Charlotte Kranepuhl, Julien Chilloux, Antonis Myridakis, Lesley Hoyles, Richard Isnard, Sophie Jaqueminet, Niklas Rye Jørgensen, Hanna Julienne, Johanne Justesen, Judith Kammer, Nikolaj Karup, Lars Køber, Michael Kuhn, Véronique Lejard, Ivica Letunic, Florence Levenez, Lajos Marko, Laura Martinez-Gili, Robin Massey, Nicolas Maziers, Lucas Moitinho-Silva, Gilles Montalescot, Ana Luisa Neves, Michael Olanipekun, Laetitia Pasero Le Pavin, Luis Pedro Coelho, Nicolas Pons, Benoit Quinquis, Andrea Rodriguez-Martinez, Christine Rouault, Hugo Roume, Sebastien Schmidt, Tatjana Schütz, Lucas Silva, Johanne Silvain, Mathilde Svendstrup, Thierry Vanduyvenboden, Stefanie Walther.

## GCKD Consortium

Kai-Uwe Eckardt, Heike Meiselbach, Markus P. Schneider, Mario Schiffer, Hans-Ulrich Prokosch, Barbara Bärthlein, Andreas Beck, André Reis, Arif B. Ekici, Susanne Becker, Ulrike Alberth-Schmidt, Anke Weigel, Sabine Marschall, Gerd Walz, Anna Köttgen, Ulla T. Schultheiß, Fruzsina Kotsis, Simone Meder, Erna Mitsch, Ursula Reinhard, Jürgen Floege, Rafael Kramann, Turgay Saritas, Elke Schaeffner, Seema Baid-Agrawal, Kerstin Theisen, Kai Schmidt-Ott, Martin Zeier, Claudia Sommerer, Gunter Wolf, Martin Busch, Thomas Sitter, Christoph Wanner, Vera Krane, Britta Bauer, Florian Kronenberg, Julia Raschenberger, Barbara Kollerits, Lukas Forer, Sebastian Schönherr, Hansi Weissensteiner, Peter Oefner, Wolfram Gronwald, Matthias Schmid, Jennifer Nadal.

