## Supplemental Figures for "Metabolome-derived kidney function scores as prognostic readouts of cardiovascular-kidney-metabolic health"

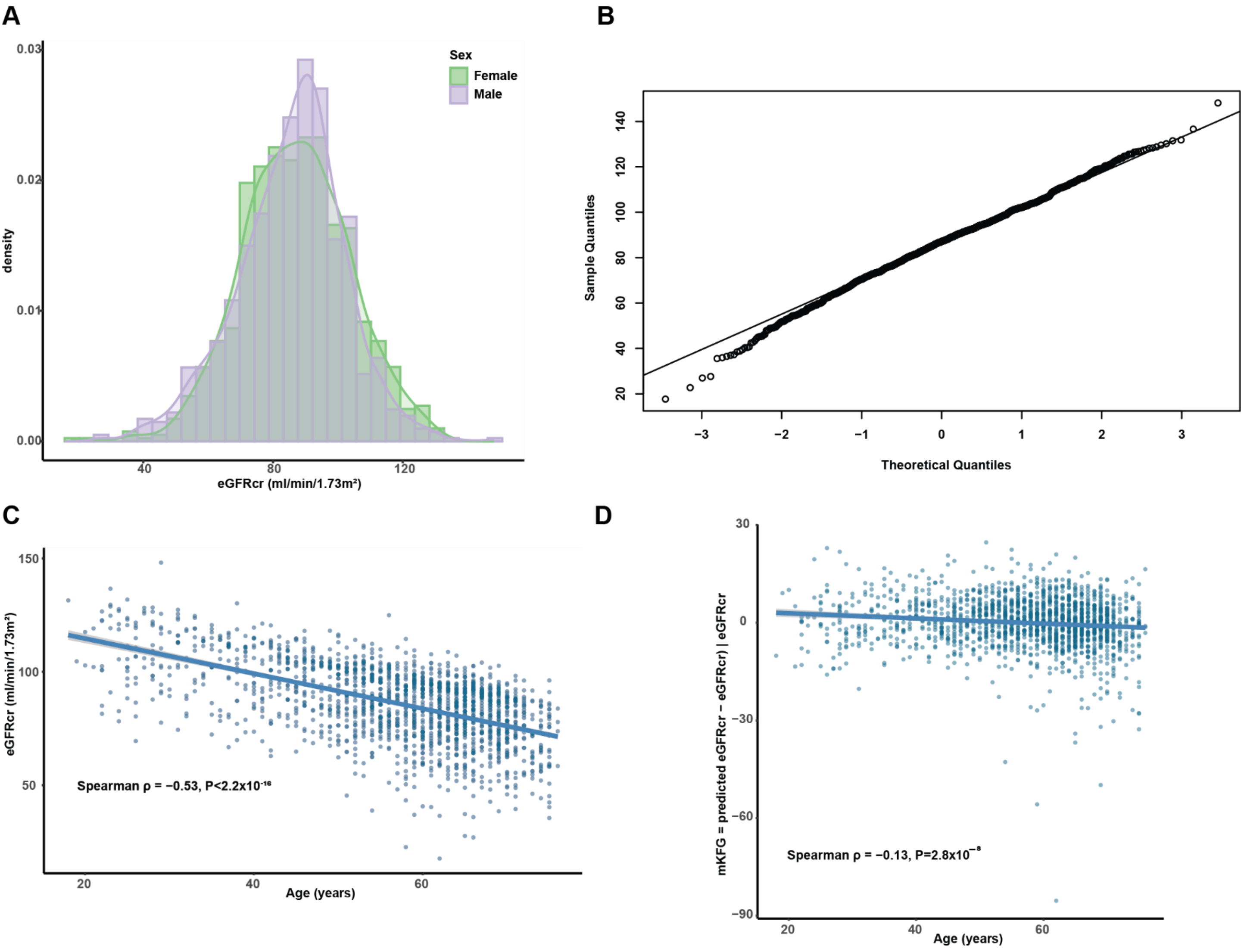
A
Sex
Female
Male
density
0.03
0.02
0.01
0.00
40
80
120
eGFRcr (ml/min/1.73m²)
B
Sample Quantiles
20
40
60
80
100
120
140
−3
−2
−1
0
1
2
3
Theoretical Quantiles
C
eGFRcr (ml/min/1.73m²)
150
100
50
Spearman ρ = −0.53, P<2.2x10⁻¹⁶
20
40
60
Age (years)
D
mKFG = predicted eGFRcr − eGFRcr) | eGFRcr
30
0
−30
−60
−90
Spearman ρ = −0.13, P=2.8x10⁻⁸
20
40
60
Age (years)

Suppl.FIG.1

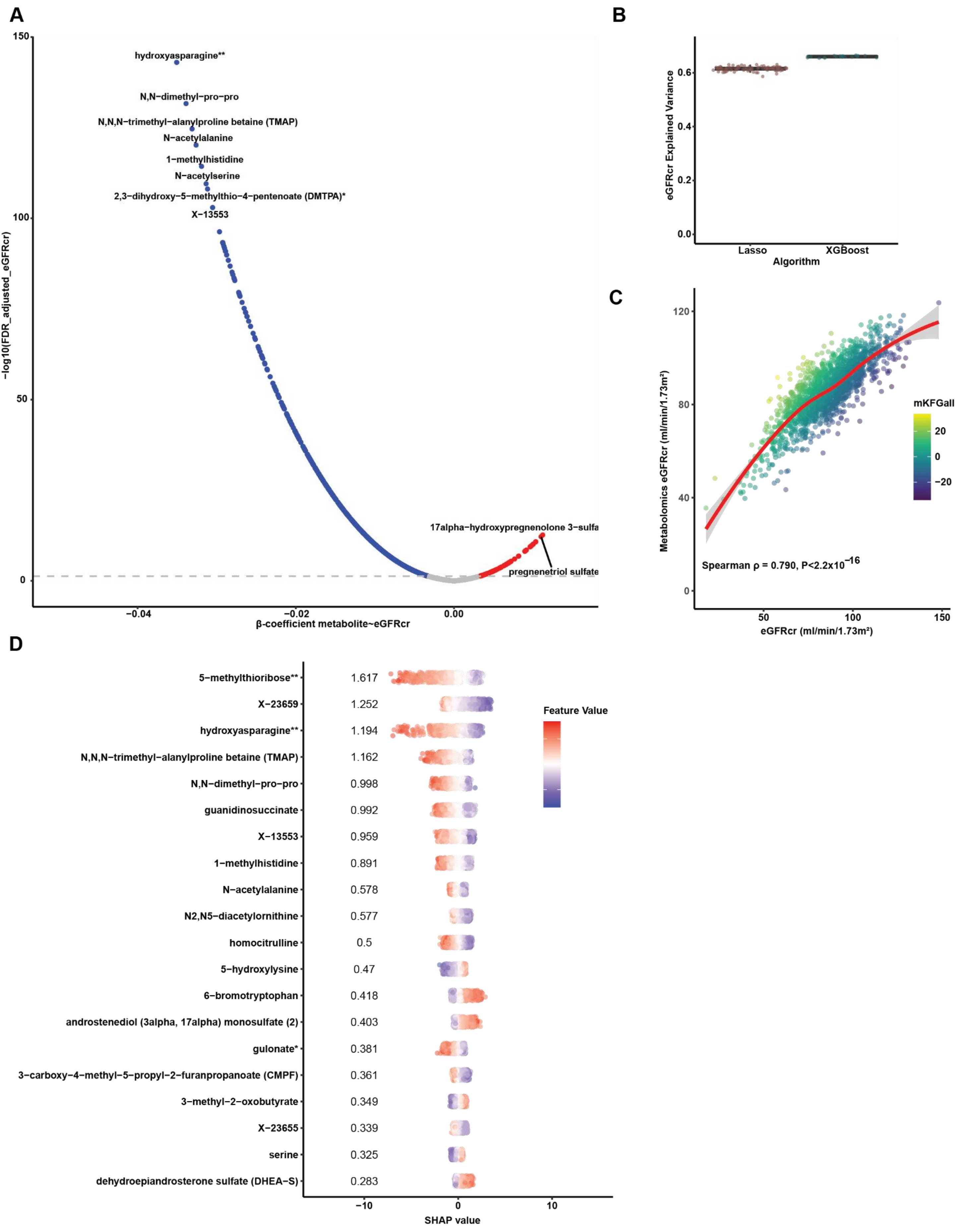
A
hydroxyasparagine**
N,N−dimethyl−pro−pro
N,N,N−trimethyl−alanylproline betaine (TMAP)
N−acetylalanine
1−methylhistidine
N−acetylserine
2,3−dihydroxy−5−methylthio−4−pentenoate (DMTPA)*
X−13553
17alpha−hydroxypregnenolone 3−sulfa
pregnenetriol sulfate
−log10(FDR_adjusted_eGFRcr)
β-coefficient metabolite~eGFRcr
B
eGFRcr Explained Variance
Lasso
XGBoost
Algorithm
C
Metabolomics eGFRcr (ml/min/1.73m²)
eGFRcr (ml/min/1.73m²)
mKFGall
Spearman ρ = 0.790, P<2.2x10^−16
D
5−methylthioribose** 1.617
X−23659 1.252
hydroxyasparagine** 1.194
N,N,N−trimethyl−alanylproline betaine (TMAP) 1.162
N,N−dimethyl−pro−pro 0.998
guanidinosuccinate 0.992
X−13553 0.959
1−methylhistidine 0.891
N−acetylalanine 0.578
N2,N5−diacetylornithine 0.577
homocitrulline 0.5
5−hydroxylysine 0.47
6−bromotryptophan 0.418
androstenediol (3alpha, 17alpha) monosulfate (2) 0.403
gulonate* 0.381
3−carboxy−4−methyl−5−propyl−2−furanpropanoate (CMPF) 0.361
3−methyl−2−oxobutyrate 0.349
X−23655 0.339
serine 0.325
dehydroepiandrosterone sulfate (DHEA−S) 0.283
Feature Value
SHAP value

Suppl.FIG.2

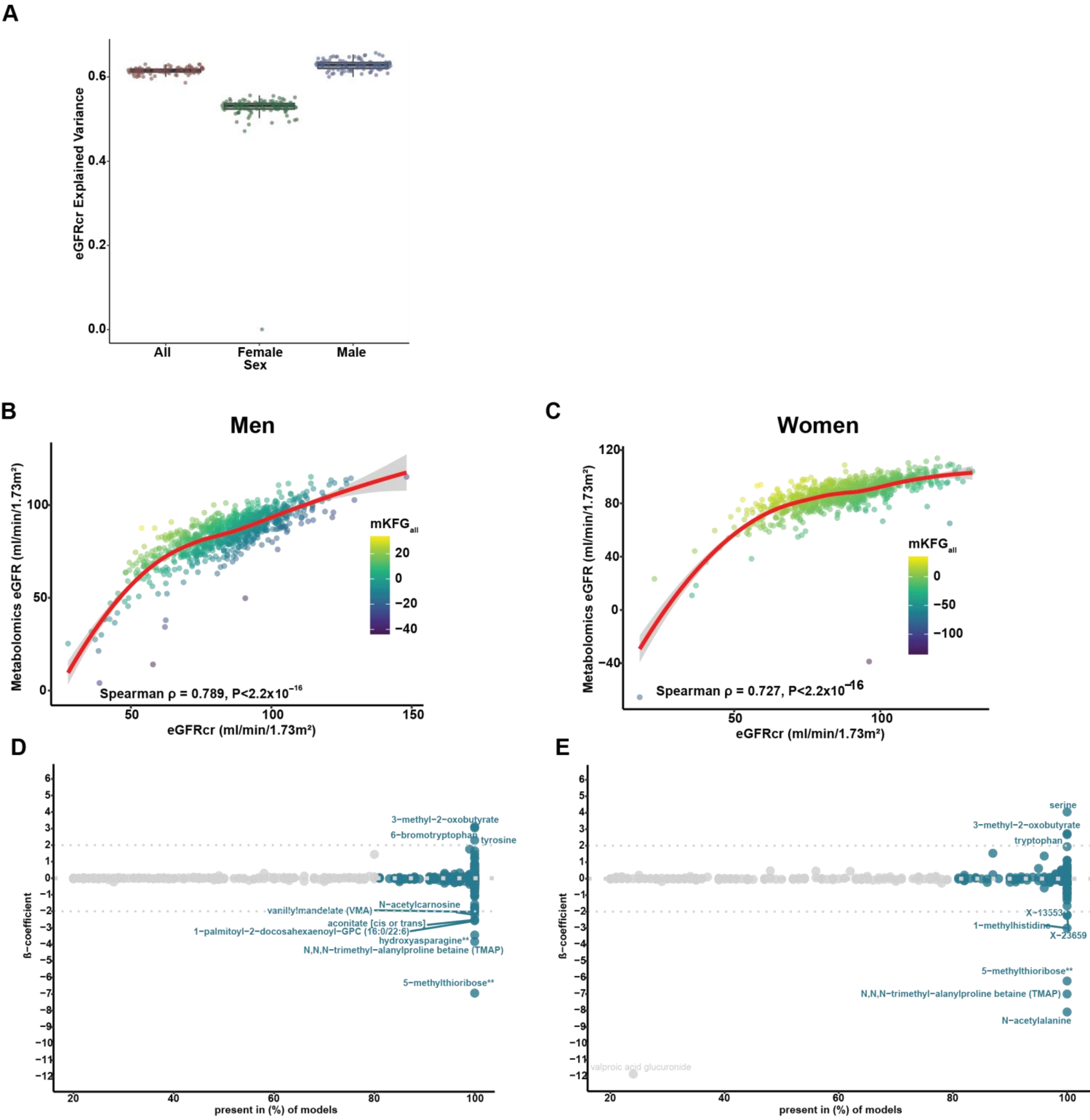
A
eGFRcr Explained Variance
0.6
0.4
0.2
0.0
All
Female
Male
Sex
B
Men
Metabolomics eGFR (ml/min/1.73m²)
mKFG$_{all}$
20
0
−20
−40
Spearman ρ = 0.789, P<2.2x10$^{-16}$
eGFRcr (ml/min/1.73m²)
C
Women
Metabolomics eGFR (ml/min/1.73m²)
mKFG$_{all}$
0
−50
−100
Spearman ρ = 0.727, P<2.2x10$^{-16}$
eGFRcr (ml/min/1.73m²)
D
ß−coefficient
3−methyl−2−oxobutyrate
6−bromotryptophan
tyrosine
N−acetylcarnosine
vanillylmandelate (VMA)
aconitate [cis or trans]
1−palmitoyl−2−docosahexaenoyl−GPC (16:0/22:6)
hydroxyasparagine**
N,N,N−trimethyl−alanylproline betaine (TMAP)
5−methylthioribose**
present in (%) of models
E
ß−coefficient
serine
3−methyl−2−oxobutyrate
tryptophan
X−13553
1−methylhistidine
X−23659
5−methylthioribose**
N,N,N−trimethyl−alanylproline betaine (TMAP)
N−acetylalanine
valproic acid glucuronide
present in (%) of models

**Suppl.FIG.3**

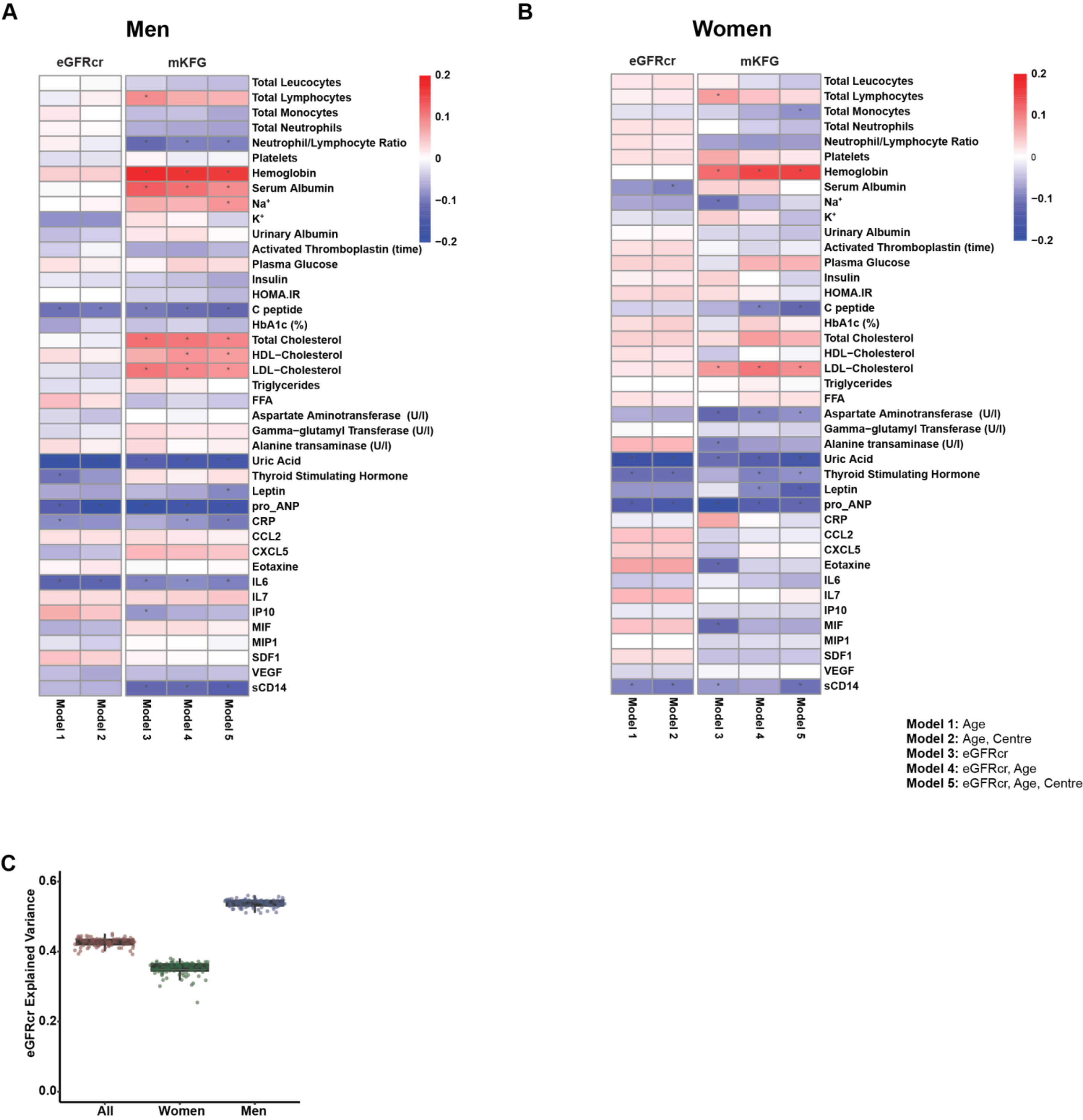
A
Men
eGFRcr
mKFG
Total Leucocytes
Total Lymphocytes
Total Monocytes
Total Neutrophils
Neutrophil/Lymphocyte Ratio
Platelets
Hemoglobin
Serum Albumin
Na+
K+
Urinary Albumin
Activated Thromboplastin (time)
Plasma Glucose
Insulin
HOMA.IR
C peptide
HbA1c (%)
Total Cholesterol
HDL−Cholesterol
LDL−Cholesterol
Triglycerides
FFA
Aspartate Aminotransferase (U/l)
Gamma−glutamyl Transferase (U/l)
Alanine transaminase (U/l)
Uric Acid
Thyroid Stimulating Hormone
Leptin
pro_ANP
CRP
CCL2
CXCL5
Eotaxine
IL6
IL7
IP10
MIF
MIP1
SDF1
VEGF
sCD14
Model 1
Model 2
Model 3
Model 4
Model 5
0.2
0.1
0
−0.1
−0.2
B
Women
eGFRcr
mKFG
Total Leucocytes
Total Lymphocytes
Total Monocytes
Total Neutrophils
Neutrophil/Lymphocyte Ratio
Platelets
Hemoglobin
Serum Albumin
Na+
K+
Urinary Albumin
Activated Thromboplastin (time)
Plasma Glucose
Insulin
HOMA.IR
C peptide
HbA1c (%)
Total Cholesterol
HDL−Cholesterol
LDL−Cholesterol
Triglycerides
FFA
Aspartate Aminotransferase (U/l)
Gamma−glutamyl Transferase (U/l)
Alanine transaminase (U/l)
Uric Acid
Thyroid Stimulating Hormone
Leptin
pro_ANP
CRP
CCL2
CXCL5
Eotaxine
IL6
IL7
IP10
MIF
MIP1
SDF1
VEGF
sCD14
Model 1
Model 2
Model 3
Model 4
Model 5
0.2
0.1
0
−0.1
−0.2
Model 1: Age
Model 2: Age, Centre
Model 3: eGFRcr
Model 4: eGFRcr, Age
Model 5: eGFRcr, Age, Centre
C
eGFRcr Explained Variance
0.6
0.4
0.2
0.0
All
Women
Men

Suppl.FIG.4

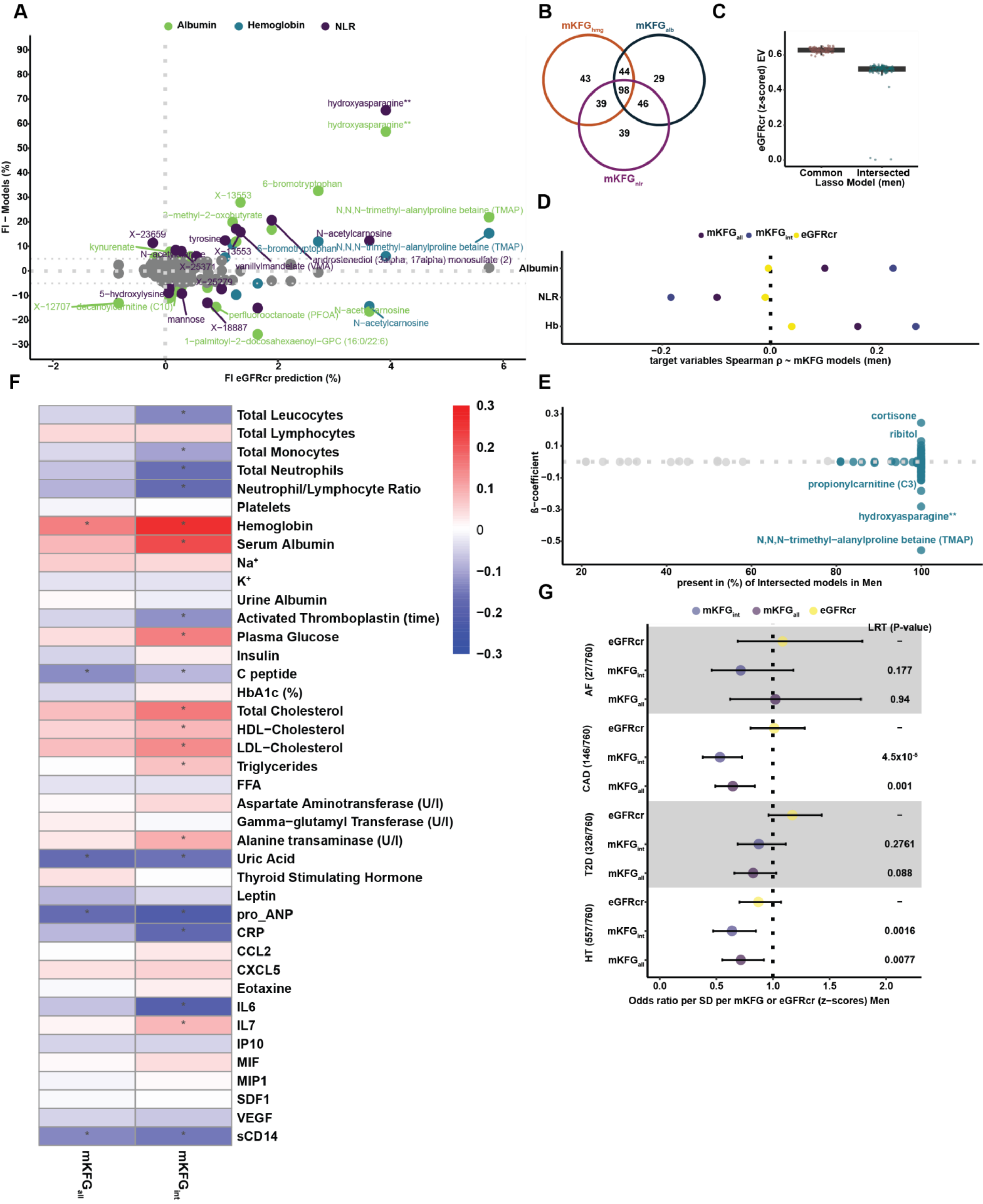
A
Albumin
Hemoglobin
NLR
FI − Models (%)
FI eGFRcr prediction (%)
hydroxyasparagine**
hydroxyasparagine**
6−bromotryptophan
X−13553
N,N,N−trimethyl−alanylproline betaine (TMAP)
3−methyl−2−oxobutyrate
X−23659
N−acetylcarnosine
tyrosine
kynurenate
6−bromotryptophan
N,N,N−trimethyl−alanylproline betaine (TMAP)
androstenediol (3alpha, 17alpha) monosulfate (2)
vanillylmandelate (VMA)
X−25371
X−25279
5−hydroxylysine
X−12707
decanoylcarnitine (C10)
perfluorooctanoate (PFOA)
N−acetylcarnosine
N−acetylcarnosine
mannose
X−18887
1−palmitoyl−2−docosahexaenoyl−GPC (16:0/22:6)
B
mKFG_hmg
mKFG_alb
mKFG_nlr
43
44
29
98
39
46
39
C
eGFRcr (z−scored) EV
Common
Intersected
Lasso Model (men)
D
mKFG_all
mKFG_int
eGFRcr
Albumin
NLR
Hb
target variables Spearman ρ ~ mKFG models (men)
E
ß−coefficient
cortisone
ribitol
propionylcarnitine (C3)
hydroxyasparagine**
N,N,N−trimethyl−alanylproline betaine (TMAP)
present in (%) of Intersected models in Men
F
Total Leucocytes
Total Lymphocytes
Total Monocytes
Total Neutrophils
Neutrophil/Lymphocyte Ratio
Platelets
Hemoglobin
Serum Albumin
Na+
K+
Urine Albumin
Activated Thromboplastin (time)
Plasma Glucose
Insulin
C peptide
HbA1c (%)
Total Cholesterol
HDL−Cholesterol
LDL−Cholesterol
Triglycerides
FFA
Aspartate Aminotransferase (U/l)
Gamma−glutamyl Transferase (U/l)
Alanine transaminase (U/l)
Uric Acid
Thyroid Stimulating Hormone
Leptin
pro_ANP
CRP
CCL2
CXCL5
Eotaxine
IL6
IL7
IP10
MIF
MIP1
SDF1
VEGF
sCD14
mKFG_all
mKFG_int
G
mKFG_int
mKFG_all
eGFRcr
LRT (P-value)
AF (27/760)
CAD (146/760)
T2D (326/760)
HT (557/760)
0.177
0.94
4.5x10-5
0.001
0.2761
0.088
0.0016
0.0077
Odds ratio per SD per mKFG or eGFRcr (z−scores) Men

Suppl.FIG.5

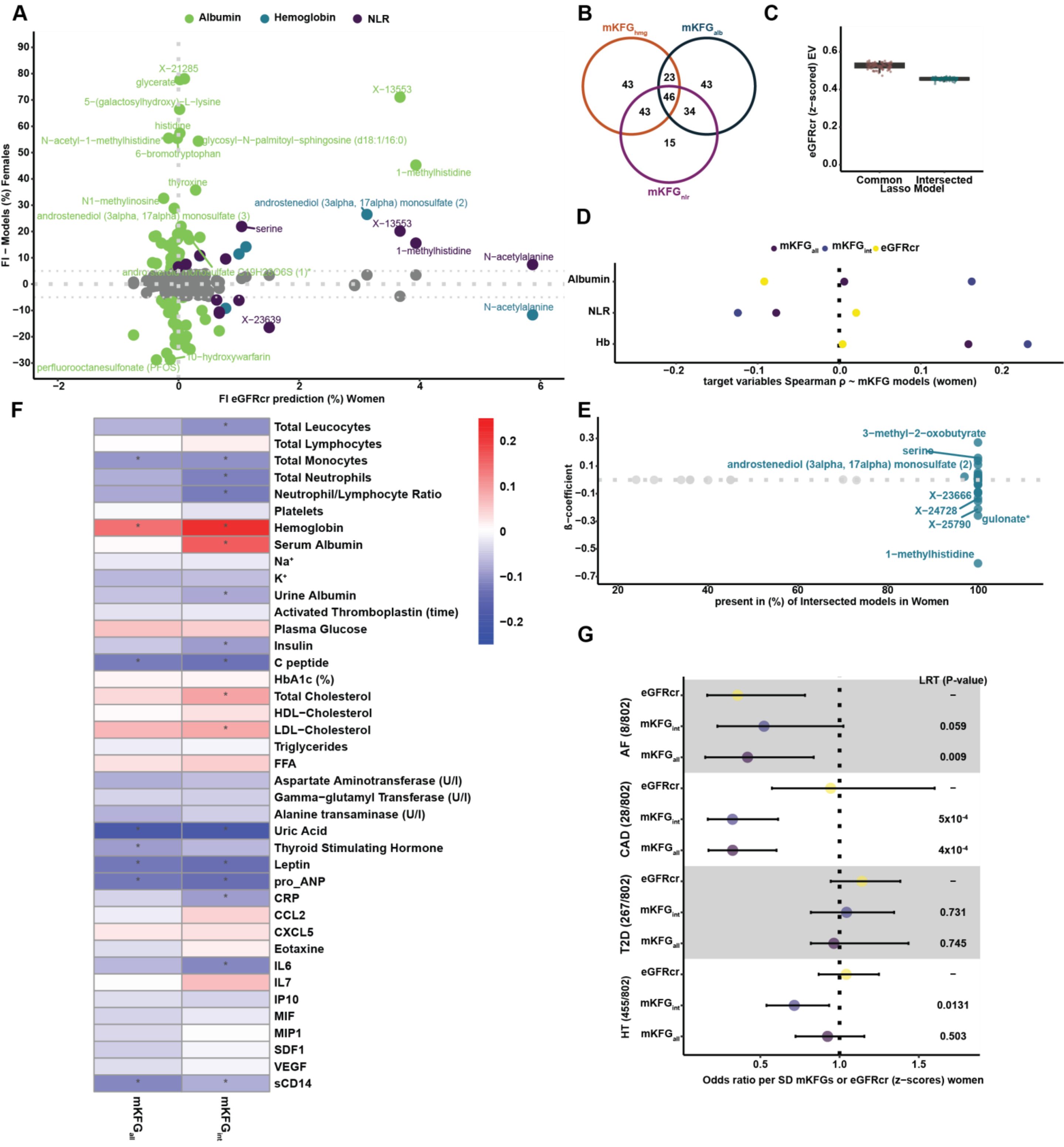

A
Albumin
Hemoglobin
NLR
FI – Models (%) Females
FI eGFRcr prediction (%) Women
X−21285
glycerate
5−(galactosylhydroxy)−L−lysine
histidine
N−acetyl−1−methylhistidine
glycosyl−N−palmitoyl−sphingosine (d18:1/16:0)
6−bromotryptophan
X−13553
1−methylhistidine
thyroxine
N1−methylinosine
androstenediol (3alpha, 17alpha) monosulfate (2)
androstenediol (3alpha, 17alpha) monosulfate (3)
serine
N−acetylalanine
X−23639
10−hydroxywarfarin
perfluorooctanesulfonate (PFOS)
B
mKFG hmg
mKFG alb
mKFG nlr
C
eGFRcr (z−scored) EV
Common
Intersected
Lasso Model
D
mKFG all
mKFG int
eGFRcr
Albumin
NLR
Hb
target variables Spearman ρ ~ mKFG models (women)
E
ß−coefficient
present in (%) of Intersected models in Women
3−methyl−2−oxobutyrate
serine
androstenediol (3alpha, 17alpha) monosulfate (2)
X−23666
X−24728
X−25790
gulonate*
1−methylhistidine
F
Total Leucocytes
Total Lymphocytes
Total Monocytes
Total Neutrophils
Neutrophil/Lymphocyte Ratio
Platelets
Hemoglobin
Serum Albumin
Na+
K+
Urine Albumin
Activated Thromboplastin (time)
Plasma Glucose
Insulin
C peptide
HbA1c (%)
Total Cholesterol
HDL−Cholesterol
LDL−Cholesterol
Triglycerides
FFA
Aspartate Aminotransferase (U/l)
Gamma−glutamyl Transferase (U/l)
Alanine transaminase (U/l)
Uric Acid
Thyroid Stimulating Hormone
Leptin
pro_ANP
CRP
CCL2
CXCL5
Eotaxine
IL6
IL7
IP10
MIF
MIP1
SDF1
VEGF
sCD14
mKFG all
mKFG int
G
LRT (P-value)
AF (8/802)
CAD (28/802)
T2D (267/802)
HT (455/802)
eGFRcr
mKFG int
mKFG all
0.059
0.009
5x10-4
4x10-4
0.731
0.745
0.0131
0.503
Odds ratio per SD mKFGs or eGFRcr (z−scores) women


Suppl.FIG.6

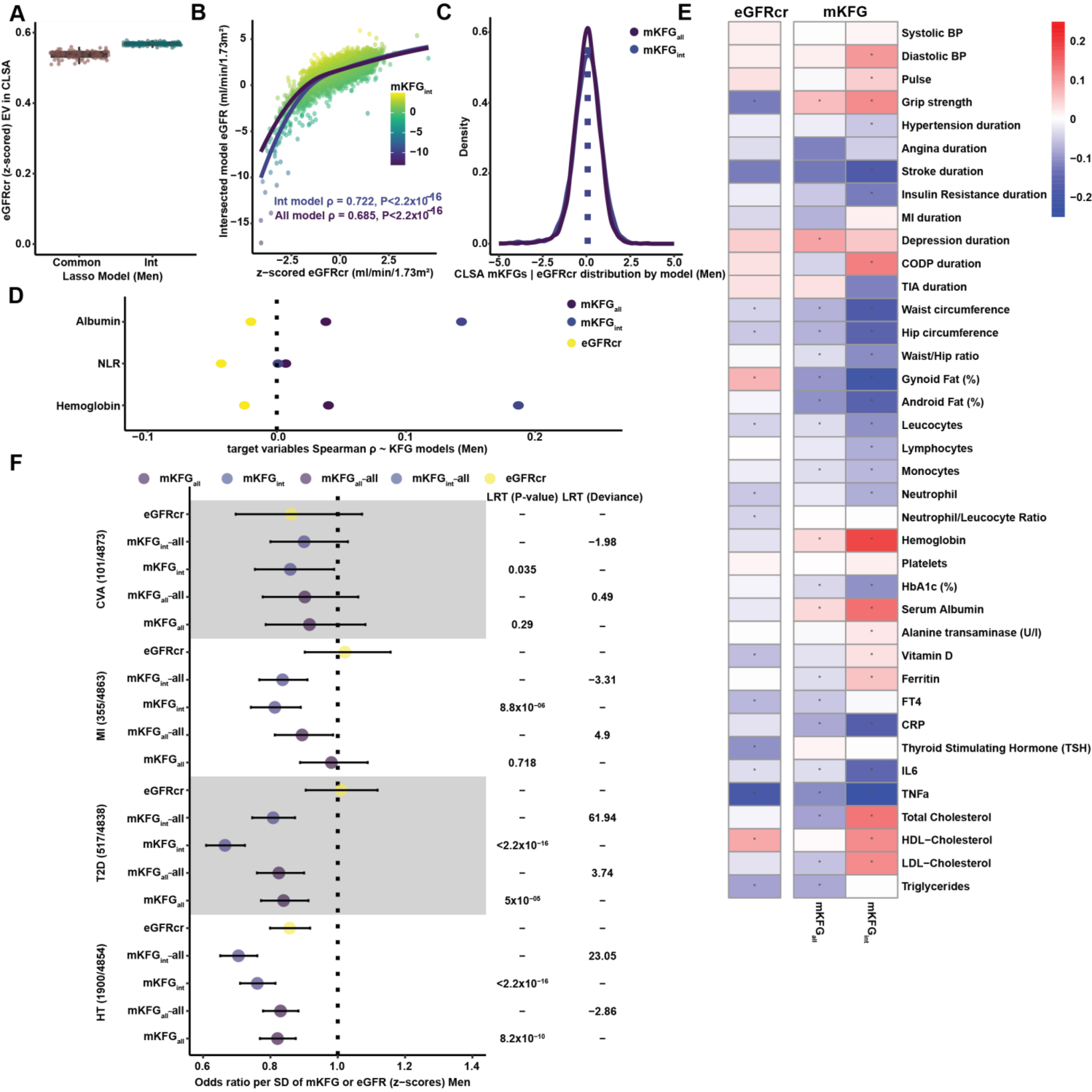


| Outcome | Model | LRT (P-value) | LRT (Deviance) |
|---|---|---|---|
| CVA (101/4873) | eGFRcr | – | – |
| | $mKFG_{int}$-all | – | −1.98 |
| | $mKFG_{int}$ | 0.035 | – |
| | $mKFG_{all}$-all | – | 0.49 |
| | $mKFG_{all}$ | 0.29 | – |
| MI (355/4863) | eGFRcr | – | – |
| | $mKFG_{int}$-all | – | −3.31 |
| | $mKFG_{int}$ | $8.8\times10^{-06}$ | – |
| | $mKFG_{all}$-all | – | 4.9 |
| | $mKFG_{all}$ | 0.718 | – |
| T2D (517/4838) | eGFRcr | – | – |
| | $mKFG_{int}$-all | – | 61.94 |
| | $mKFG_{int}$ | $<2.2\times10^{-16}$ | – |
| | $mKFG_{all}$-all | – | 3.74 |
| | $mKFG_{all}$ | $5\times10^{-05}$ | – |
| HT (1900/4854) | eGFRcr | – | – |
| | $mKFG_{int}$-all | – | 23.05 |
| | $mKFG_{int}$ | $<2.2\times10^{-16}$ | – |
| | $mKFG_{all}$-all | – | −2.86 |
| | $mKFG_{all}$ | $8.2\times10^{-10}$ | – |

Suppl.FIG.7

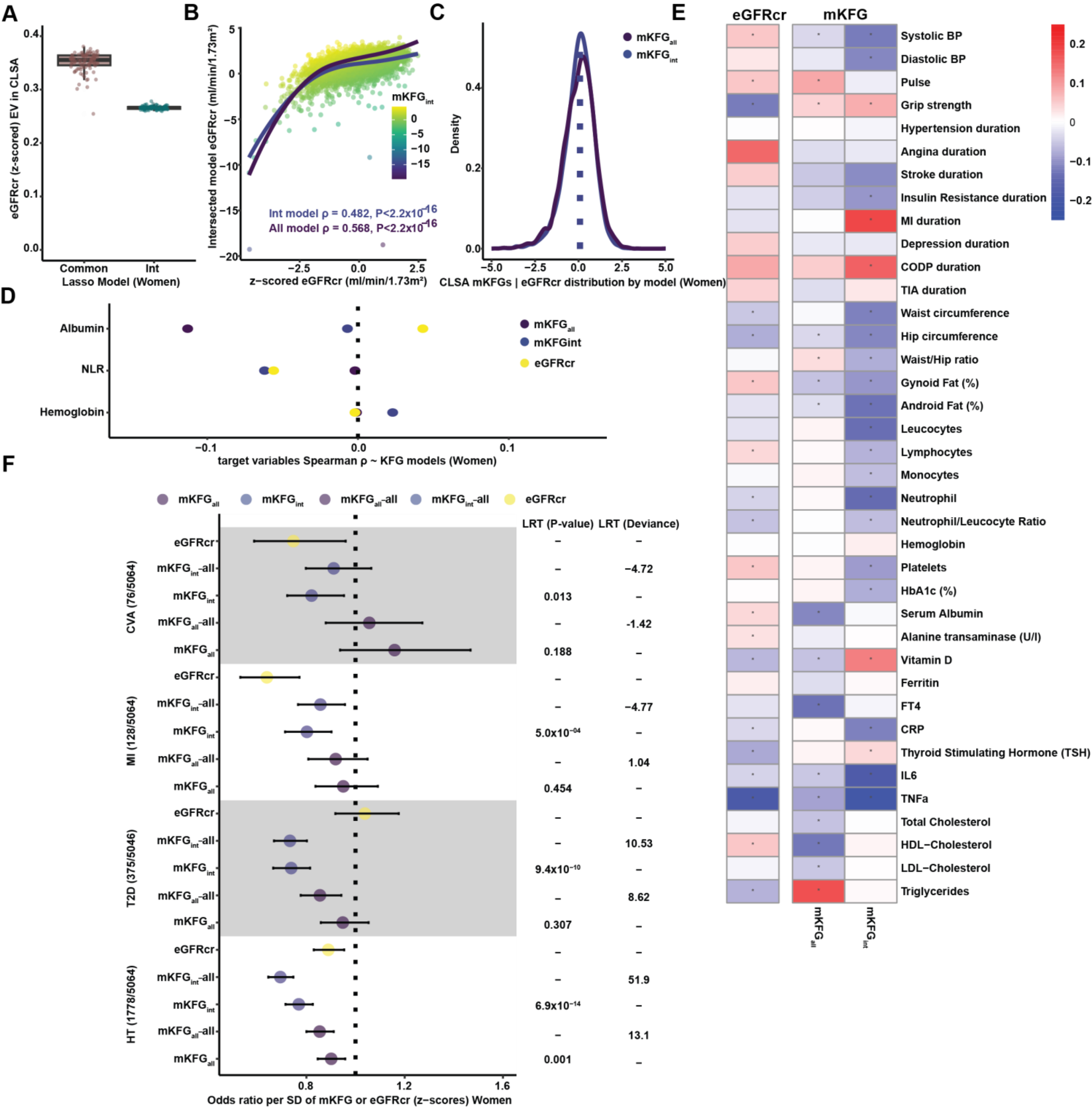
A
eGFRcr (z-scored) EV in CLSA
Common
Int
Lasso Model (Women)
B
Intersected model eGFRcr (ml/min/1.73m²)
z-scored eGFRcr (ml/min/1.73m²)
mKFGint
Int model ρ = 0.482, P<2.2x10-16
All model ρ = 0.568, P<2.2x10-16
C
Density
mKFGall
mKFGint
CLSA mKFGs | eGFRcr distribution by model (Women)
D
Albumin
NLR
Hemoglobin
mKFGall
mKFGint
eGFRcr
target variables Spearman ρ ~ KFG models (Women)
E
eGFRcr
mKFG
Systolic BP
Diastolic BP
Pulse
Grip strength
Hypertension duration
Angina duration
Stroke duration
Insulin Resistance duration
MI duration
Depression duration
CODP duration
TIA duration
Waist circumference
Hip circumference
Waist/Hip ratio
Gynoid Fat (%)
Android Fat (%)
Leucocytes
Lymphocytes
Monocytes
Neutrophil
Neutrophil/Leucocyte Ratio
Hemoglobin
Platelets
HbA1c (%)
Serum Albumin
Alanine transaminase (U/l)
Vitamin D
Ferritin
FT4
CRP
Thyroid Stimulating Hormone (TSH)
IL6
TNFa
Total Cholesterol
HDL−Cholesterol
LDL−Cholesterol
Triglycerides
mKFGall
mKFGint
F
mKFGall
mKFGint
mKFGall-all
mKFGint-all
eGFRcr
LRT (P-value)
LRT (Deviance)
CVA (76/5064)
MI (128/5064)
T2D (375/5046)
HT (1778/5064)
0.013
0.188
−4.72
-1.42
5.0x10−04
0.454
−4.77
1.04
9.4x10−10
0.307
10.53
8.62
6.9x10−14
0.001
51.9
13.1
Odds ratio per SD of mKFG or eGFRcr (z−scores) Women

Suppl.FIG.8

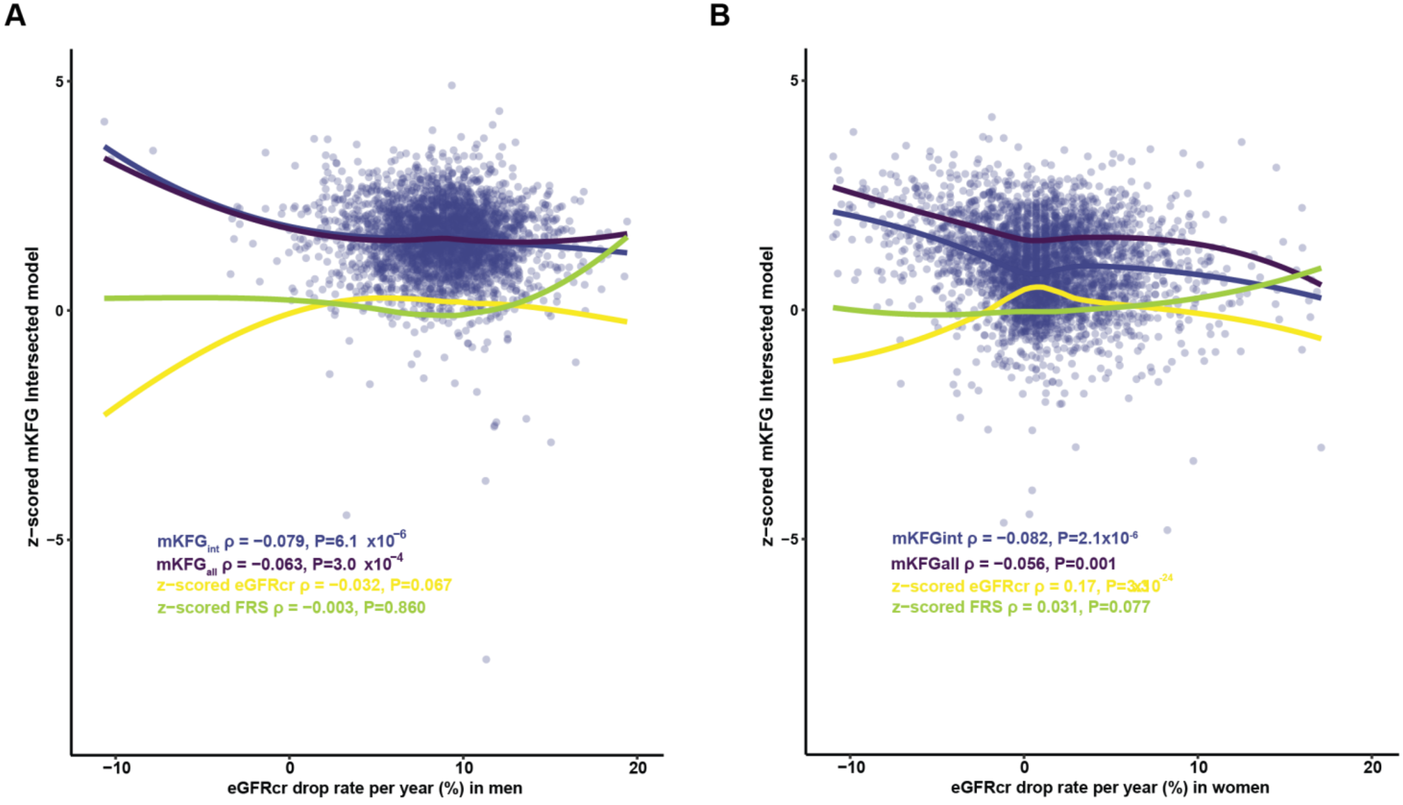

A
B
z−scored mKFG Intersected model
5
0
−5
−10
10
20
eGFRcr drop rate per year (%) in men
eGFRcr drop rate per year (%) in women
mKFGint ρ = −0.079, P=6.1 x10−6
mKFGall ρ = −0.063, P=3.0 x10−4
z−scored eGFRcr ρ = −0.032, P=0.067
z−scored FRS ρ = −0.003, P=0.860
mKFGint ρ = −0.082, P=2.1x10-6
mKFGall ρ = −0.056, P=0.001
z−scored eGFRcr ρ = 0.17, P=3x10-24
z−scored FRS ρ = 0.031, P=0.077


Suppl.FIG.9

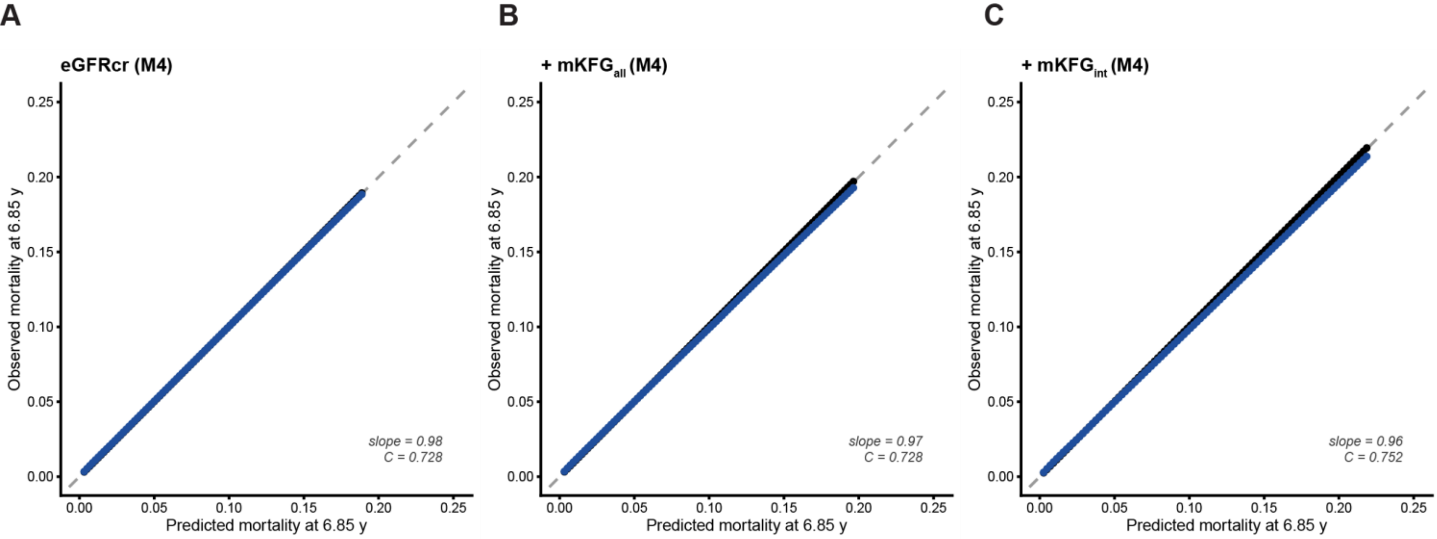
A
eGFRcr (M4)
B
+ mKFG_all (M4)
C
+ mKFG_int (M4)
Observed mortality at 6.85 y
Predicted mortality at 6.85 y
slope = 0.98
C = 0.728
slope = 0.97
C = 0.728
slope = 0.96
C = 0.752
0.00
0.05
0.10
0.15
0.20
0.25

Suppl.FIG.10

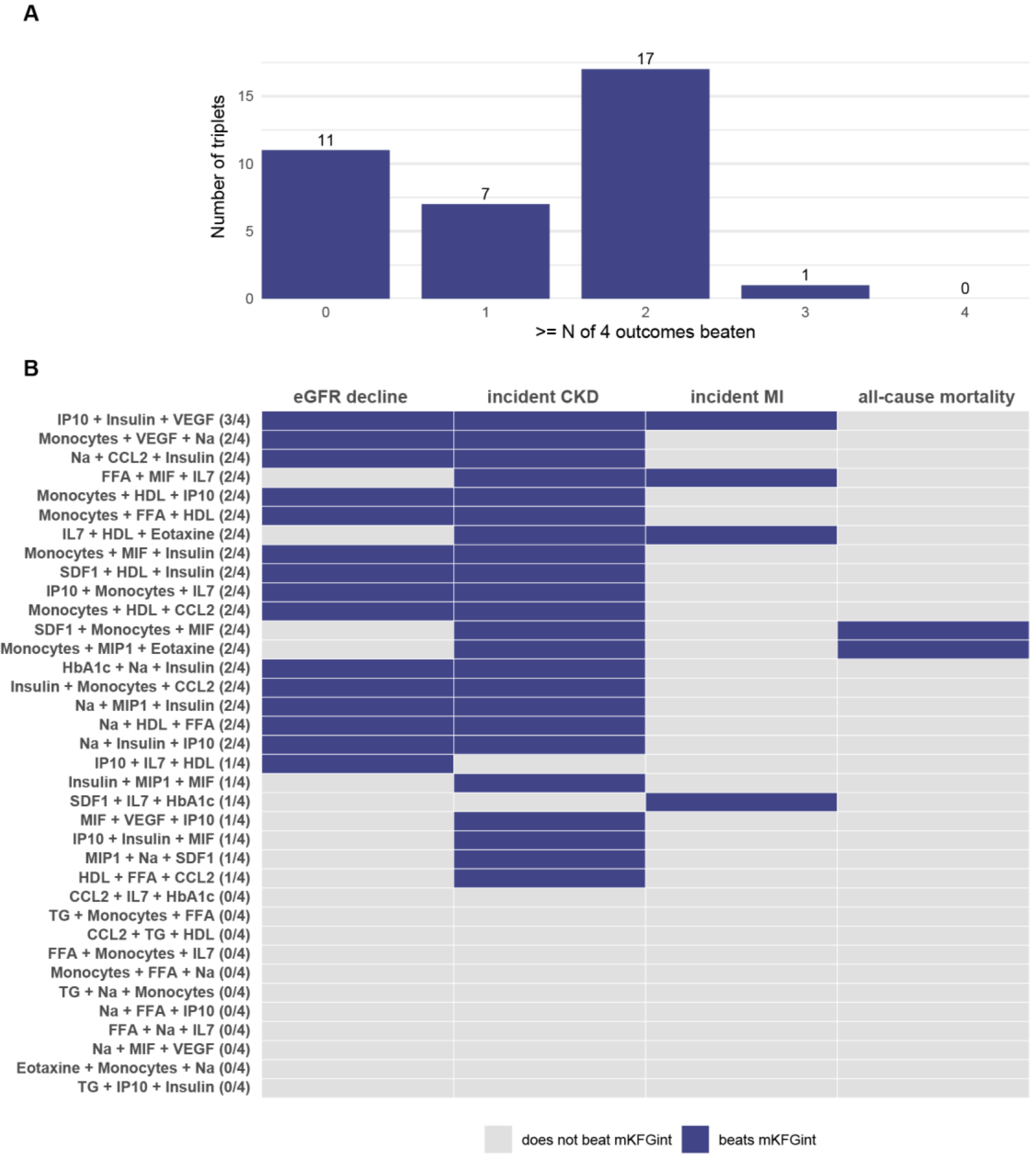

A
Number of triplets
>= N of 4 outcomes beaten
11
7
17
1
0
B
eGFR decline
incident CKD
incident MI
all-cause mortality
IP10 + Insulin + VEGF (3/4)
Monocytes + VEGF + Na (2/4)
Na + CCL2 + Insulin (2/4)
FFA + MIF + IL7 (2/4)
Monocytes + HDL + IP10 (2/4)
Monocytes + FFA + HDL (2/4)
IL7 + HDL + Eotaxine (2/4)
Monocytes + MIF + Insulin (2/4)
SDF1 + HDL + Insulin (2/4)
IP10 + Monocytes + IL7 (2/4)
Monocytes + HDL + CCL2 (2/4)
SDF1 + Monocytes + MIF (2/4)
Monocytes + MIP1 + Eotaxine (2/4)
HbA1c + Na + Insulin (2/4)
Insulin + Monocytes + CCL2 (2/4)
Na + MIP1 + Insulin (2/4)
Na + HDL + FFA (2/4)
Na + Insulin + IP10 (2/4)
IP10 + IL7 + HDL (1/4)
Insulin + MIP1 + MIF (1/4)
SDF1 + IL7 + HbA1c (1/4)
MIF + VEGF + IP10 (1/4)
IP10 + Insulin + MIF (1/4)
MIP1 + Na + SDF1 (1/4)
HDL + FFA + CCL2 (1/4)
CCL2 + IL7 + HbA1c (0/4)
TG + Monocytes + FFA (0/4)
CCL2 + TG + HDL (0/4)
FFA + Monocytes + IL7 (0/4)
Monocytes + FFA + Na (0/4)
TG + Na + Monocytes (0/4)
Na + FFA + IP10 (0/4)
FFA + Na + IL7 (0/4)
Na + MIF + VEGF (0/4)
Eotaxine + Monocytes + Na (0/4)
TG + IP10 + Insulin (0/4)
does not beat mKFGint
beats mKFGint


**Suppl.FIG.11**

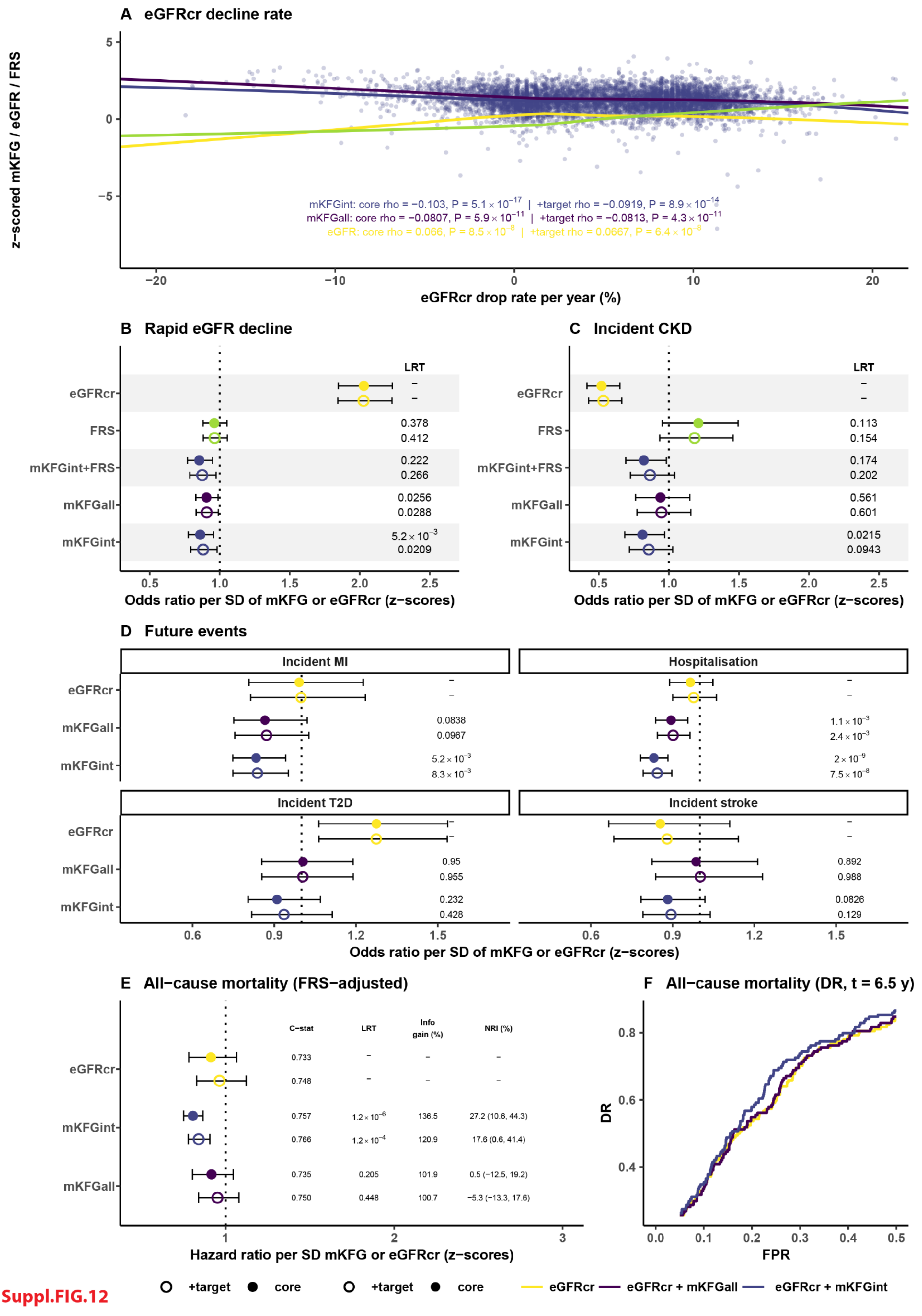
A eGFRcr decline rate
z−scored mKFG / eGFR / FRS
mKFGint: core rho = −0.103, P = 5.1 × 10−17 | +target rho = −0.0919, P = 8.9 × 10−14
mKFGall: core rho = −0.0807, P = 5.9 × 10−11 | +target rho = −0.0813, P = 4.3 × 10−11
eGFR: core rho = 0.066, P = 8.5 × 10−8 | +target rho = 0.0667, P = 6.4 × 10−8
eGFRcr drop rate per year (%)
B Rapid eGFR decline
C Incident CKD
LRT
eGFRcr
FRS
mKFGint+FRS
mKFGall
mKFGint
0.378
0.412
0.222
0.266
0.0256
0.0288
5.2 × 10−3
0.0209
0.113
0.154
0.174
0.202
0.561
0.601
0.0215
0.0943
Odds ratio per SD of mKFG or eGFRcr (z−scores)
D Future events
Incident MI
Hospitalisation
Incident T2D
Incident stroke
0.0838
0.0967
5.2 × 10−3
8.3 × 10−3
1.1 × 10−3
2.4 × 10−3
2 × 10−9
7.5 × 10−8
0.95
0.955
0.232
0.428
0.892
0.988
0.0826
0.129
E All−cause mortality (FRS−adjusted)
C−stat
LRT
Info gain (%)
NRI (%)
0.733
0.748
0.757
1.2 × 10−6
136.5
27.2 (10.6, 44.3)
0.766
1.2 × 10−4
120.9
17.6 (0.6, 41.4)
0.735
0.205
101.9
0.5 (−12.5, 19.2)
0.750
0.448
100.7
−5.3 (−13.3, 17.6)
Hazard ratio per SD mKFG or eGFRcr (z−scores)
F All−cause mortality (DR, t = 6.5 y)
DR
FPR
+target
core
eGFRcr
eGFRcr + mKFGall
eGFRcr + mKFGint

Suppl.FIG.12

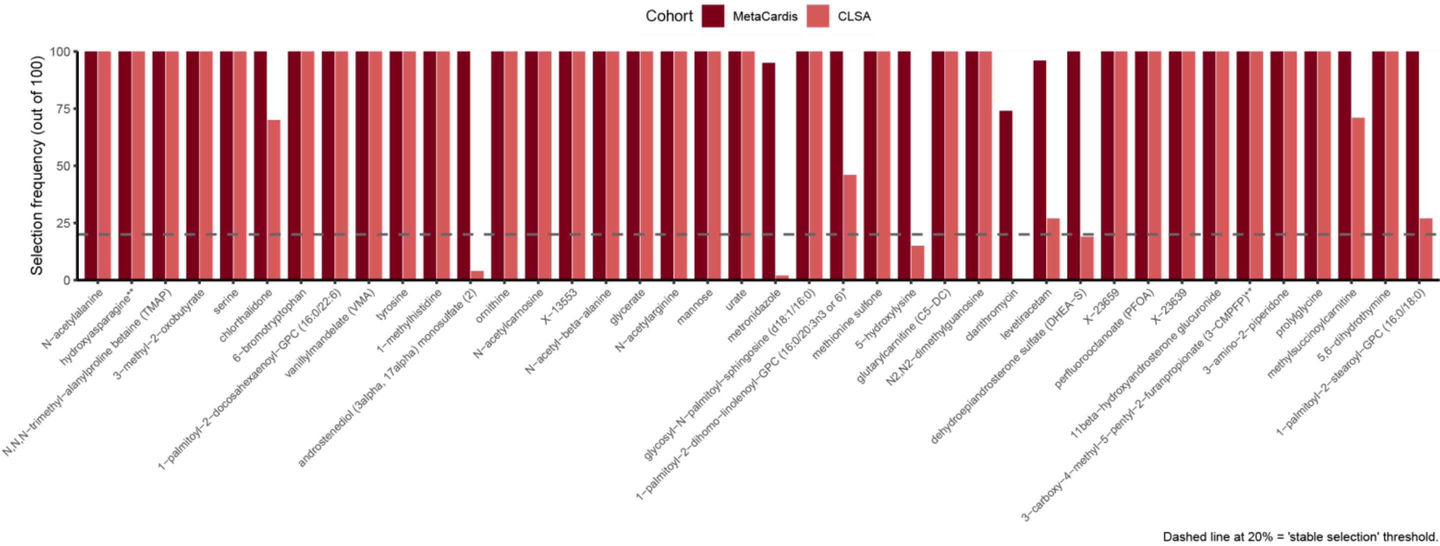

Cohort
MetaCardis
CLSA
Selection frequency (out of 100)
100
75
50
25
0
N−acetylalanine
hydroxyasparagine**
N,N,N−trimethyl−alanylproline betaine (TMAP)
3−methyl−2−oxobutyrate
serine
chlorthalidone
6−bromotryptophan
1−palmitoyl−2−docosahexaenoyl−GPC (16:0/22:6)
vanillylmandelate (VMA)
tyrosine
1−methylhistidine
androstenediol (3alpha, 17alpha) monosulfate (2)
ornithine
N−acetylcarnosine
X−13553
N−acetyl−beta−alanine
glycerate
N−acetylarginine
mannose
urate
metronidazole
glycosyl−N−palmitoyl−sphingosine (d18:1/16:0)
1−palmitoyl−2−dihomo−linolenoyl−GPC (16:0/20:3n3 or 6)*
methionine sulfone
5−hydroxylysine
glutarylcarnitine (C5−DC)
N2,N2−dimethylguanosine
clarithromycin
levetiracetam
dehydroepiandrosterone sulfate (DHEA−S)
X−23659
perfluorooctanoate (PFOA)
X−23639
11beta−hydroxyandrosterone glucuronide
3−carboxy−4−methyl−5−pentyl−2−furanpropionate (3−CMPFP)**
3−amino−2−piperidone
prolylglycine
methylsuccinoylcarnitine
5,6−dihydrothymine
1−palmitoyl−2−stearoyl−GPC (16:0/18:0)
Dashed line at 20% = 'stable selection' threshold.


**Suppl.FIG.13**

A

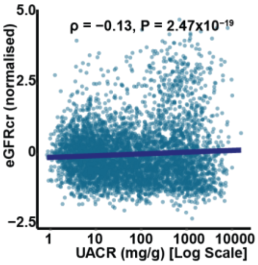


B

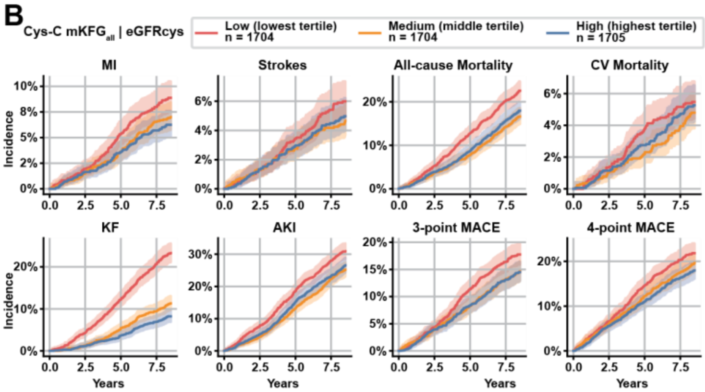


C

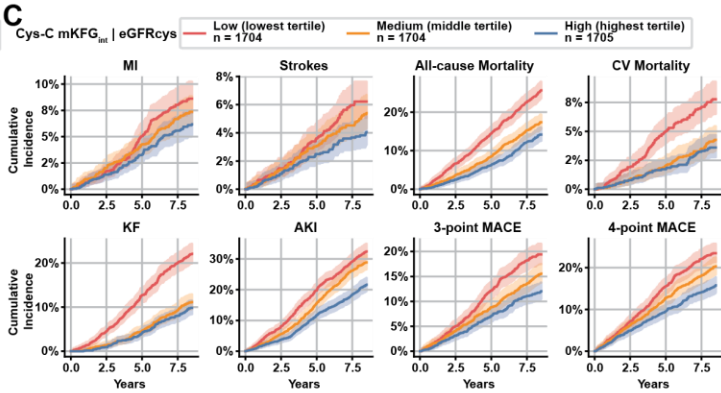


D

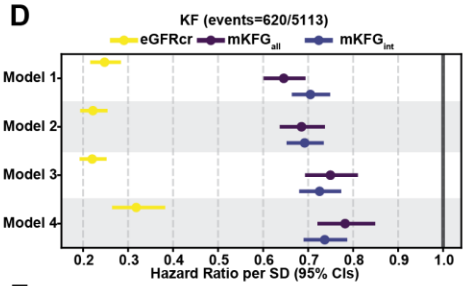


| p-value | Information Gain (%) | C-statistic | NRI 5 years (95% CI) |
|---|---|---|---|
| | | 0.783 (0.763 - 0.804) | |
| <0.0001 | 178.84 | 0.801 (0.781 - 0.822) | 12.5% (-0.1% - 28.0%) |
| <0.0001 | 167.74 | 0.802 (0.783 - 0.822) | 8.2% (-11.2% - 21.8%) |
| | | 0.819 (0.800 - 0.837) | |
| <0.0001 | 138.81 | 0.833 (0.815 - 0.851) | 5.0% (-8.0% - 19.7%) |
| <0.0001 | 143.36 | 0.842 (0.825 - 0.859) | 13.1% (-4.9% - 24.0%) |
| | | 0.847 (0.830 - 0.865) | |
| <0.0001 | 114.65 | 0.856 (0.839 - 0.872) | -4.5% (-16.1% - 10.1%) |
| <0.0001 | 116.23 | 0.864 (0.848 - 0.879) | 9.8% (-10.0% - 29.3%) |
| | | 0.852 (0.835 - 0.870) | |
| <0.0001 | 105.81 | 0.858 (0.842 - 0.875) | 5.3% (-8.2% - 23.4%) |
| <0.0001 | 108.48 | 0.868 (0.852 - 0.883) | 15.3% (1.6% - 29.1%) |

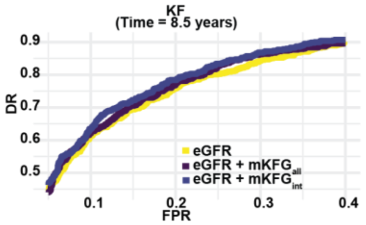


E

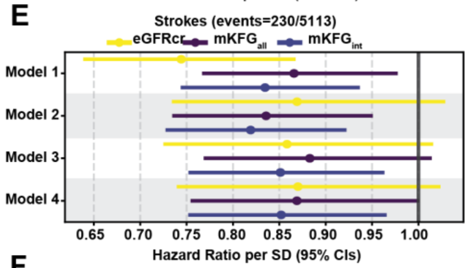


| p-value | Information Gain (%) | C-statistic | NRI 5 years (95% CI) |
|---|---|---|---|
| | | 0.582 (0.545 - 0.619) | |
| 0.0293 | 155.36 | 0.602 (0.565 - 0.639) | 1.7% (-16.5% - 20.0%) |
| 0.0035 | 187.46 | 0.623 (0.586 - 0.660) | 16.5% (-3.3% - 37.6%) |
| | | 0.678 (0.644 - 0.711) | |
| 0.0105 | 116.69 | 0.691 (0.657 - 0.725) | 7.6% (-8.1% - 22.5%) |
| 0.0017 | 124.5 | 0.701 (0.668 - 0.734) | 19.7% (-1.8% - 38.0%) |
| | | 0.685 (0.651 - 0.720) | |
| 0.0902 | 105.3 | 0.692 (0.657 - 0.726) | 1.8% (-11.2% - 26.4%) |
| 0.0138 | 110.94 | 0.701 (0.667 - 0.734) | 11.5% (-6.8% - 31.7%) |
| | | 0.694 (0.659 - 0.729) | |
| 0.0614 | 105.48 | 0.699 (0.664 - 0.735) | 1.2% (-12.7% - 23.0%) |
| 0.0154 | 109.0 | 0.706 (0.672 - 0.741) | 11.4% (-5.7% - 30.6%) |

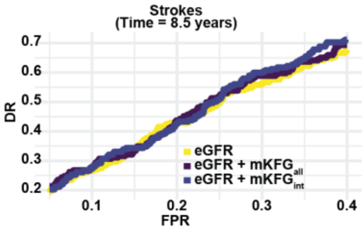


F

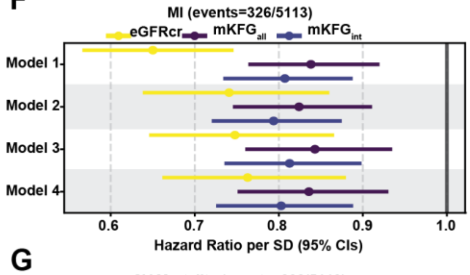


| p-value | Information Gain (%) | C-statistic | NRI 5 years (95% CI) |
|---|---|---|---|
| | | 0.613 (0.582 - 0.644) | |
| <0.0010 | 161.9 | 0.632 (0.602 - 0.663) | 18.0% (2.5% - 30.2%) |
| <0.0001 | 177 | 0.639 (0.609 - 0.670) | 14.2% (4.0% - 32.8%) |
| | | 0.708 (0.679 - 0.736) | |
| <0.0010 | 119.05 | 0.716 (0.688 - 0.744) | 14.9% (-2.5% - 25.4%) |
| <0.0001 | 127.65 | 0.724 (0.697 - 0.752) | 14.0% (-0.9% - 35.7%) |
| | | 0.715 (0.686 - 0.744) | |
| 0.0026 | 110.86 | 0.722 (0.694 - 0.750) | 12.8% (-7.4% - 26.4%) |
| <0.0010 | 116.11 | 0.730 (0.702 - 0.757) | 12.1% (-0.7% - 33.2%) |
| | | 0.728 (0.699 - 0.757) | |
| 0.0023 | 110.03 | 0.737 (0.709 - 0.765) | 10.8% (-4.5% - 24.6%) |
| <0.0001 | 116.2 | 0.745 (0.717 - 0.773) | 16.7% (4.7% - 31.1%) |

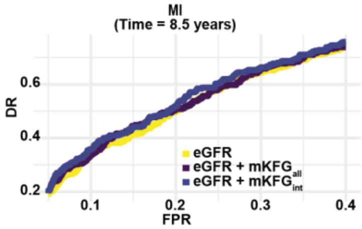


G

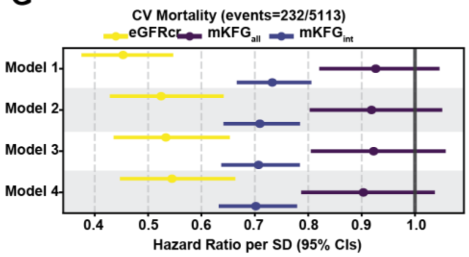


| p-value | Information Gain (%) | C-statistic | NRI 5 years (95% CI) |
|---|---|---|---|
| | | 0.678 (0.643 - 0.714) | |
| 0.2323 | 109.7 | 0.682 (0.647 - 0.718) | 6.6% (-13.2% - 22.1%) |
| <0.0001 | 205.98 | 0.721 (0.687 - 0.755) | 37.0% (21.4% - 55.7%) |
| | | 0.788 (0.762 - 0.815) | |
| 0.2268 | 102.88 | 0.790 (0.764 - 0.816) | 0.8% (-15.1% - 26.0%) |
| <0.0001 | 140.69 | 0.812 (0.787 - 0.837) | 41.1% (20.6% - 56.5%) |
| | | 0.791 (0.764 - 0.818) | |
| 0.2572 | 102.54 | 0.792 (0.765 - 0.819) | 1.2% (-13.7% - 23.4%) |
| <0.0001 | 139.01 | 0.814 (0.788 - 0.839) | 38.2% (18.3% - 52.9%) |
| | | 0.803 (0.776 - 0.830) | |
| 0.1597 | 102.79 | 0.805 (0.779 - 0.832) | 2.5% (-14.5% - 23.6%) |
| <0.0001 | 134.83 | 0.830 (0.805 - 0.856) | 38.3% (22.6% - 56.3%) |

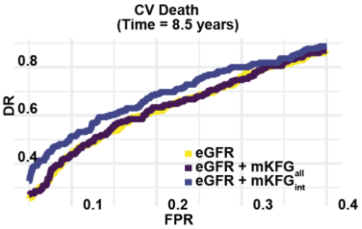


Suppl.FIG.14

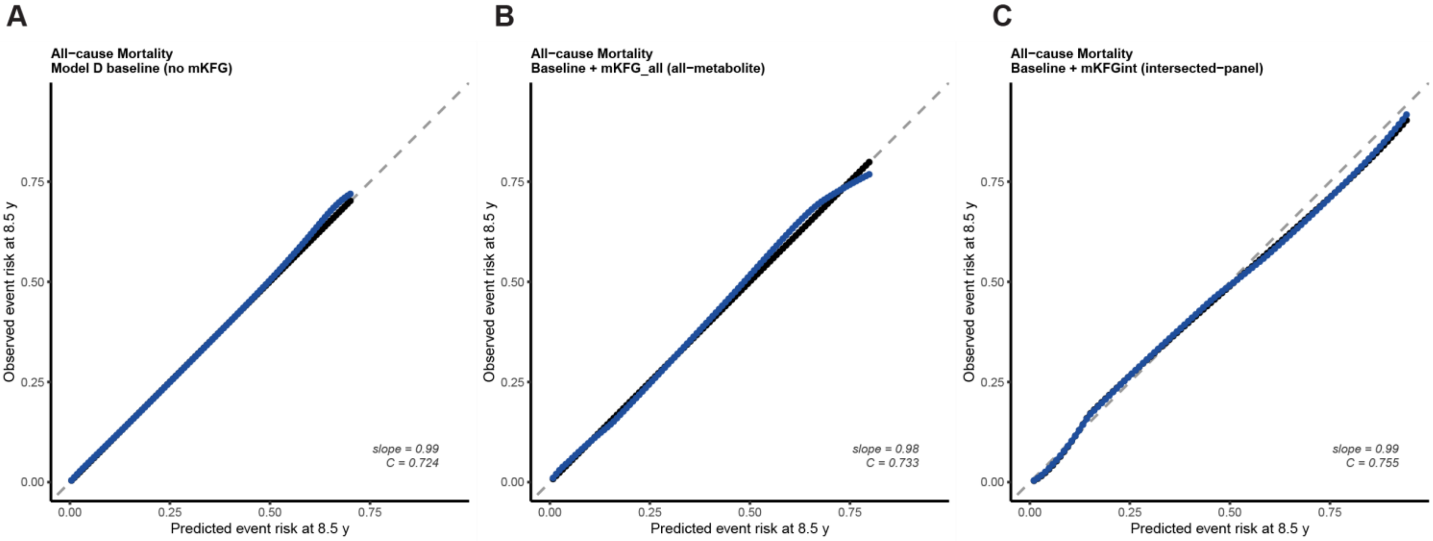

A
All−cause Mortality
Model D baseline (no mKFG)
Observed event risk at 8.5 y
Predicted event risk at 8.5 y
0.00
0.25
0.50
0.75
slope = 0.99
C = 0.724
B
All−cause Mortality
Baseline + mKFG_all (all−metabolite)
Observed event risk at 8.5 y
Predicted event risk at 8.5 y
slope = 0.98
C = 0.733
C
All−cause Mortality
Baseline + mKFGint (intersected−panel)
Observed event risk at 8.5 y
Predicted event risk at 8.5 y
slope = 0.99
C = 0.755


**Suppl.FIG.15**

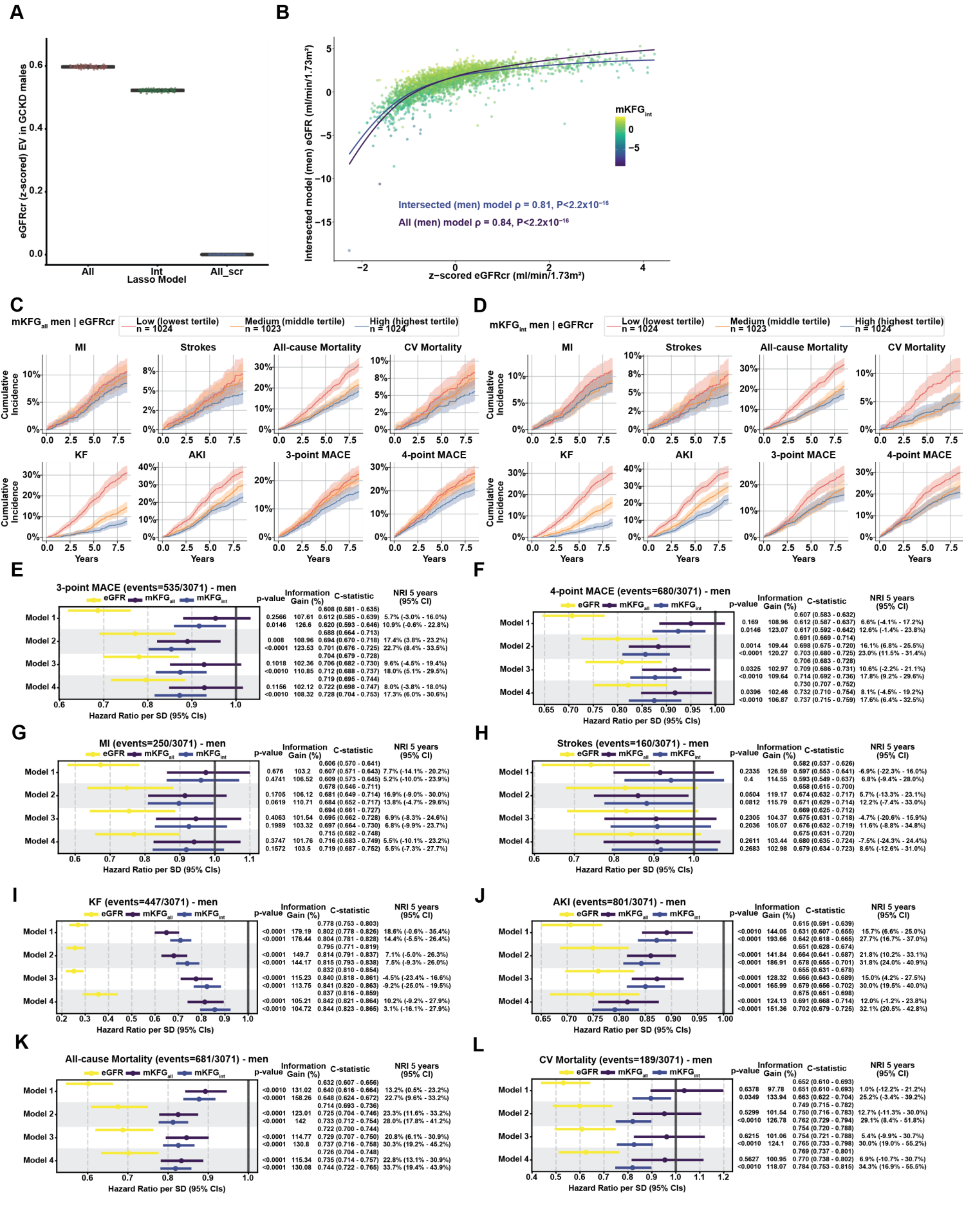
A
eGFRcr (z-scored) EV in GCKD males
Lasso Model
All
Int
All_scr
B
Intersected model (men) eGFR (ml/min/1.73m²)
z−scored eGFRcr (ml/min/1.73m²)
Intersected (men) model ρ = 0.81, P<2.2x10⁻¹⁶
All (men) model ρ = 0.84, P<2.2x10⁻¹⁶
C
mKFGall men | eGFRcr
Low (lowest tertile) n = 1024
Medium (middle tertile) n = 1023
High (highest tertile) n = 1024
MI
Strokes
All-cause Mortality
CV Mortality
KF
AKI
3-point MACE
4-point MACE
Cumulative Incidence
Years
D
mKFGint men | eGFRcr
E
3-point MACE (events=535/3071) - men
F
4-point MACE (events=680/3071) - men
G
MI (events=250/3071) - men
H
Strokes (events=160/3071) - men
I
KF (events=447/3071) - men
J
AKI (events=801/3071) - men
K
All-cause Mortality (events=681/3071) - men
L
CV Mortality (events=189/3071) - men
eGFR
mKFGall
mKFGint
p-value
Information Gain (%)
C-statistic
NRI 5 years (95% CI)
Model 1
Model 2
Model 3
Model 4
Hazard Ratio per SD (95% CIs)

Suppl.FIG.16

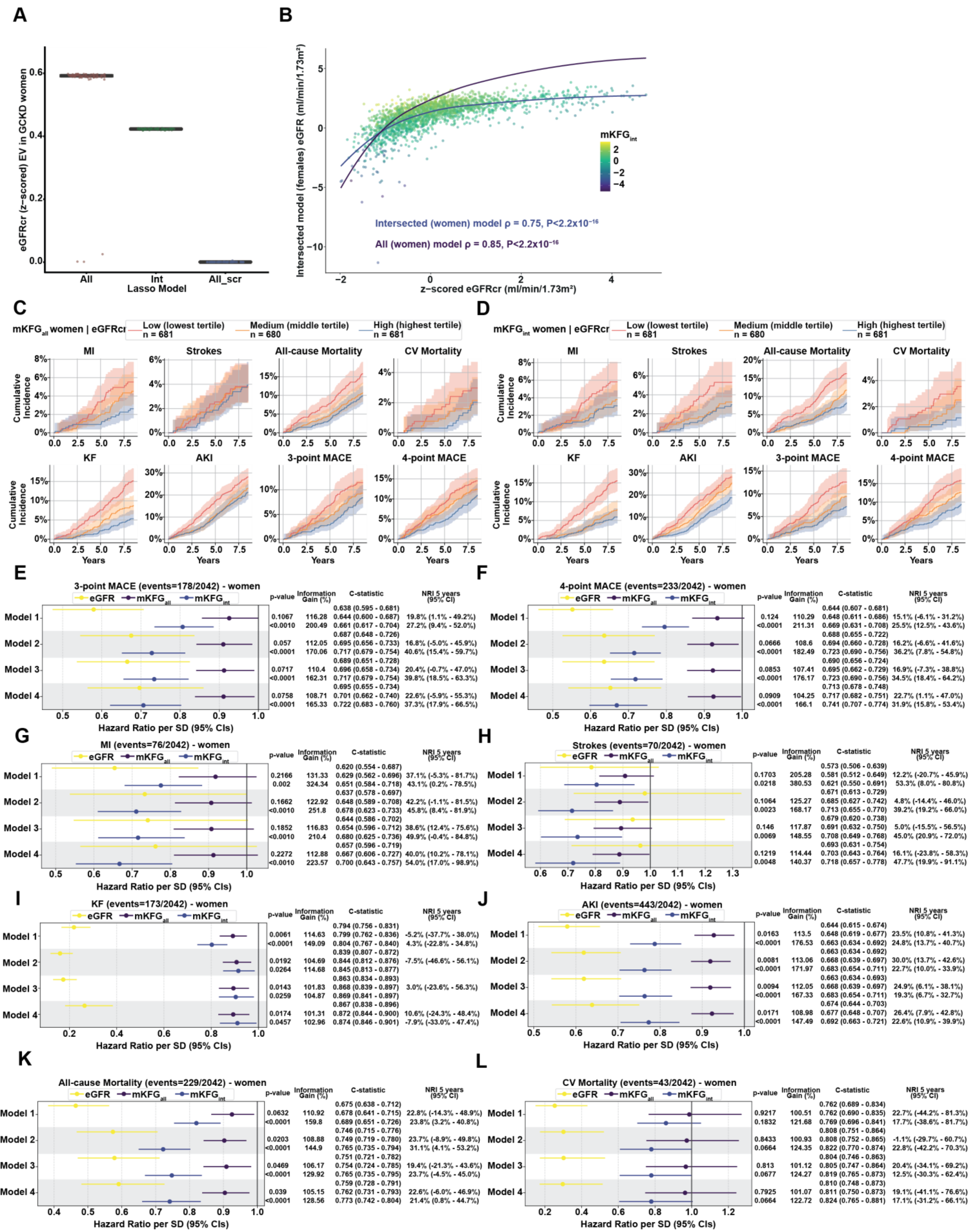
A
eGFRcr (z-scored) EV in GCKD women
Lasso Model
All
Int
All_scr
B
Intersected model (females) eGFR (ml/min/1.73m²)
z−scored eGFRcr (ml/min/1.73m²)
mKFGint
Intersected (women) model ρ = 0.75, P<2.2x10−16
All (women) model ρ = 0.85, P<2.2x10−16
C
mKFGall women | eGFRcr
Low (lowest tertile) n = 681
Medium (middle tertile) n = 680
High (highest tertile) n = 681
MI
Strokes
All-cause Mortality
CV Mortality
KF
AKI
3-point MACE
4-point MACE
Cumulative Incidence
Years
D
mKFGint women | eGFRcr
Low (lowest tertile) n = 681
Medium (middle tertile) n = 680
High (highest tertile) n = 681
E
3-point MACE (events=178/2042) - women
eGFR
mKFGall
mKFGint
p-value
Information Gain (%)
C-statistic
NRI 5 years (95% CI)
Model 1
Model 2
Model 3
Model 4
0.638 (0.595 - 0.681)
0.1067 116.28 0.644 (0.600 - 0.687) 19.8% (1.1% - 49.2%)
<0.0010 200.49 0.661 (0.617 - 0.704) 27.2% (9.4% - 52.0%)
0.687 (0.648 - 0.726)
0.057 112.05 0.695 (0.656 - 0.733) 16.8% (-5.0% - 45.9%)
<0.0010 170.06 0.717 (0.679 - 0.754) 40.6% (15.4% - 59.7%)
0.689 (0.651 - 0.728)
0.0717 110.4 0.696 (0.658 - 0.734) 20.4% (-0.7% - 47.0%)
<0.0001 162.31 0.717 (0.679 - 0.754) 39.8% (18.5% - 63.3%)
0.695 (0.655 - 0.734)
0.0758 108.71 0.701 (0.662 - 0.740) 22.6% (-5.9% - 55.3%)
<0.0001 165.33 0.722 (0.683 - 0.760) 37.3% (17.9% - 66.5%)
Hazard Ratio per SD (95% CIs)
F
4-point MACE (events=233/2042) - women
0.644 (0.607 - 0.681)
0.124 110.29 0.648 (0.611 - 0.686) 15.1% (-6.1% - 31.2%)
<0.0001 211.31 0.669 (0.631 - 0.708) 25.5% (12.5% - 43.6%)
0.688 (0.655 - 0.722)
0.0666 108.6 0.694 (0.660 - 0.728) 16.2% (-6.6% - 41.6%)
<0.0001 182.49 0.723 (0.690 - 0.756) 36.2% (7.8% - 54.8%)
0.690 (0.656 - 0.724)
0.0853 107.41 0.695 (0.662 - 0.729) 16.9% (-7.3% - 38.8%)
<0.0001 176.17 0.723 (0.690 - 0.756) 34.5% (18.4% - 64.2%)
0.713 (0.678 - 0.748)
0.0909 104.25 0.717 (0.682 - 0.751) 22.7% (1.1% - 47.0%)
<0.0001 166.1 0.741 (0.707 - 0.774) 31.9% (15.8% - 53.4%)
G
MI (events=76/2042) - women
0.620 (0.554 - 0.687)
0.2166 131.33 0.629 (0.562 - 0.696) 37.1% (-5.3% - 81.7%)
0.002 324.34 0.651 (0.584 - 0.718) 43.1% (0.2% - 78.5%)
0.637 (0.578 - 0.697)
0.1662 122.92 0.648 (0.589 - 0.708) 42.2% (-1.1% - 81.5%)
<0.0010 251.8 0.678 (0.623 - 0.733) 45.8% (8.4% - 81.9%)
0.644 (0.586 - 0.702)
0.1852 116.83 0.654 (0.596 - 0.712) 38.6% (12.4% - 75.6%)
<0.0010 210.4 0.680 (0.625 - 0.736) 49.9% (-0.4% - 84.8%)
0.657 (0.596 - 0.719)
0.2272 112.88 0.667 (0.606 - 0.727) 40.0% (10.2% - 78.1%)
<0.0010 223.57 0.700 (0.643 - 0.757) 54.0% (17.0% - 98.9%)
H
Strokes (events=70/2042) - women
0.573 (0.506 - 0.639)
0.1703 205.28 0.581 (0.512 - 0.649) 12.2% (-20.7% - 45.9%)
0.0218 380.53 0.621 (0.550 - 0.691) 53.3% (8.0% - 80.8%)
0.671 (0.613 - 0.729)
0.1064 125.27 0.685 (0.627 - 0.742) 4.8% (-14.4% - 46.0%)
0.0023 168.17 0.713 (0.655 - 0.770) 39.2% (19.2% - 66.0%)
0.679 (0.620 - 0.738)
0.146 117.87 0.691 (0.632 - 0.750) 5.0% (-15.5% - 56.5%)
0.0069 148.55 0.708 (0.649 - 0.768) 45.0% (20.9% - 72.0%)
0.693 (0.631 - 0.754)
0.1219 114.44 0.703 (0.643 - 0.764) 16.1% (-23.8% - 58.3%)
0.0048 140.37 0.718 (0.657 - 0.778) 47.7% (19.9% - 91.1%)
I
KF (events=173/2042) - women
0.794 (0.756 - 0.831)
0.0061 114.63 0.799 (0.762 - 0.836) -5.2% (-37.7% - 38.0%)
<0.0001 149.09 0.804 (0.767 - 0.840) 4.3% (-22.8% - 34.8%)
0.839 (0.807 - 0.872)
0.0192 104.69 0.844 (0.812 - 0.876) -7.5% (-46.6% - 56.1%)
0.0264 114.68 0.845 (0.813 - 0.877)
0.863 (0.834 - 0.893)
0.0143 101.83 0.868 (0.839 - 0.897) 3.0% (-23.6% - 56.3%)
0.0259 104.87 0.869 (0.841 - 0.897)
0.867 (0.838 - 0.896)
0.0174 101.31 0.872 (0.844 - 0.900) 10.6% (-24.3% - 48.4%)
0.0457 102.96 0.874 (0.846 - 0.901) -7.9% (-33.0% - 47.4%)
J
AKI (events=443/2042) - women
0.644 (0.615 - 0.674)
0.0163 113.5 0.648 (0.619 - 0.677) 23.5% (10.8% - 41.3%)
<0.0001 176.53 0.663 (0.634 - 0.692) 24.8% (13.7% - 40.7%)
0.663 (0.634 - 0.692)
0.0081 113.06 0.668 (0.639 - 0.697) 30.0% (13.7% - 42.6%)
<0.0001 171.97 0.683 (0.654 - 0.711) 22.7% (10.0% - 33.9%)
0.663 (0.634 - 0.693)
0.0094 112.05 0.668 (0.639 - 0.697) 24.9% (6.1% - 38.1%)
<0.0001 167.33 0.683 (0.654 - 0.711) 19.3% (6.7% - 32.7%)
0.674 (0.644 - 0.703)
0.0171 108.98 0.677 (0.648 - 0.707) 26.4% (7.9% - 42.8%)
<0.0001 147.49 0.692 (0.663 - 0.721) 22.6% (10.9% - 39.9%)
K
All-cause Mortality (events=229/2042) - women
0.675 (0.638 - 0.712)
0.0632 110.92 0.678 (0.641 - 0.715) 22.8% (-14.3% - 48.9%)
<0.0001 159.8 0.689 (0.651 - 0.726) 23.8% (3.2% - 40.8%)
0.746 (0.715 - 0.776)
0.0203 108.88 0.749 (0.719 - 0.780) 23.7% (-8.9% - 49.8%)
<0.0001 144.9 0.765 (0.735 - 0.794) 31.1% (4.1% - 53.2%)
0.751 (0.721 - 0.782)
0.0469 106.17 0.754 (0.724 - 0.785) 19.4% (-21.3% - 43.6%)
<0.0001 129.92 0.765 (0.735 - 0.795) 23.7% (-4.5% - 45.0%)
0.759 (0.728 - 0.791)
0.039 105.15 0.762 (0.731 - 0.793) 22.6% (-6.0% - 46.9%)
<0.0001 128.56 0.773 (0.742 - 0.804) 21.4% (0.8% - 44.7%)
L
CV Mortality (events=43/2042) - women
0.762 (0.689 - 0.834)
0.9217 100.51 0.762 (0.690 - 0.835) 22.7% (-44.2% - 81.3%)
0.1832 121.68 0.769 (0.696 - 0.841) 17.7% (-38.6% - 81.7%)
0.808 (0.751 - 0.864)
0.8433 100.93 0.808 (0.752 - 0.865) -1.1% (-29.7% - 60.7%)
0.0664 124.35 0.822 (0.770 - 0.874) 22.8% (-42.2% - 70.3%)
0.804 (0.746 - 0.863)
0.813 101.12 0.805 (0.747 - 0.864) 20.4% (-34.1% - 69.2%)
0.0677 124.27 0.819 (0.765 - 0.873) 12.5% (-30.3% - 62.4%)
0.810 (0.748 - 0.873)
0.7925 101.07 0.811 (0.750 - 0.873) 19.1% (-41.1% - 76.6%)
0.0664 122.72 0.824 (0.765 - 0.881) 17.1% (-31.2% - 66.1%)


Suppl.FIG.17
